# Rare protein-coding variation and the genetic architecture of height in >1.4 million individuals

**DOI:** 10.64898/2026.06.22.26355163

**Authors:** Jack A. Kosmicki, Liron Ganel, Kyoko Watanabe, Tyler Joseph, Sheila M. Gaynor, Michael D. Kessler, Joshua D. Backman, Joelle Mbatchou, Andrey Ziyatdinov, David Blair, Jonas Bovijn, Niek Verweij, Kathie Y. Sun, Chuanyi Zhang, Suganthi Balasubramanian, Adrián I. Campos, Alexander W. Charney, Olle Melander, Robert Weinreb, Jason Torres, Pablo Kuri-Morales, Roberto Tapia-Conyer, Jesús Alegre-Díaz, Jaime Berumen, BELIEVE Study Group, Colorado Center for Personalized Medicine - RGC Collaboration, GHS-RGC DiscovEHR Collaboration, Mayo Clinic Project Generation, México City Prospective Study, Penn Medicine BioBank, Regeneron Genetics Center, University of California Los Angeles-RGC ATLAS Collaboration, Aris Baras, Luca A. Lotta, Jonathan Marchini, Gonçalo R. Abecasis, Manuel A. R. Ferreira, Adam E. Locke

**Author notes:** J.A. Kosmicki and L. Ganel equally contributed to this work. G.R. Abecasis, M.A.R. Ferreira, and A.E. Locke jointly supervised this work. A full list of authors and their affiliations is provided in the Supplementary Note (BELIEVE Study Group, Colorado Center for Personalized Medicine - RGC Collaboration, GHS-RGC DiscovEHR Collaboration, Mayo Clinic Project Generation, México City Prospective Study, Penn Medicine BioBank, Regeneron Genetics Center, University of California Los Angeles-RGC ATLAS Collaboration).

## Abstract

Highly heritable, polygenic, and easily measured, adult height has long been the model trait in human genetics^1,2^. While the landscape of height-associated common genetic variation has been studied extensively^2^, rare variation remains relatively unexplored^1^. Using rare protein-altering variants in a discovery set of 826,066 exomes, we identify 207 height-associated genes - 98% of which replicate in an additional 624,567 individuals. The rarest and most deleterious class of variation, singleton (frequency <0.0001%) putative loss-of-function (pLoF) variants implicated 17 genes with large effects on height ranging from -17 cm (*ACAN*) to +11 cm (*FBN1*) per allele, 52× larger than the average effect of common height-associated variants and comparable to the 1% tails of a common variant polygenic score. Several genes (e.g., *TET1*, *DTL*, *IGF2BP2*) have effect sizes at least as large as established Mendelian height genes but lack documented stature or skeletal growth syndromes. This is particularly true for genes in which rare variants associate with increased height. We performed the largest rare-variant study of height to date, directly implicate 207 genes that broadly overlap with both GWAS associations and Mendelian height syndromes, assess the impact of rare variants on heritability and prediction, provide evidence that height is an underappreciated clinical feature of Mendelian disorders, and demonstrate the utility of large population-scale sequencing studies for classifying individual variants and dissecting complex trait architecture.

## Introduction

Although height was described as a heritable trait as early as 1886^3^, it was not until the late 20^th^ century that researchers began to uncover its genetic architecture by identifying the genetic causes of Mendelian syndromes with a significant stature component^4–7^. Identification of non-syndromic genetic modulators of height remained largely elusive until 2007 with the introduction of genome-wide association studies (GWAS) that identified the first common variants associated with height^8^. Fifteen years later, a height GWAS comprising 5.3 million individuals identified 12,111 independent variants that collectively accounted for 40% of the variance in height across individuals of European descent^2^. As with other traits, height GWAS have largely implicated common noncoding variants with modest effect sizes (<1 cm)^2^, making it challenging to directly link associations to specific genes and functional mechanisms.

In contrast, sequencing-based studies of rare variation have the potential to directly implicate causal genes by identifying large-effect loss-of-function and protein altering variants, improve trait prediction accuracy, and uncover new biology^9^. To systematically assess the contribution of rare coding variation to height, we carried out a multi-ancestry exome sequencing analysis of adult height in 1,450,633 individuals across 12 cohorts (**Supplementary Table 1**), in which we 1) identify 17 genes where the burden of singleton pLoFs significantly associates with height; 2) identify additional height-associated genes by including additional pLoF and nonsynonymous variants up to 1% alternate allele frequency (AAF) and including non-burden style tests; and 3) estimate the impact of rare protein-altering variation on genetic-based height prediction.

## Results

Using 826,066 individuals from six cohorts (54.8% from the UK Biobank [UKB]; 77.5% EUR; **Supplementary Table 1**), we performed three discovery association analyses of adult height – 1) individual imputed common variants [AAF>0.01], 2) individual rare exome variants (AAF≤0.01, alternate allele count [AAC]≥10), and 3) gene-based tests grouping rare coding variants within each gene. We used a single, unified Bonferroni significance threshold of 1.75×10^-9^ across all three analyses (**Methods**). For gene-based analyses, we employed a gene-wide omnibus statistic, gene-P^10^, that aggregates evidence across burden tests (which assume similar effects for all variants in a group) and two other gene-based tests, ACAT-V^11^ and SKAT-O^12^, which allow variability in effect size and direction (**Supplementary Figure 1**; **Methods**). We used 624,567 additional individuals from six cohorts for replication analyses (**Table 1**) and assessed the impact of rare variants in height prediction models using a subset of these additional individuals (N = 242,679).

**Table 1:** 17 genes associated with height via singleton pLoF burden tests in 826,066 individuals.

| Gene symbol | $\beta$ (SE) [cm] | P-value | Carriers | Function | OMIM syndrome(s) |
| --- | --- | --- | --- | --- | --- |
| *ACAN*<sup>$\alpha,\beta,\gamma,\delta$</sup> | -16.55 (1.09) | $1.87 \times 10^{-52}$ | 42 | ECM, proteoglycan | Spondyloepiphyseal dysplasia |
| *ANKRD11*<sup>$\alpha,\gamma,\delta$</sup> | -11.16 (1.71) | $7.48 \times 10^{-11}$ | 17 | Transcription | KBG syndrome |
| *COL1A1*<sup>$\beta,\gamma,\delta$</sup> | -10.36 (1.62) | $1.51 \times 10^{-10}$ | 19 | ECM | Ehlers-Danlos syndrome, OI |
| *EXT1*<sup>$\gamma,\delta$</sup> | -8.7 (1.29) | $1.40 \times 10^{-11}$ | 30 | ECM, GAG biosynthesis | Multiple osteochondromas |
| *IGF2BP2*<sup>$\alpha,\beta,\gamma$</sup> | -8.68 (1.36) | $1.52 \times 10^{-10}$ | 27 | IGF | - |
| *ADAMTS6*<sup>$\alpha,\beta,\gamma$</sup> | -8.4 (1.35) | $5.53 \times 10^{-10}$ | 27 | ECM | - |
| *ADAMTS10*<sup>$\alpha,\beta,\gamma,\delta$</sup> | -8.24 (1.36) | $1.35 \times 10^{-9}$ | 27 | ECM | Weill-Marchesani syndrome |
| *IGF1R*<sup>$\alpha,\beta,\gamma,\delta$</sup> | -8.23 (0.97) | $1.75 \times 10^{-17}$ | 53 | IGF | IGF1 resistance |
| *DTL*<sup>$\alpha,\beta$</sup> | -7.34 (1.08) | $1.28 \times 10^{-11}$ | 42 | TGF-β, DNA damage repair | - |
| *SCUBE3*<sup>$\alpha,\beta,\gamma,\delta$</sup> | -7.18 (1.0) | $7.08 \times 10^{-13}$ | 49 | ECM, TGF-β | Short stature/skeletal anomalies |
| *NF1*<sup>$\beta,\gamma,\delta$</sup> | -5.55 (0.6) | $1.33 \times 10^{-20}$ | 140 | ECM | NF1 |
| *NRK*<sup>$\alpha,\beta$</sup> | 3.79 (0.62) | $1.29 \times 10^{-9}$ | 56 | Transcription | - |
| *ZFAT*<sup>$\alpha,\beta$</sup> | 7.86 (1.1) | $9.58 \times 10^{-13}$ | 41 | Transcription | Autoimmune thyroid disease |
| *TET1*<sup>$\alpha,\beta,\gamma$</sup> | 8.32 (1.09) | $2.23 \times 10^{-14}$ | 42 | Epigenetic mark maintenance | - |
| *LCORL*<sup>$\alpha,\beta,\gamma$</sup> | 9.99 (0.84) | $6.21 \times 10^{-33}$ | 71 | Transcription | - |
| *CHD8*<sup>$\alpha,\gamma$</sup> | 10.22 (1.54) | $3.44 \times 10^{-11}$ | 21 | Transcription, Wnt/β-catenin | ASD, ID, macrocephaly |
| *FBN1*<sup>$\alpha,\gamma,\delta$</sup> | 11.14 (1.11) | $1.45 \times 10^{-23}$ | 40 | ECM, TGF-β | Marfan syndrome |

<sup>$\alpha$</sup> nearest gene to the GWAS locus; <sup>$\beta$</sup> associated with standing height in Backman *et al.*<sup>9</sup>;
<sup>$\gamma$</sup> constrained (based on LOEUF<sup>18</sup>); <sup>$\delta$</sup> OMIM gene for abnormal stature or skeletal growth in
Yengo *et al.*<sup>2</sup>; SE: standard error; ASD: autism spectrum disorder; ECM: extra-cellular matrix;
GAG: glycosaminoglycan; ID: intellectual disability; IGF: insulin-like growth factor pathway;
NF1: neurofibromatosis, type 1; OI: osteogenesis imperfecta; TGF-β : transforming growth factor
beta pathway; P-values for singleton pLoF burden tests come from REGENIE.

Common variant association analysis identified 3,034 conditionally independent height associations (**Supplementary Table 2**) that when combined in a polygenic score (PGS) explained ∼25% of the observed variation in height in replication data (**Methods**; **Supplementary Note**). We conditioned on these 3,034 common variants to ensure rare variant and gene-based associations were independent of common signals (**Supplementary Note**; **Supplementary Figure 2**). We identified 105 rare nonsynonymous variants in 87 genes (median AAF = 2.16×10^-3^; **Supplementary Table 3**) and 207 genes associated with height via gene-based aggregate tests (**Extended Data Figure 1A**; **Supplementary Table 4**). Eighty-one of the 85 genes with a significantly associated rare nonsynonymous variant were also among the 207 genes identified via gene-based tests.

### 17 genes associated with height through singleton pLoF variants

Since singleton pLoFs represent the rarest and putatively most deleterious class of variants, we first focused on this subset of rare coding variation. Of the 207 genes associated with height, singleton pLoF burden tests identified 17 (11 height-decreasing, six height-increasing; **Figures 1A** & **1B**; **Table 1**; **Supplementary Tables 5 & 6**) with a mean absolute effect of 8.92 cm per allele (**Figure 1C**). Effects ranged from a 16.6 cm decrease in stature for *ACAN* singleton pLoF carriers to an 11.1 cm increase in stature for *FBN1* singleton pLoF carriers. Four genes - *ANKRD11*, *CHD8*, *EXT1*, and *FBN1* - were not previously identified via burden tests (*P*<1.75×10^-9^) for standing height in 454,787 UKB participants^9^. Furthermore, both *CHD8* (associated with autism spectrum disorder and intellectual disability) and *COL1A1* (associated with Ehlers-Danlos syndrome^13^ and osteogenesis imperfecta) were only identified via singleton pLoF burdens; when higher frequency pLoFs in these genes were included, the effect sizes plummeted by 78% for *COL1A1* and 83% for *CHD8* and were no longer statistically significant (**Figure 1A**). These 17 genes belong to well-known height biological processes and pathways including extracellular matrix organization and skeletal/growth pathways such as growth hormone/insulin-like growth factor (GH/IGF), Wnt/β-catenin, and transforming growth factor β (TGF-β**; Tables 1** & **2**)^1,14–16^.

**Figure 1:**
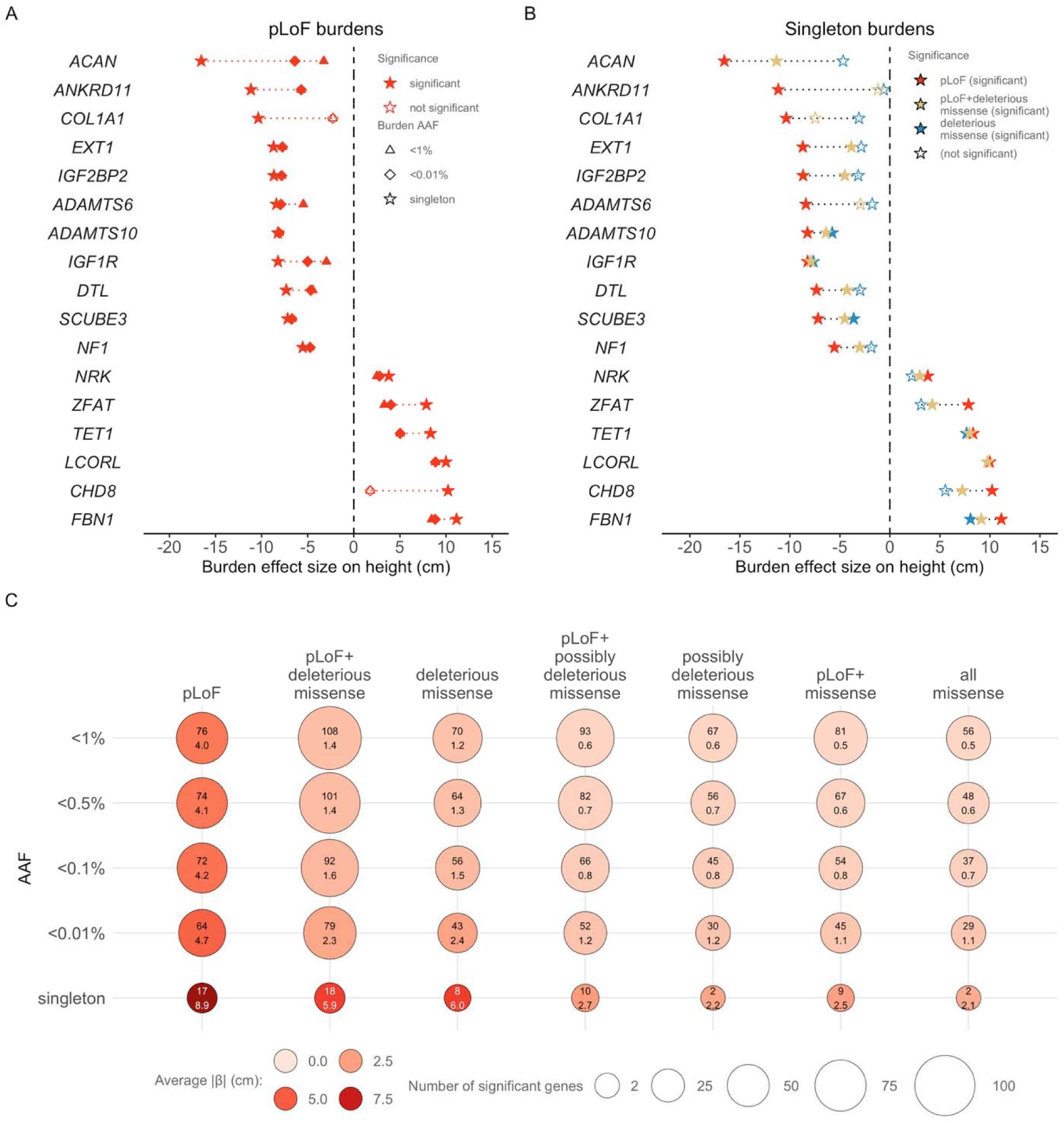
**A**) pLoF burden effect sizes across AAFs for 17 genes identified by a significant burden of singleton pLoFs (**Table 1**). **B**) Same as **A** but only effect sizes from singleton burden tests are displayed. **C**) Relationship between AAF, variant class, effect size, and discovery yield for 179 genes identified from burden tests (**Supplementary Tables 6 and 10)**. Inside each point is the number of genes discovered (top) and the mean effect (in cm; bottom). Effect sizes were estimated by multiplying by the standard deviation of observed height (∼7.99 cm). Significance was assessed using a *P*-value threshold of 1.75×10^-9^.

Prior height GWAS^15^ consistently observed an enrichment of Mendelian height syndrome genes among height-associated common variant loci^17^. We observed a similar enrichment in genes implicated through rare variation: eleven of the 17 genes have a documented Mendelian disorder in the Online Mendelian Inheritance in Man (OMIM) database (**Table 1**), a 6.24-fold enrichment for Mendelian disease genes compared to the remaining genes in the genome (95% CI=[2.11, 20.56]; *P =* 2.55×10^-4^; Fisher’s exact test; see **Supplementary Note** for sensitivity analyses). Nine of the eleven OMIM entries include a stature or skeletal growth component among reported clinical characteristics (compared to all OMIM genes: OR=38.55; 95% CI=7.55-367.77; *P* = 7.19×10^-8^; Fisher’s exact test)^2^. Among the eleven genes where rare singleton pLoFs are associated with decreased stature, eight (73%) have an OMIM entry describing a syndrome that includes stature or skeletal growth as a feature^2^. In contrast, of the six genes where singleton pLoFs are associated with increased height, only *FBN1* includes such an entry and three genes (*LCORL*, *NRK*, and *TET1)* have no Mendelian syndrome entry in OMIM. This suggests increased stature might be under-reported as a clinical characteristic or might co-occur less often with deleterious health outcomes typical of OMIM syndromes. To assess whether this is simply due to a difference in pleiotropy, we compared the number of traits from a phenome-wide association study using burden tests for >2,000 UKB phenotypes^9^ and observed no difference in the average number of trait associations between height-increasing and height-decreasing pLoF singleton genes (10.17 [95% CI=0-22.39] versus 8.91 [95% CI=3.96-13.86], respectively; *P*=0.54; two-sided Mann-Whitney U test; **Supplementary Note**). Consistent with their overrepresentation amongst OMIM genes and indicative of strong negative selection, 14 of these 17 genes are intolerant to loss-of-function variation (i.e., LoF-constrained^18^) – 11× more likely than the remaining 190 gene-P genes (95% CI=[2.51, 106.77]; *P*=2.34×10^-4^; Fisher’s exact test; **Methods**; **Supplementary Note**) and 7-fold more likely than the 76 genes with a significant pLoF < 1% burden (95% CI=[1.54, 72.05]; *P*=0.005; Fisher’s exact test). We observed no difference in constraint between height-increasing and height-decreasing singleton pLoF genes (*P*=1; Fisher’s exact test; **Supplementary Note**). However, notable exceptions such as *DTL* and *ZFAT* show large effects on height despite being tolerant to loss-of-function variation.

Previous studies estimated that the gene nearest to a GWAS peak is the causal gene in ∼70% of instances^9,19^. With this criterion, our 3,034 GWAS index variants implicate 2,127 protein-coding genes (∼11% of all protein-coding genes in the genome). All but three (*COL1A1*, *EXT1*, *NF1*) of the 17 pLoF singleton genes were also the gene nearest to a GWAS peak (**Supplementary Table 2**; **Supplementary Figure 3**; OR=36.41; 95% CI=[10.14, 197.74]; *P =* 3.01×10^-11^; Fisher’s exact test), reinforcing prior observations that common noncoding variants and rare, large-effect coding variants largely implicate the same genes^9,20,21^. We note that prior work suggests GWAS and burden tests tend to rank these genes differently^21^.

Given the extensive overlap between genes implicated by common and rare variants, we systematically compared the effect of common variants with those of singleton pLoFs. The average effect of a singleton pLoF in these 17 genes is 8.92 cm (**Figure 1C**), ∼52× larger than the average GWAS variant (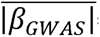=0.17 cm) and ∼44× larger (range 14.4×–78.3×) when comparing effects gene-by-gene (**Supplementary Table 7**; **Methods**). Furthermore, the effect of carrying a singleton pLoF in one of these 17 genes approaches that of the 1% tails of the common variant PGS (top 1%: +9.60 cm; 95% CI=[9.34, 9.86 cm]; bottom 1%: -9.12 cm; 95% CI=[-9.33, -8.91 cm]; **Figure 2A**; **Supplementary Tables 8** & **9**; **Methods**). These incredibly rare pLoFs, cumulatively carried by fewer than 0.1% of individuals (744 carriers in 826,066 individuals), have a larger impact on height than all but the most extreme combinations of common variants.

**Figure 2:**
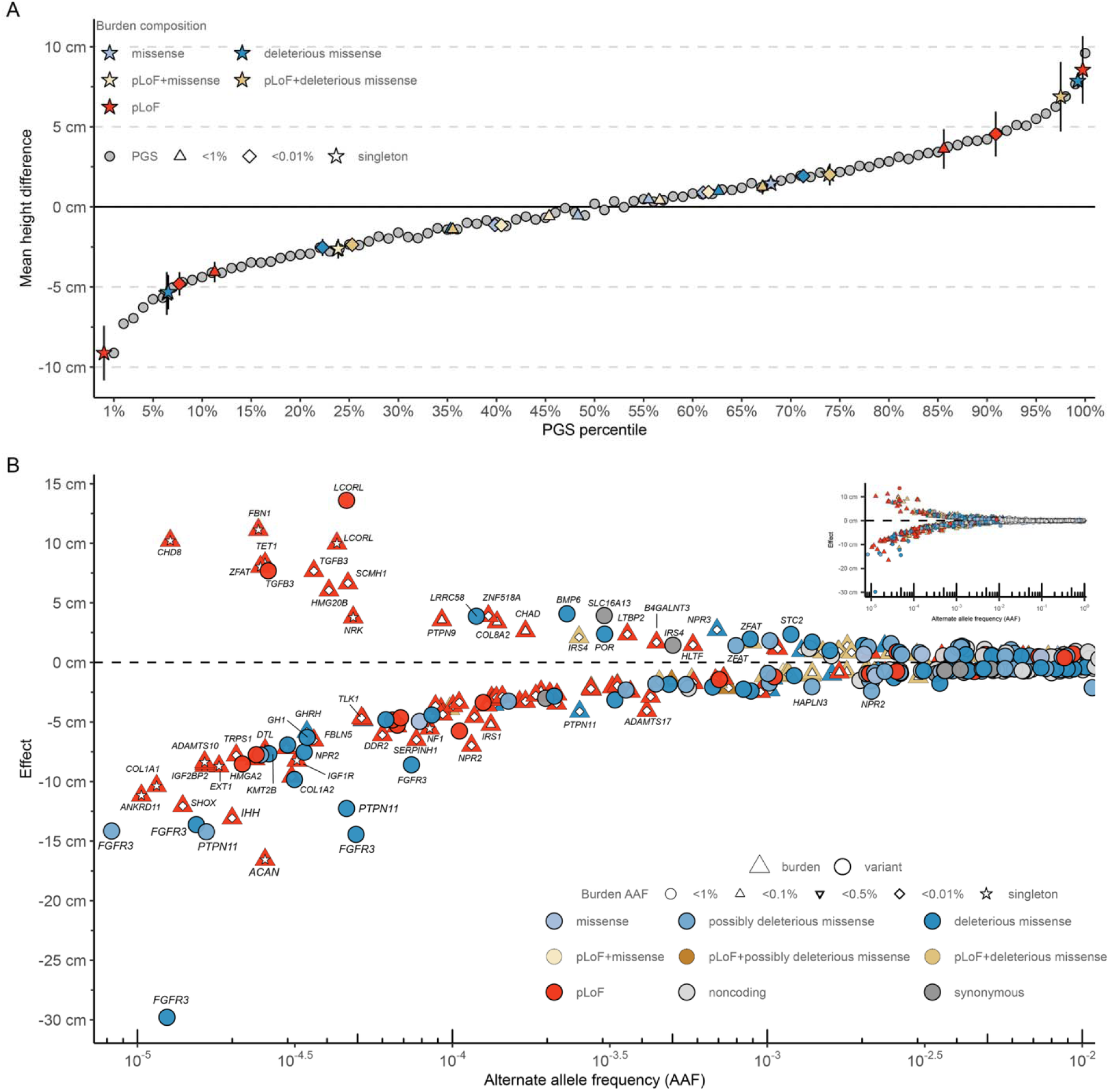
**A**) Percentiles of a height polygenic score (PGS; grey) derived from common (AAF>1%) variants (3,034 GWAS variants + 5 common exome variants; **Supplementary Table 8**) representing the difference in the average height of individuals in each percentile compared to the rest of the ancestry/sex-matched dataset (**Supplementary Table 9**). Average effect sizes for the significant burden tests (split by direction of effect) are placed in comparison to the PGS. Error bars represent 95% confidence intervals (CI) around the mean effect estimated from REGENIE^132^. **B**) The inverse relationship between effect size (in cm; y-axis) and AAF (x-axis) for burden tests (**Supplementary Table 6**), exome variants (**Supplementary Table 3**), and conditionally independent common variants (**Supplementary Table 2**). Burden tests are denoted with triangles (with a smaller shape for the burden AAF threshold embedded inside), individual variants with circles. The color denotes the predicted consequence of individual variants or variants included in the burden test. The largest, Bonferroni significant associations observed in the discovery analysis come from individual rare variants, not burden tests. The full distribution is depicted in the inset.

### Expanding analysis from singletons to low frequency pLoFs

Incorporating more common variation into burden tests can increase statistical power for gene discovery by adding variant carriers. We sought to identify and characterize additional height-associated genes by expanding our analysis to include increasingly frequent pLoFs then by including missense variants. In total, these analyses identified an additional 162 genes (**Extended Data Figure 1**).

Among pLoFs, relaxing the burden test AAF threshold from singletons (∼0.00006%) to 1% identified 76 associated genes, 62 (82%) of which were not discovered with pLoF singletons alone. On average, burden tests aggregating more common pLoFs had a 3.96 cm effect - less than half the 8.92 cm effect of pLoF singletons, but still ∼24× larger than the average GWAS association (**Figure 1C**; **Supplementary Table 10**). Among the 15 genes with pLoF singleton associations that remained significant at 0.01%, we observed a 35% attenuation in the effect size (from 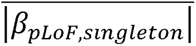=8.71 cm to 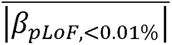=5.68 cm), suggesting a substantial fraction of more frequent pLoF-annotated variants may not represent complete loss-of-function. Nevertheless, we identified three genes with absolute effects on height >1 SD (7.99 cm) not observed in the pLoF singleton analysis: *SHOX*^22^ in the fibroblast growth factor pathway (-12.05 cm; *P* = 2.85×10^-16^), transcriptional regulator *ERF* (-8.67 cm; *P* = 4.41×10^-21^), and *HMGA2,* the insulin-like growth factor pathway gene underlying the first height locus discovered via GWAS^8^ (-8.11 cm; *P* = 6.11×10^-13^). These three genes each had very few singleton pLoF variants (1, 6, and 16 carriers, respectively). Therefore, aggregating singletons with additional rare pLoFs was essential to achieve significance.

This expanded set of height-associated pLoF burden genes highlighted additional canonical growth pathways and biology (e.g., extracellular matrix organization and fibroblast growth factor pathway; **Table 2)**, as well as additional members from the same gene families and pathways implicated by pLoF singleton genes (**Supplementary Table 11**). We observed that 43 of 76 genes (57%) with significant pLoF burden signals at an AAF<1% had a documented Mendelian syndrome in OMIM, significantly more than expected (OR = 4.46; 95% CI = [2.76, 7.26]; *P* = 1.83×10^-10^; Fisher’s exact test). Of the 43 genes, 21 had annotated stature or skeletal growth features^2^, an 8.37-fold enrichment (95% CI = [4.34, 16.10]; *P* = 2.56×10^-10^; Fisher’s exact test). Genes with pLoF burden height associations and a Mendelian syndrome listed in OMIM are >5× more likely to be associated with decreased height (36 of 43, 83%) than increased height (7 of 43, 17%), again consistent with the idea that increased height is under-reported as a syndromic feature.

**Table 2:**
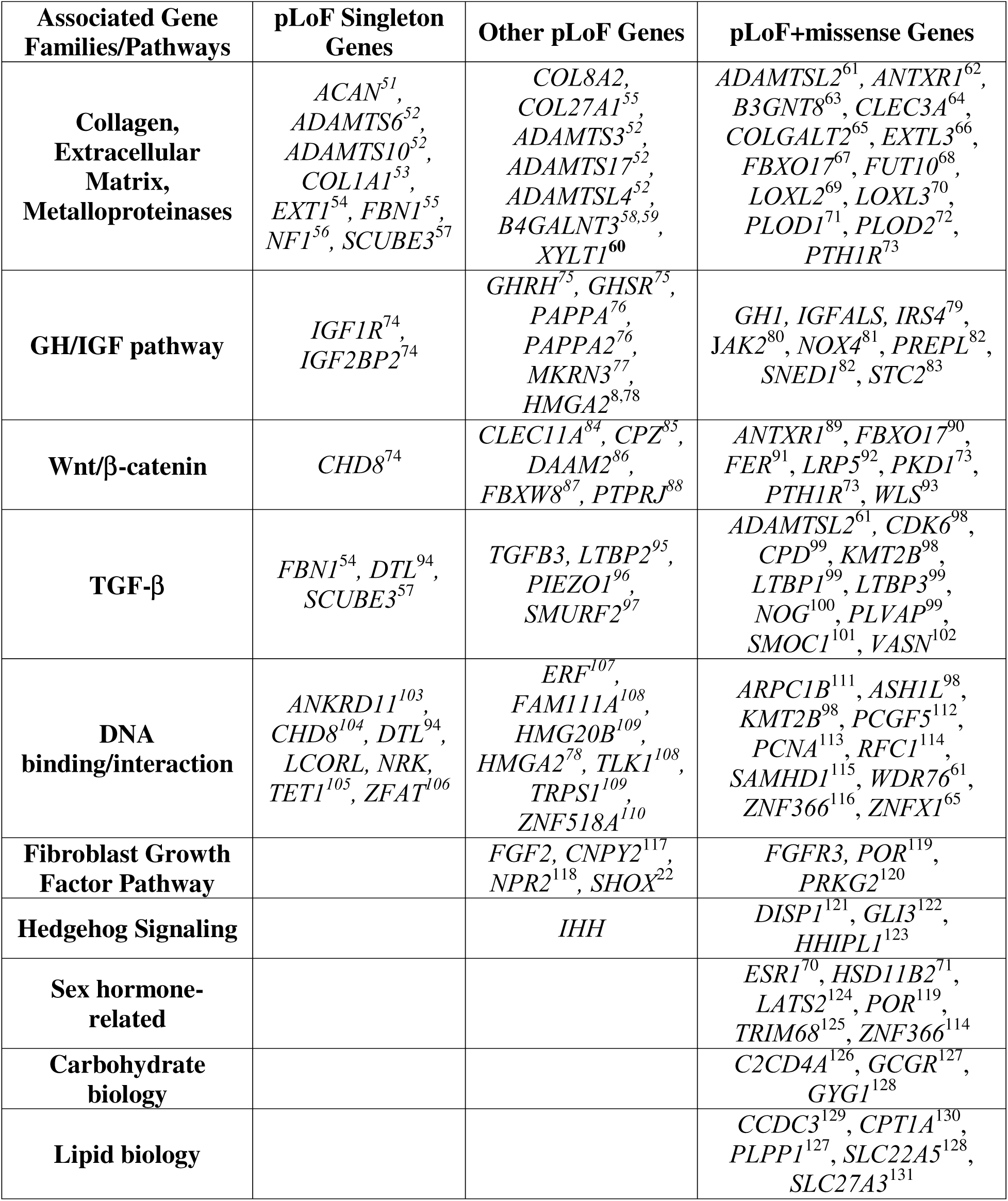
Biological functions and families of genes identified via burden tests. Categories are not mutually exclusive, nor is this intended to be a comprehensive accounting of all genes significantly associated with height. The table represents a high-level description of several trends we observed in the list of genes identified by burden testing of rare variation.

Of the 76 genes identified via pLoF burden tests with AAF<1%, nearly two-thirds (50) are the nearest gene to a lead GWAS variant (OR = 15.2; 95% CI = [9.2, 25.6]; *P* = 3.49×10^-29^; Fisher’s exact test). Lastly, as with singleton pLoFs, genes identified by aggregating low-frequency pLoFs were significantly more likely to be LoF-constrained than the rest of the genome (OR = 3.54; 95% CI = [2.17, 5.76]; *P* = 2.37×10^-7^; Fisher’s exact test).

### Combining pLoFs with missense variants

Whereas pLoFs represent the rarest and most deleterious class of coding variation, missense variants represent the most frequent class of protein altering variants^9,18,23^. If missense variants ablate protein function to a similar extent as pLoFs, we might expect both an increase in statistical power and similar effect size when combining both in burden tests. Conversely, if missense variants only partially diminish protein function, or have opposing effects due to gain of function (GoF) mechanisms, burden effect sizes will be attenuated.

We combined pLoFs with three sets of missense variants: (1) all missense variants, (2) missense variants inconsistently classified as deleterious by *in silico* algorithms (“possibly deleterious missense”), and (3) missense variants confidently classified as deleterious by *in silico* algorithms (“deleterious missense”; **Methods**). Collectively, these tests identified 154 genes associated with height, 85 of which were not discovered by pLoFs alone. Filtering rare missense variants for predicted functional impact increased discovery power. Gene-based tests including pLoFs and all missense variants identified 96 genes, while limiting to those with modest evidence of deleteriousness yielded 116 genes, and further restricting to the most confidently predicted deleterious missense variants increased discovery to 132 genes (**Figure 1C**; **Extended Data Figure 1B**).

Among the 14 genes with both significant singleton pLoF and singleton pLoF and deleterious missense burdens (all significant singleton pLoF genes except *ANKRD11*, *ADAMTS6*, and *COL1A1*; **Table 1**), the absolute effect size decreases 28.5% on average when adding deleterious missense singletons (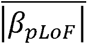=8.70 cm vs. 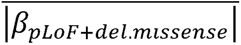=6.22 cm; **Figure 1B**). This observation holds broadly across height-associated genes (**Figure 1C)**, with diminished pLoF+missense effect sizes reflecting both that missense variants account for most rare allele carriers (68.5% in singleton burdens and 84.1% in AAF<1% burden tests) and that, on average, the impact of missense variants is substantially smaller than that of pLoFs (**Figures 1C**; **Supplementary Table 10**). Despite the overall diminished effect sizes, by more than doubling the number of associated height genes, we show that including missense variants increases overall statistical power through higher allele counts.

Of the 85 genes not also discovered via pLoFs alone, *UHRF2* (singleton pLoF & deleterious missense variants: *β*=-4.74 cm; *P* = 3.6×10^-12^) – an E3 ubiquitin ligase cell cycle regulator^24^ – and *PTPN11* (pLoF & deleterious missense variants AAF<0.01%: *β*=-4.00 cm; *P* = 2.5×10^-32^) – a signaling protein involved in development and hormone response through the Ras/MAPK and PI3K/AKT signaling pathways^25^ – had the largest effect sizes.

The new genes identified by incorporating missense variants further underscore previously implicated pathways, biological functions, and gene families (**Supplementary Table 11**), but also implicate different pathways - Sonic hedgehog signaling – and biological patterns – including sex hormones and transport, storage, and metabolism of carbohydrates (**Table 2**).

Among these 154 genes, 76 (49%) are causes of documented Mendelian disorders in OMIM, of which 32 have a stature or skeletal growth component^2^, a 6.50-fold enrichment compared to the remaining OMIM genes (95% CI=[3.95, 10.62]; *P* = 7.66×10^-13^; Fisher’s exact test). Some of these syndromic genes include *PTPN11* (Noonan syndrome), *FGFR3* (achondroplasia), the estrogen receptor *ESR1* (estrogen resistance), and the anthrax receptor *ANTXR1* (GAPO syndrome).

Nearly 60% (92) of the 154 genes are the nearest gene to a GWAS locus, representing a 12-fold enrichment (95% CI=[8.55, 16.86]) relative to the remaining genes in the genome (*P* = 2.25×10^-47^; Fisher’s exact test), once again demonstrating convergence among common and rare variant associations. However, four genes are >1 MB from the nearest common variant association signal: *FIBIN*, *HHIPL1*, *PTPN11,* and *WLS*. Even when compared to the 12,111 index variants from Yengo *et al.*^2^, *PTPN11* is not among the 10 nearest genes to any GWAS peak and the closest is 393 KB away.

### Genes discovered under non-burden gene-based tests and alternative genetic models

While burden tests identified 179 height-associated genes (**Extended Data Figure 1A**), these tests were not designed to identify genes where variants with opposite phenotypic effects co-exist or where a large fraction of variants have no effect. For these situations, SKAT-O^12^ and ACAT-V^11^ tests are expected to be better powered, as they do not assume a similar effect for all variants.

Collectively, ACAT-V and SKAT-O identified 28 genes that were not detected via burden tests (**Extended Data Figure 1A**) after conditioning on common variants (we observed ACAT-V and SKAT-O associations were more likely to be driven by LD with common variants than burden tests; **Supplementary Note**; **Supplementary Figure 2**; **Supplementary Table 12**). Consistent with underlying assumptions, SKAT-O associations highlight genes harboring variants with opposite effects on height. For example, in *PTCH1*, a transmembrane protein receptor involved in embryonic development through the hedgehog signaling pathway^26^, the SKAT-O association (*P* = 1.94×10^-15^) was driven by two variants: a missense variant associated with increased height (9:95458026:G:A; p.T1052M; *β*=1.00 cm; *P* = 4.8×10^-11^; **Supplementary Table 3**), and a splice acceptor variant associated with decreased height (9:95485875:C:T; *β*=-4.28 cm; *P* = 1.9×10^-9^). Meanwhile, ACAT-V identified genes like *FBN2* (*P =* 1.72×10^-12^), a component of microfibrils which store and regulate growth factors including TGF-β^27^, where the association was driven by one significant missense variant (5:128332993:G:T; p.H1381N; *β*=1.07 cm; *P* = 9.8×10^-14^; AAF=0.48%; **Supplementary Table 3**).

We inquired whether any genes were identified solely via a burden of missense but not pLoFs. We found six genes (*ESR1*, *FGFR3*, *HSD11B2*, *MTMR11*, *PTPN11*, *RPL5*) where missense-only burden tests not only exhibited the strongest association, but pLoFs were, at best, only nominally significant (**Supplementary Table 13**). For four of these genes (*FGFR3*, *HSD11B2*, *PTPN11,* and *RPL5*), we observed opposite directions of effect on height between pLoF and missense burden tests, although the pLoF signals were not statistically significant. It is also noteworthy that three of these genes - *FGFR3*^7,28^, *PTPN11*^29^, and *ESR1*^30^ - have previously reported gain-of-function (GoF) mutations that drive syndromic conditions with notable skeletal growth phenotypes, including achondroplasia (*FGFR3*)^28^ and Noonan syndrome (*PTPN11*)^29^. Given the known GoF mechanisms for three of these genes, it seems plausible the other missense variant driven genes may also affect height via GoF mechanisms.

### Individual rare exome variant associations

Although gene-based tests are useful for boosting rare variant discovery power, we still identified 121 significant rare (AAF<1%) exome variants after conditioning on common GWAS variants (**Figure 2B**; **Supplementary Table 3**; see **Supplementary Table 14** and **Supplementary Note** for additional statistically significant nonsynonymous variants that did not meet the AAC<10 threshold). Of these 121 variants, 105 were nonsynonymous (85 missense, 17 pLoFs, 3 inframe indels) and implicated 87 genes, 82 of which overlap genes implicated by gene-based tests (60 genes from burden tests; 22 genes from ACAT-V or SKAT-O).

Having both statistically significant burden tests and individual rare variants for the same gene enabled us to compare effect sizes (**Methods**). Focusing on 79 rare nonsynonymous variants (59 missense, 17 pLoFs, 3 inframe indels) across 60 genes with a significant burden gene-based test, we found 52 (66%) had larger effect sizes than the largest effect burden test. The neurodevelopmental disorder gene *KMT2B*^31^ had the largest discrepancy, with a nearly 20-fold difference between 19:35720959:C:T (p.R538C; -7.67 cm; *P* = 2.6×10^-10^; AAF=2.61×10^-5^) and the largest effect burden test (-0.38 cm; *P* = 5.0×10^-10^; comprising pLoF and possibly deleterious missense variants with an AAF<0.01%).

Two genes with GoF missense variants, *FGFR3* and *PTPN11*, harbor multiple missense variants with effects among the largest observed on height (**Figure 2B**). Among the six exome-wide significant variants in *FGFR3*, a pathogenic GoF missense variant causing achondroplasia^7^ (4:1804392:G:A; p.G380R; *P =* 3.22×10^-64^) decreases height by 29.78 cm (95% CI=[-33.23, - 26.33]), an effect ∼13× larger than the largest *FGFR3* burden test effect (**Extended Data Figure 2**; deleterious missense variants at AAF<0.0001; *β*=-2.31 cm; *P =* 2×10^-41^). *PTPN11* also contains two significant pathogenic GoF missense variants^29,32^, p.R265Q (12:112472981:G:A; *β*=-12.26 cm; 95% CI=[-14.96, -9.55]; *P =* 6×10^-19^) and p.N308D (12:112477719:A:G; *β*=-14.21 cm; 95% CI=[-17.89, -10.53]; *P =* 4×10^-14^), with effects ∼3× larger than the largest burden signal (deleterious missense variants AAF<0.0001; *β*=-4.12 cm; *P =* 3.6×10^-33^). These examples emphasize the challenges presented by allelic heterogeneity in interpreting gene burden effect sizes. We discuss the implications of effect size heterogeneity on phenotypic prediction, later in the section **Rare variant heritability, rare variant enrichment, and impact on height prediction.**

Recent studies^33,34^ demonstrated non-additive models can identify associations missed by standard approaches, so we also tested rare coding variants for association with height under a recessive model. After conditioning on common variants, the recessive model identified only one association missed by additive tests: the *CFTR* ΔF508 mutation that causes cystic fibrosis (CF)^35^ did not appear to associate with height under an additive model (*β_add_* = -0.05 cm; *β_add_*= 0.14), but was associated with a 5.87 cm reduction in height in homozygotes (*β_rec_* = 5×10^-10^; see **Supplementary Note** for replication and additional details). This result is consistent with both the recessive nature of CF mutations and the established impact of CF on growth and stature. The inability of CF treatments to fully restore growth trajectory and stature combined with the observation that newborns with CF exhibit diminished IGF1 levels, are both compatible with *CFTR* having a direct effect on height via fetal or neonatal growth pathways^36,37^.

We sought to understand the differences in genetic architecture across genetic ancestry groups by stratifying the single-variant and gene-based tests by ancestry (**Methods**). We detected no effect size heterogeneity by genetic ancestry, consistent with prior findings^2,9,38^. We discovered several ancestry-specific signals generally due to differences in AAF and sample size. For example, most associations specific to a single ancestry were identified in the European subset (**Supplementary Table 15**; **Supplementary Note**) that accounts for the majority of our data (N=640,240). One notable exception is a rare p.V496E substitution in *HHIP* that was significant only in the AMR subpopulation (AMR AAF = 0.05%; *P*=1.52×10^-9^; *β*=-3.96 cm; **Supplementary Table 15**; **Supplementary Note**). This short-stature associated variant is only observed in individuals from the Mexico City Prospective Study^39^ (MCPS). We speculate that the p.V496E variant is a gain-of-function missense variant, since rare *HHIP* pLoFs appear to be associated with *increased* height (pLoF singletons; *P* = 3.72×10^-7^; β = +9.92 cm). Similarly, we replicate a previously reported *FBN1* missense variant associated with shorter stature in the Peruvian population^40^ (p.E1297G; *β*=-2.12 cm; *P =* 2.9×10^-17^; **Supplementary Table 16**). This variant is substantially more common in admixed Americans (∼1%) than other ancestries (0.004% in Europeans) and occurs almost exclusively on indigenous haplotypes (**Supplementary Figure 4)**. These discoveries underscore the value of sequencing understudied populations for uncovering novel variation.

### Replication in 624,567 additional individuals

To assess the validity of our results, we performed a replication analysis across six cohorts comprising an additional 624,567 sequenced individuals (**Supplementary Table 1**). Despite the 24.4% smaller sample size, 203/207 (98.1%) genes and 113/125 (90.4%) single variants replicated with nominal significance (11 = 0.05; **Supplementary Figure 5**; **Supplementary Tables 3** & **4**; **Supplementary Note**; **Methods**) demonstrating both remarkable consistency in the associations as well as robustness of our Bonferroni threshold. Replication rates were consistent across the different gene-based tests as well: BURDEN-ACAT: 98.1% (155/158), SBAT: 93.8% (166/177), SKAT-O: 95.3% (164/172), and ACAT-V: 94.9% (94/99; **Supplementary Figure 5B**-**E**). Replication performance for single variants improved to 96.1% (98/102) after restricting to variants with >80% estimated replication power (**Supplementary Figure 5F**). Irrespective of power, all 179 burden genes and all but one variant had consistent directions of effect between the discovery and replication analyses (**Supplementary Note**). To further leverage the large replication sample, we evaluated replication rates across a range of *P*-value thresholds (**Supplementary Note**; **Supplementary Figures 5 and 6**). Gene-based tests maintained high replication rates well above our study-wide threshold. For example, at a discovery gene-P threshold of 2.5×10□^6^, roughly equivalent to correcting for one test per gene, 91.6% of genes replicated with nominal significance (*P*<0.05), identifying 368 genes (77.8% more than at our experiment-wide Bonferroni threshold; **Supplementary Figure 5A**). Replication rates for single variants and singleton burden tests overall declined more rapidly, requiring *P*-value thresholds of 4×10□□ and 1×10□□ to achieve a comparable 91.6% replication rate (both restricted to tests with >80% power; **Supplementary Figures 5F and 6**).

Similarly, given that burden tests were much more likely to have negative (height-decreasing) effect estimates, we compared the ratio of effect directions for genes across a range of significance thresholds (**Supplementary Note**; **Extended Data Figure 3**). We note the excess of height-decreasing burden effects persists well below the genome-wide significance threshold, particularly for pLoF burden tests. The degree of imbalance varies both by significance and variant class. Both results are consistent with considerable additional rare coding burden associations remaining to be discovered for height.

### Rare variant heritability, rare variant enrichment, and impact on height prediction

Rare variants are frequently offered as an answer to the “missing heritability” problem in complex trait genetics^41^. Here, we sought to estimate the cumulative impact of rare coding variation on height at the population level by calculating the proportion of phenotypic variance attributable to the burden of rare coding variants and assessing the potential of large-effect height-associated coding variants to improve height predictions.

First, burden heritability regression^42^ estimated that irrespective of *P*-value, the burden of rare nonsynonymous variants explain 3.97% (95% CI=[3.77%, 4.17%]) of phenotypic variance (**Extended Data Figure 4**; **Supplementary Table 17**; **Supplementary Note)**, nearly identical to the published estimate of 3.82% (95% CI=[3.55%, 4.09%])^42^. pLoFs with AAF<0.001% accounted for the largest fraction of burden heritability, consistent with the large effects we observed for these extremely rare, damaging variants. This is likely an underestimate of the true contribution of rare coding variants to height heritability because the burden approach underestimates heritability when there is effect size heterogeneity^42^. Together, the most significant burden test for each of the 179 burden genes explains ∼1.1% of phenotypic variance in the replication data (excluding the *All of Us* cohort), nearly 30% of the total estimated contribution of the burden of rare variants to adult height (**Extended Data Figure 4**).

We next tested for enrichment of rare, deleterious variants in individuals whose height was poorly predicted by common variants in our replication data (N = 242,679, excluding the *All of Us* cohort) as rare variants unaccounted for in the common variant prediction may contribute to the discrepancy in observed and genetically-predicted height^43–45^. Individuals deviating from their common variant predicted height by 2.5-3 SDs (± ∼17-20 cm) were 22.7-fold (95% CI = [11.3, 41.7]; *P =* 1.29×10^-12^; Fisher’s exact test) more likely to carry a singleton pLoF in one of the 17 singleton pLoF genes (**Figure 3A**; **Supplementary Table 18**). In comparison, individuals at the extreme ends of the height distribution (2.5-3 SDs), were only 4.8-fold enriched (*P*=2.14×10^-5^; Fisher’s exact test; **Supplementary Table 19**; **Supplementary Note**), consistent with previous results^43,45^.

**Figure 3:**
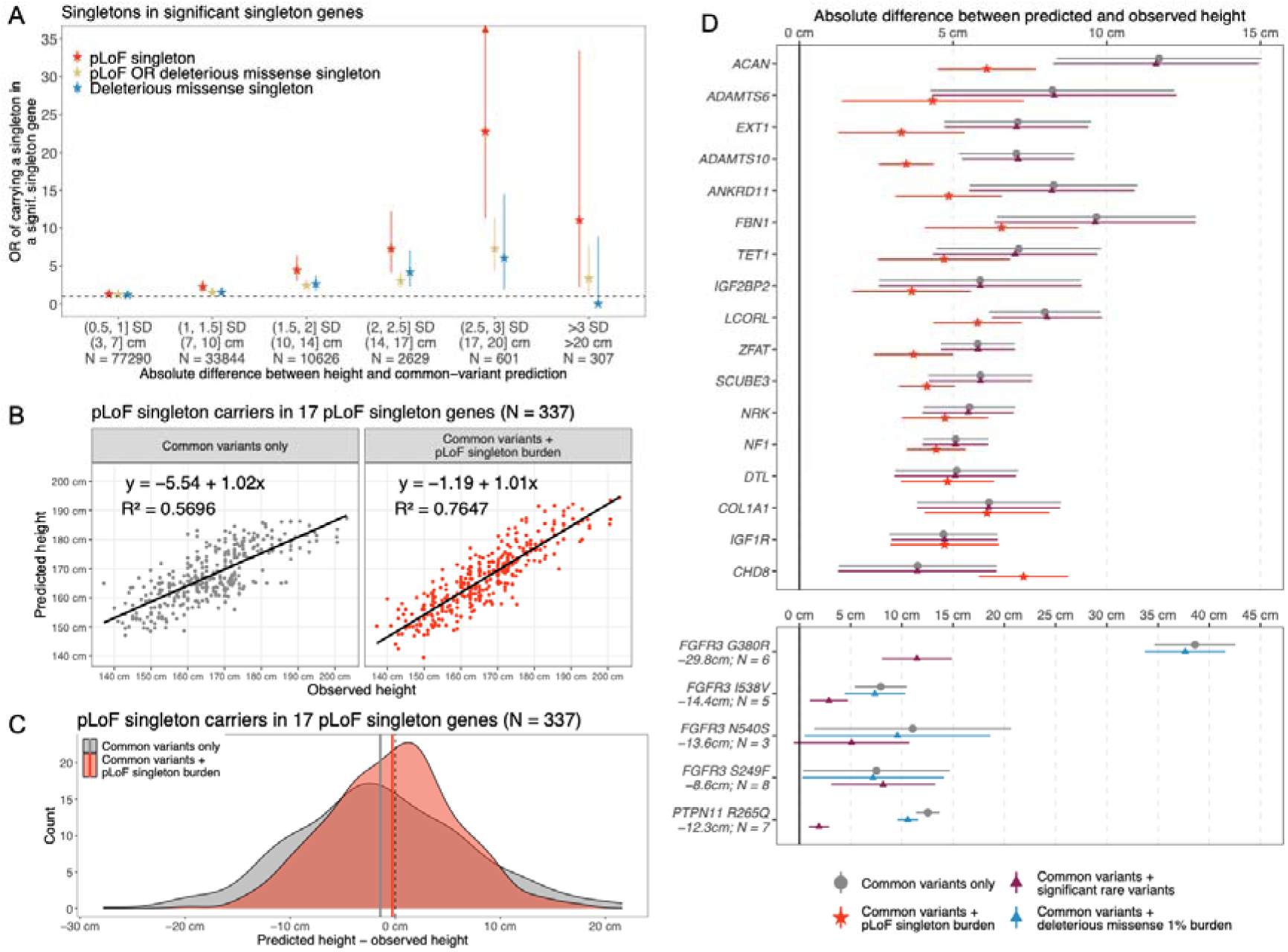
**A**) Enrichment (two-sided Fisher’s exact test; **Methods**; **Supplementary Table 18**) of individuals carrying singleton variants in genes from significant singleton burden tests (pLoFs: 17 genes; deleterious missense: eight genes; pLoF+deleterious missense: 17 genes) based on accuracy of common-variant prediction in standard deviations (SD). Each point represents enrichment of a given burden (color) among individuals with the given predictive error, relative to individuals with predictive error < 1 SD. Synonymous variants are not shown as we did not observe any significant height associations with burdens of singleton synonymous variants. These distributions separated by gene are provided in **Extended Data** Figure 6. Note: Because prediction was done separately by cohort and sex, the exact value of a standard deviation in centimeters varies; however, we provide weighted average values in centimeters across ten replication cohort-sex pairs for reference. On average, 1 SD = 6.71 cm in replication data. **B**) Predicted and observed heights among the 337 individuals that carry a pLoF singleton variant among 17 genes with pLoF singleton burdens that are significantly associated with height (**Table 1**). Values shown are with and without including a pLoF singleton burden PGS in a predictive model including covariates and common variants. Equations, R^2^ values, and regression lines are all from the univariate linear regression of predicted on observed height. **C**) Difference between predicted and observed height among the same 337 individuals and predictive models as **B**. Solid vertical lines represent means within each group (common variants only: mean = -1.4 cm, SD = 8.2 cm; common variants + pLoF singleton burden: mean = 0.3 cm, SD = 6.1 cm). **D**) Average predictive error (in cm) for carriers of pLoF singleton variants in the 17 pLoF singleton genes (top), and for carriers of significant deleterious missense variants in *FGFR3* and *PTPN11* (bottom). Each point represents predictive error among carriers of a given pLoF singleton burden or single variant. Full deleterious missense variant IDs are as follows: *FGFR3* p.G380R - 4:1804392:G:A; *FGFR3* p.I538V - 4:1805636:A:G; *FGFR3* p.N540S - 4:1805643:A:G; *FGFR3* p.S249F 4:1801841:C:T; *PTPN11* R265Q - 12:112472981:G:A. Each variant is labeled with the effect size (cm) in discovery data and the number of carriers in replication data (excluding *All of Us*). pLoF singleton burden effect sizes are shown in **Table 1** and the numbers of carriers in replication data are given in **Extended Data** Figure 6. Error bars represent 95% confidence intervals of the observed distributions in **A** and **D**, and arrows are included when they exceed the plotting area. All individuals shown in all panels are from replication cohorts (except *All of Us*; **Supplementary Table 1**) and all height prediction is done including covariates from GWAS (**Supplementary Table 24**; **Methods**).

We next focused on the 337 individuals in the replication data (excluding *All of Us*) carrying a pLoF singleton in one of the 17 genes with significant pLoF singleton burden (**Figure 1A**; **Supplementary Table 5**). In these individuals, adding burden effects to a common variant prediction model (**Methods**) improved predictive error by an average of 1.80 cm, increasing R^2^ from 0.57 to 0.76 (**Figure 3**; **Supplementary Note**). In carriers of *ACAN*, *ADAMTS6*, *EXT1*, and *ADAMTS10* pLoF singletons, however, mean improvement was >3.5 cm. Including pLoF singleton burden effects into prediction also reduced the standard deviation of the residual distribution by 2.14 cm, signifying more consistent predictions; however, improvement varied substantially by gene (**Figure 3C & D**; **Extended Data Figure 6**).

Finally, due to the large effect sizes of *FGFR3* and *PTPN11* rare missense variants (**Figure 2B**), we evaluated the performance of several rare variant based prediction models compared to standard common variant PGS in these variant carriers (**Figure 3D**; **Methods**). Among 29 carriers of five rare deleterious missense variants in these two genes in the replication dataset (excluding *All of Us*), the prediction improved ∼1.0 cm when using a PGS built from a deleterious missense (AAF < 1%) burden effect size. In contrast, prediction improved by an average of 9.5 cm when using a PGS based on individually significant rare variant effect sizes. This difference in prediction performance was especially pronounced among the six carriers of *FGFR3* p.G380R (4:1804392:G:A; β*_discovery_* = -29.8 cm; *P* = 3.2×10^-64^; **Figure 2B**), a ClinVar pathogenic variant reported in achondroplasia^7^ (**Figure 3D**), for whom height prediction improved by an average of 27.1 cm after accounting for *FGFR3* p.G380R. When substantial effect size heterogeneity exists among rare coding variants, as in *FGFR3* and *PTPN11* (**Supplementary Note**), aggregate burden effects are unsuitable for use in prediction. Still, individual rare variants are difficult to incorporate in prediction, as variants are frequently not observed in reference samples, and for those that are, effect sizes are often estimated from few observations. Overall, we observe that polygenic score models that supplement common variants with both aggregate rare variant effects and individual rare variants with extreme effects perform best for height (**Supplementary Figure 7**). We speculate that similar combination approaches will enable the best predictions for other traits as well.

### Comparison of rare variant results with ClinVar annotations

Finally, population-based data is a useful orthogonal resource to aid variant interpretation in clinical contexts^23^. We assessed whether our results could inform clinical interpretation by systematically comparing rare variants in our study with those in ClinVar. First, we evaluated whether our results could inform on the pathogenicity of ClinVar variants with conflicting (CCOP) or insufficient evidence (VUS). To maximize power for these ultra-rare variants we performed a meta-analysis of our discovery and replication data (**Supplementary Tables 20** & **21**; **Extended Data Figure 7**) for the 5,233 rare variants in our 207 height-associated genes and evaluated in ClinVar for links to stature-related syndromes (**Methods**). Of these 5,233 rare variants, 39 were significant: two benign/likely benign, 17 pathogenic/likely pathogenic (P/LP), and 20 VUS/CCOP. Of the 20 significant VUS/CCOP variants, only one – a splice region variant near *ADAMTS10* – was associated with increased height (*β*= +2.3 cm; *P =* 3.9×10^-15^), the opposite direction of the rare variant burden tests (**Supplementary Table 6**). All three categories contained more significant height-associated variants than expected by chance (*P* = 6×10^-120^ for P/LP, binomial test; **Supplementary Table 22**).

We next looked for variants absent from ClinVar with absolute effect sizes greater than 7.99 cm (1 SD), an effect size greater than 77% of the stature-related P/LP variants in the 207 genes (**Figure 4**; mean and median absolute effect: 5.2 cm and 4.3 cm, respectively). At this effect size threshold, four significant variants were completely absent from ClinVar as of July 2025 (**Supplementary Table 21**): pLoFs in *HMGA2* (p.E98*), *LCORL* (p.S1370Rfs*10), and *ADAMTS17* (p.S971*), and a missense substitution in *ZFAT* (p.R762C).

**Figure 4:**
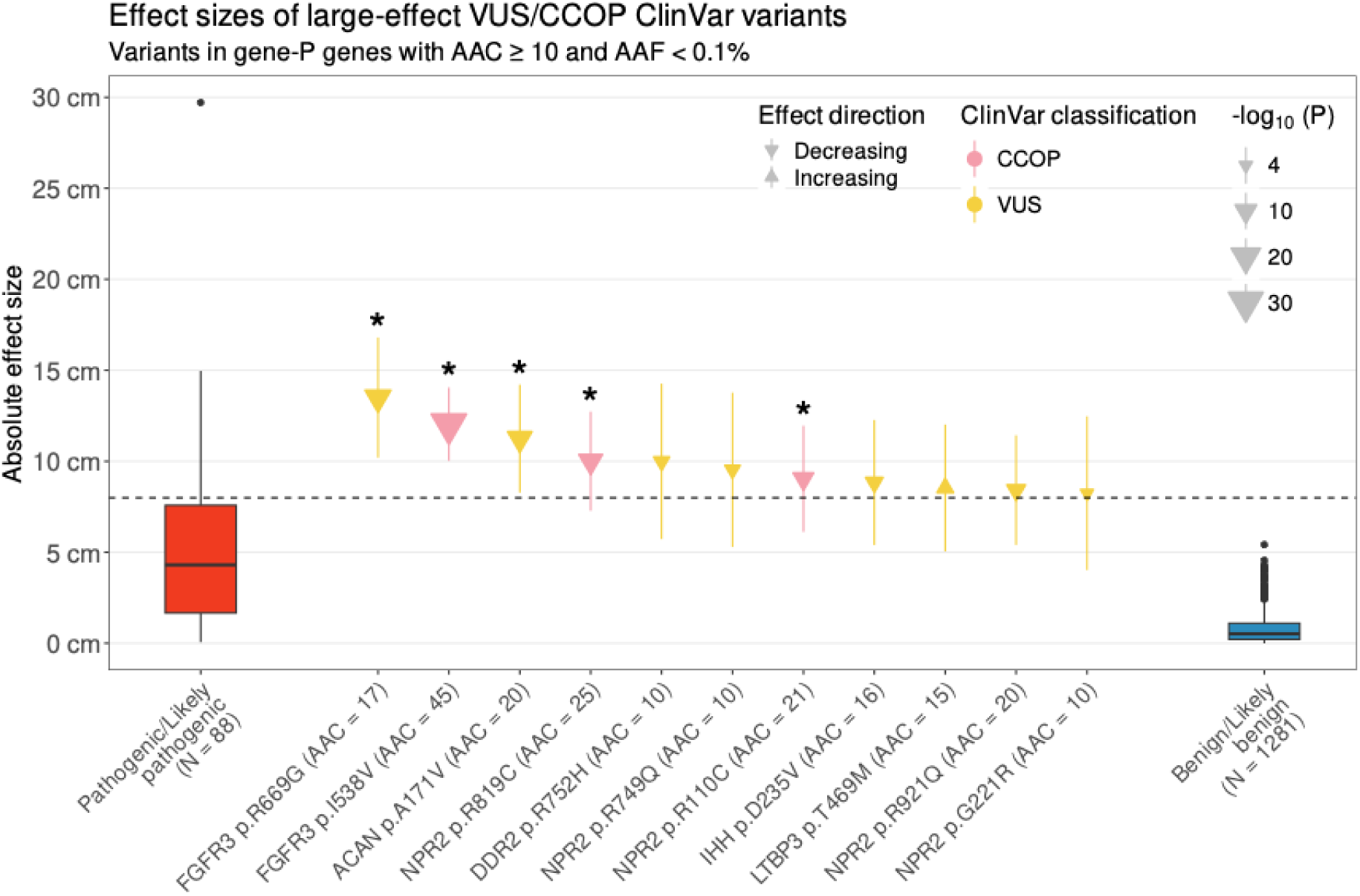
Effect sizes of large-effect VUS and CCOP ClinVar variants compared to ClinVar variants with other classifications. All variants shown have AAC≥10, AAF < 0.1%, and impact one of the 207 genes identified in the discovery analysis. VUS/CCOP variants (in yellow) also have effect estimates of at least 7.99 cm (1 SD – indicated by the horizontal dashed line). Asterisks indicate variants which were genome-wide significant (*P* < 1.75×10^-9^). Effect estimates and *P*-values were estimated from the meta-analysis of discovery and replication data (**Supplementary Table 20**). The outlier red variant near 30 cm is *FGFR3* p.G380R, a ClinVar pathogenic variant associated with achondroplasia (4:1804392:G:A; **Supplementary Table 3**). VUS – variant of uncertain significance; CCOP – conflicting classifications of pathogenicity; AAC – alternate allele count; AAF – alternate allele frequency.

Eleven VUS/CCOP variants with AAC≥10 had absolute effect sizes greater than 7.99 cm, with P values ranging from 1.6×10^-4^ to 1.1×10^-31^ (five are significant with P < 1.75×10^-9;^ **Figure 4**; **Supplementary Table 20**). Of note are three missense variants in *NPR2* – p.G221R, p.R749Q, and p.R921Q – classified as VUS in ClinVar but large effects on height (between -8.24 and -9.53 cm). Although none of these missense variants achieves genome-wide significance in our study, p.G221R and p.R921Q have been experimentally validated as LoF^46^. We present a limited set of large-effect variants that are either not annotated for clinical effects on height or have uncertain (VUS/CCOP) annotations, that serve as examples of how population-scale rare variant association analyses provide valuable data relevant to clinical interpretation that can be added to the clinical geneticist’s toolkit for evaluating disease-causing variants.

## Discussion

We undertook an exome-wide association study in 1,450,633 individuals, identifying 207 genes associated with height after conditioning on common variants, 115 more gene associations than an earlier large scale sequencing study^9^. In 17 genes, the rarest and most deleterious class of variation, singleton pLoFs, were associated with absolute effects on height 52× larger than those of the average common GWAS variant (8.92 cm compared to 0.17 cm). Though we noted a substantial overlap with Mendelian syndromes, less than a quarter (44/207) of identified genes include a stature or skeletal growth component in their OMIM description^2^. Moreover, there was even less overlap for height-increasing genes, suggesting that many height-increasing genetic syndromes or syndromic features may be overlooked. Collectively, large-effect rare variant associations show strong concordance with genes implicated by common variant associations and reflect established biological processes influencing height including growth pathways, extracellular matrix organization, and transcriptional regulation.

Our results illustrate classic trade-offs in genetic association studies between effect size and allele frequency. Expanding our analysis beyond singletons to include low frequency variants up to 1% increased the number of implicated genes, but at the cost of diminished effect sizes. Similarly, expanding from pLoFs to also include missense variants yielded the most associated genes, but effect sizes decreased further. Together, these increased the number of implicated genes from 17 to 179 but greatly reduced the average absolute effect size. Tests that more explicitly allow for heterogeneity in effect size^12^ or small proportions of causal variants^11^ within each gene increase gene discovery another 16%, bringing the total to 207 genes. Overall, no single analysis captured all implicated genes. We maximized gene discovery by combining gene-based *P*-values from multiple models with a well-calibrated omnibus gene-based test^10^. The single most powerful analysis included the combination of pLoF and damaging missense variants with frequency <1%, identifying 108 genes. Our results emphasize the value of combining association analyses at various allele frequency thresholds and levels of variant deleteriousness: whereas singleton pLoFs identified large genetic effects in 17 genes, extending the analysis to less rare pLoFs and including deleterious missense variants increased the number of associations nearly 10-fold and identified some genes with effect sizes rivaling that of singleton pLoFs (e.g., *SHOX*). In contrast, while we did find examples where it was important to model heterogeneity in effect sizes within a gene (e.g., *PTCH1*), burden tests discovered most genes.

Of the 17 genes with significant singleton pLoF burden signals, a majority (8/11) of height-decreasing genes have a documented stature or skeletal growth-related Mendelian syndrome in OMIM; however, this is only true for one of the six height-increasing pLoF singleton burdens (*FBN1*), raising the question whether increased height is under-reported in OMIM as a phenotypic characteristic of Mendelian syndromes or if genes whose perturbation lead to increased height are less likely to have other phenotypic consequences. In some genes, deleterious variation may simply be too rare to have been described. For example, *TET1* is a highly constrained gene in which singleton pLoFs are associated with increased height (+8.32 cm). Even across all 1.4 million individuals in our dataset, we only observe 90 carriers of pLoF singletons in this gene, emphasizing the strong evolutionary constraint in this gene (pLI = 1). Other genes with height-increasing singleton pLoFs do have Mendelian diseases described in OMIM but lack a stature or skeletal growth component. For example, while singleton pLoF variation in *ZFAT* is associated with a 7.86 cm height increase, no stature or skeletal growth syndrome is explicitly noted in OMIM. While this pattern appears consistent in our results, we do note that for consistency and ease of comparison, we used the OMIM list compiled by Yengo *et al*. (2022), which derived height and skeletal growth genes from late-2019 to mid-2020, so this observation may reflect the logistical challenge of keeping a single database up to date with the extensive literature as well as updates since 2020.

The 207 genes implicated in height include those in relatively well-understood pathways but also implicate potentially new areas of height biology. For example, in the *GH/IGF* pathway, growth hormone (*GH1*) signals for production of IGF1, which stimulates skeletal growth via cartilage generation in the growth plates^47^. While Mendelian syndromes with altered stature as a major feature are linked to eight members of the *GH/IGF* pathway: *GH1, GHR, GHRHR, GHSR, IGF1, IGF1R, IGFLAS, PAPPA2*^48^, our results include not only seven of these eight (**Supplementary Table 23**), but further implicate three other pathway genes (*PAPPA*, *STC2*, and *GHRH*). Furthermore, for several genes previously considered to have autosomal recessive modes of inheritance (*GHRHR, IGFALS, IGF1*, and *PAPPA2*), our results show that loss-of-function and deleterious missense variants have apparent effects in the heterozygous state (**Supplementary Table 23**). Finally, while GH1/IGF1 is generally considered the major driver of post-natal growth and development, the IGF2 pathway is believed to act mainly during pre-natal development with uncertain effects on adult height^49^. Human genetic evidence for a role of the IGF2 pathway in height is limited to extreme short stature (>3 SDs below average) in Silver-Russell syndrome, caused by disruptions in the *IGF2* gene itself (but not implicated here)^50^. Our results include association in an additional *IGF2* pathway gene: *IGF2BP2* via singleton pLoFs (*β* = -8.67 cm; *P =* 1.52×10^-10^; **Table 1**), an mRNA binding protein that stabilizes *IGF2* mRNA, further evidence of a role of developmental IGF signaling influencing adult height.

Accurate prediction of complex traits is a key aspiration of human genetics. Current height prediction efforts focus on using several thousand common variants with effects that are individually small, but large in aggregate; we supplement this approach by adding rare variants with individually large effects. While we expect many more rare variants will be implicated in the future, those discovered here demonstrate substantial added value towards accurately predicting height for those who carry them. Our replication data show that, for 337 singleton pLoF carriers across our 17 genes with height-associated singleton pLoF burden tests in the discovery dataset, common variant prediction alone underperforms – with an average absolute error of 6.62 cm (as opposed to 4.17 cm across the population). The effect was even more pronounced for carriers of large-effect rare missense variants in *FGFR3* and *PTPN11*, whose height predictions were off by ∼15 cm when predicted using common variants alone.

*FGFR3* and *PTPN11* frequently arose as interesting case studies throughout this work. First, they represent an unusual case where rare missense variants collectively yielded a significant burden association, but where pLoFs did not contribute to the signal. They are also known to reduce height via gain-of-function mechanisms^28^ and implicated in achondroplasia^7^ and Noonan syndrome^29^, respectively. However, burden test signals were relatively muted (3-15× smaller; - 2.31 cm for *FGFR3*, -4.12 cm for *PTPN11*) in these genes compared to the much larger effects of individual variants: a -29.8 cm effect for p.G380R in *FGFR3* and -14.2 cm for p.N308D and – 12.2 cm for p.R265Q in *PTPN11*). For most other genes, effect sizes were more consistent for missense and pLoFs and the most statistically powerful analysis combined missense variants with pLoFs.

Collectively, this work dramatically expands the breadth of rare genetic associations for adult height. Beyond directly implicating 207 genes, we bridge the gap between rare Mendelian and population-scale quantitative genetics and provide valuable data for interpreting clinically observed variation in patients with abnormal stature. Our analytic approach provides an unparalleled guide for causal gene identification and the interpretation of functional mechanisms at hundreds of GWAS loci. Just as the early common variant height studies informed subsequent GWAS of other diseases^2^, we hope the study design, methodological considerations, and conclusions presented here will inform the rare variant studies of other traits and diseases.

## Acknowledgements

This research has been conducted using the UK Biobank Resource (Project 26041). We gratefully acknowledge *All of Us* participants for their contributions, without whom this research would not have been possible. We also thank the National Institutes of Health’s *All of Us* Research Program for making available the participant data examined in this study. The Penn Medicine BioBank is funded by a gift from the Smilow family, the National Center for Advancing Translational Sciences of the National Institutes of Health under CTSA Award Number UL1TR001878, and the Perelman School of Medicine at the University of Pennsylvania. The Mexico City Prospective Study received funding from the Mexican Health Ministry, the Mexican National Council of Science and Technology, the Wellcome Trust (058299/Z/99), the British Heart Foundation, Cancer Research UK, Kidney Research UK, the UK Medical Research Council (MC_UU_00017/2 and MR/Z504543/1), and the Nuffield Department of Population Health at the University of Oxford. Genotyping, exome sequencing and whole-genome sequencing were funded through an academic partnership between the UNAM, the University of Oxford, Regeneron, AstraZeneca, and AbbVie. The Malmö Diet and Cancer study was funded by grants from the Swedish Medical Research Council, the Swedish Cancer Foundation, the Albert Påhlsson and Gunnar Nilsson Foundations, AFA insurance, and the Malmö city council. The University of California Los Angeles (UCLA) ATLAS Collaboration is supported by the David Geffen School of Medicine, UCLA Health, and the UCLA Clinical and Translational Science Institute (UL1TR001881). We thank everyone who made this work possible, the professionals from the member institutions who contributed to and supported this work, and especially all research participants, without whom this research would not be possible. Regeneron Pharmaceuticals and the Regeneron Genetics Center funded this study.

## Author contributions

J.A.K., L.G., and A.E.L. performed the analyses. J.A.K., L.G., G.R.A., M.A.R.F., A.E.L. wrote the manuscript. J.A.K. and L.G. created the figures. G.R.A., M.A.R.F., and A.E.L. jointly supervised the work. T.J. performed the fine-scale ancestry analysis. S.M.G. performed follow-up population genetic analyses. K.Y.S., C.Z., and S.B. provided ClinVar data and annotations. A.I.C. wrote the pipeline for meta-analysis of interactions. K.W. implemented the iterative conditional analysis pipeline. A.Z., J. Mbatchou, and J. Marchini implemented gene-P. J.A.K., L.G., M.D.K., D.B., J. Bovijn, N.V., L.A.L., and A.E.L. contributed to phenotypic data curation and biological interpretation of results. J.D.B. ran the SVM QC and provided UK Biobank burden test results. A.W.C., O.M., R.W., J.T., P.K.M., R.T., J.A., J. Berumen, L.A.L., and A.B. provided data. All authors reviewed and approved the final version of the manuscript.

---

**Extended Data Figure 1.**
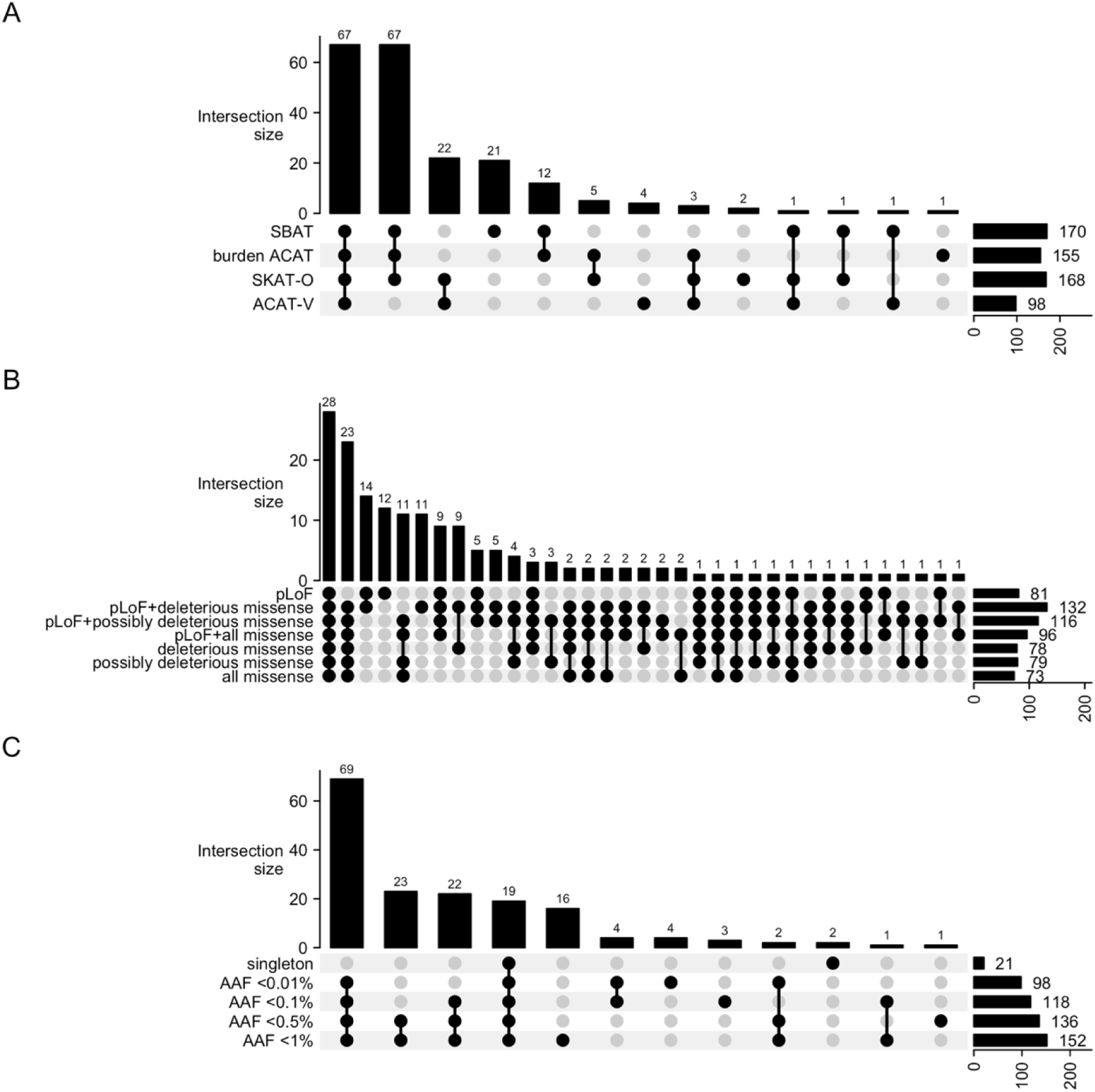
The contribution of (**A**) gene-based models, (**B**) variant class, and (**C**) AAF thresholds towards gene discovery. (**A**) UpSet plot decomposing the 207 genes identified by gene-P into the four underlying gene-based test methods: SBAT, burden ACAT, SKAT-O, or ACAT-V. The union of SBAT and burden ACAT yielded 179 genes from which we considered as our set of burden genes. UpSet plots for the 179 burden genes broken down by (**B**) variant class and (**C**) AAF threshold.

**Extended Data Figure 2:**
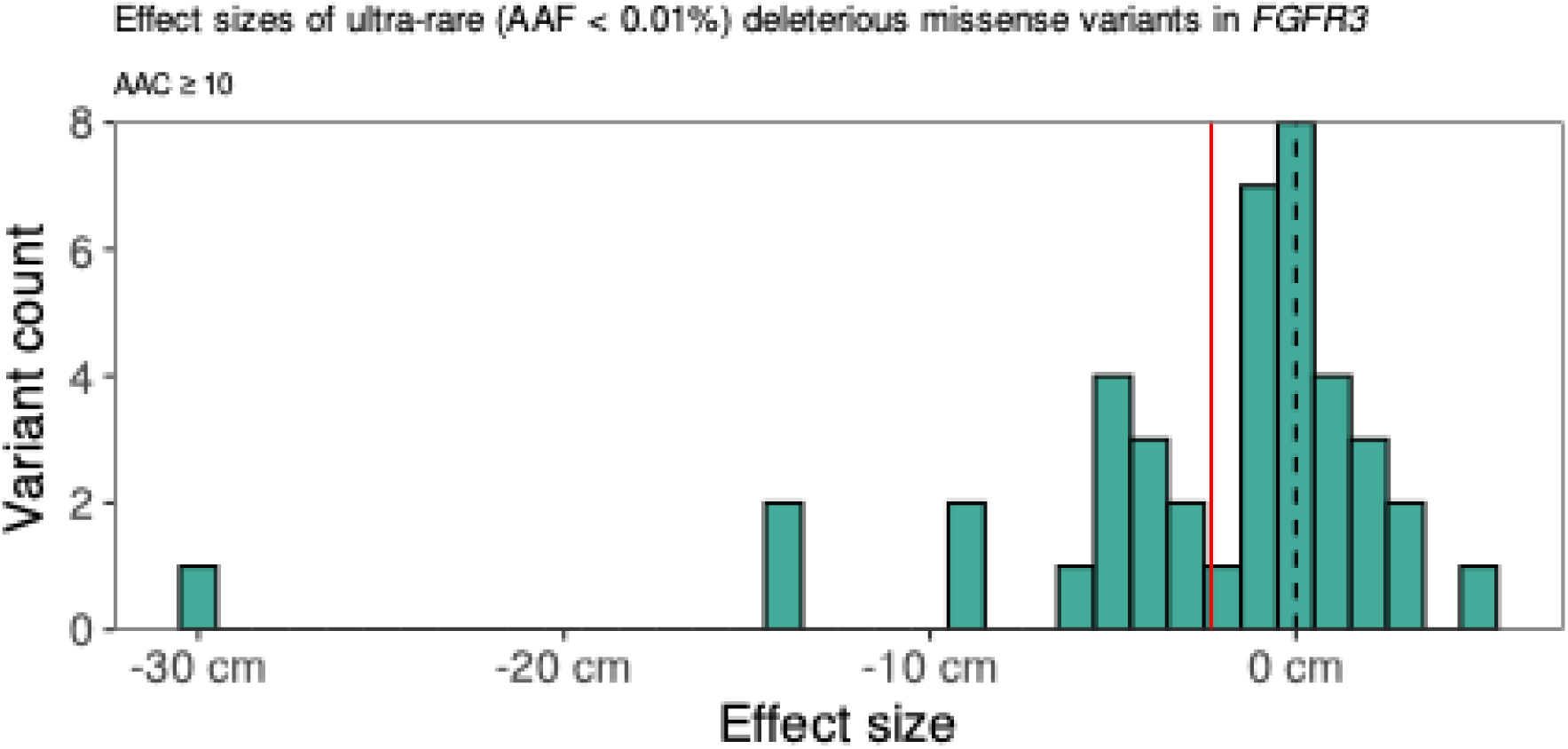
Effect size distribution of ultra-rare (AAF < 0.01%) deleterious missense variants in *FGFR3*. The dashed black line indicates no effect (0 cm), and the solid red line indicates the burden effect size when aggregating all the variants in the figure (β = -3.1 cm).

**Extended Data Figure 3:**
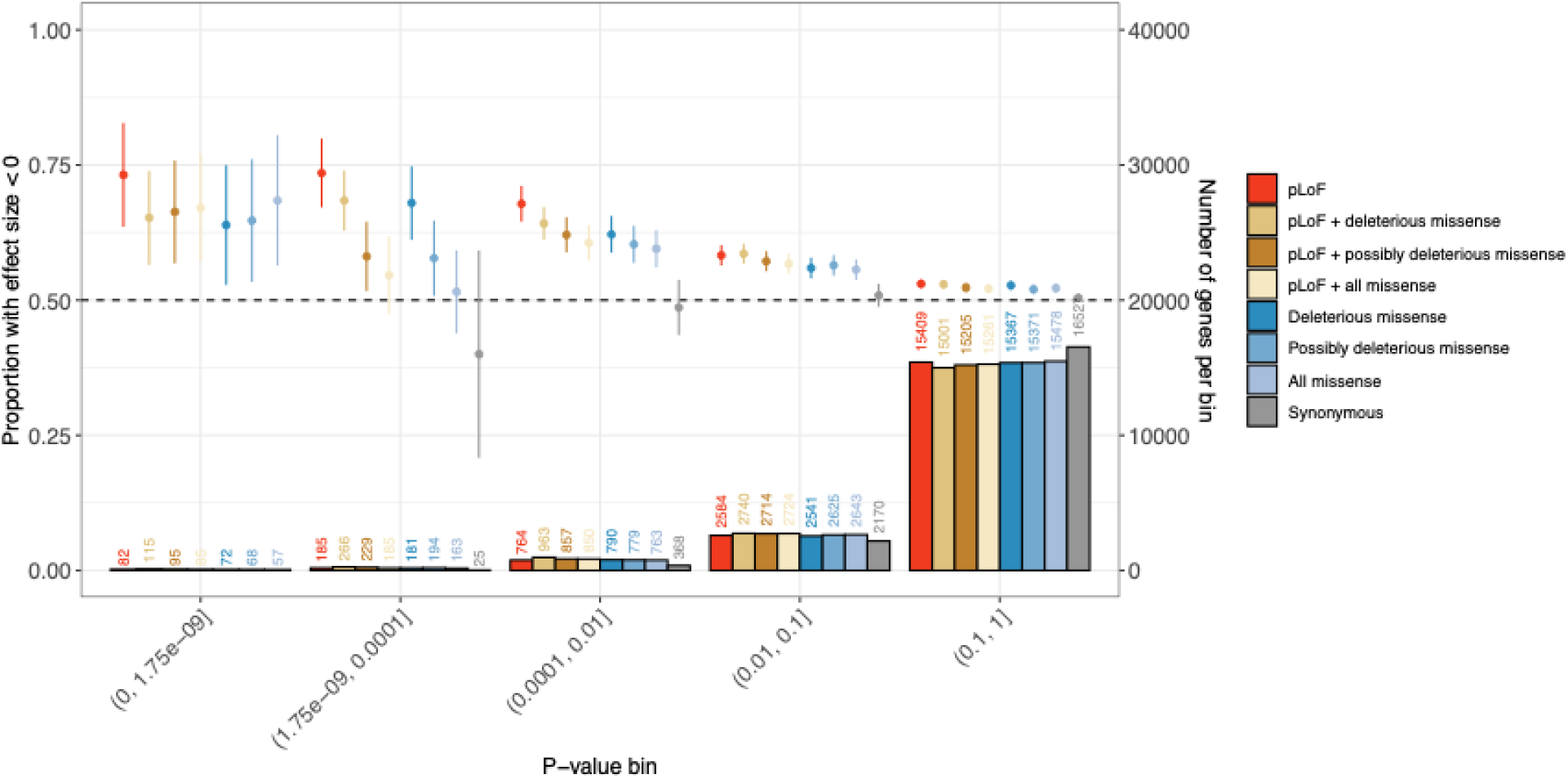
Proportion of AAF < 1% burden effects that are negative. Each point represents a group of burdens of a given type (specified by color) that meet a given *P*-value threshold. Proportion of negative synonymous burdens with P < 1.75×10^-9^ (n<5) not shown. Error bars represent 95% confidence intervals around the point estimate. Vertical bars represent the number of genes in each bin.

**Extended Data Figure 4:**
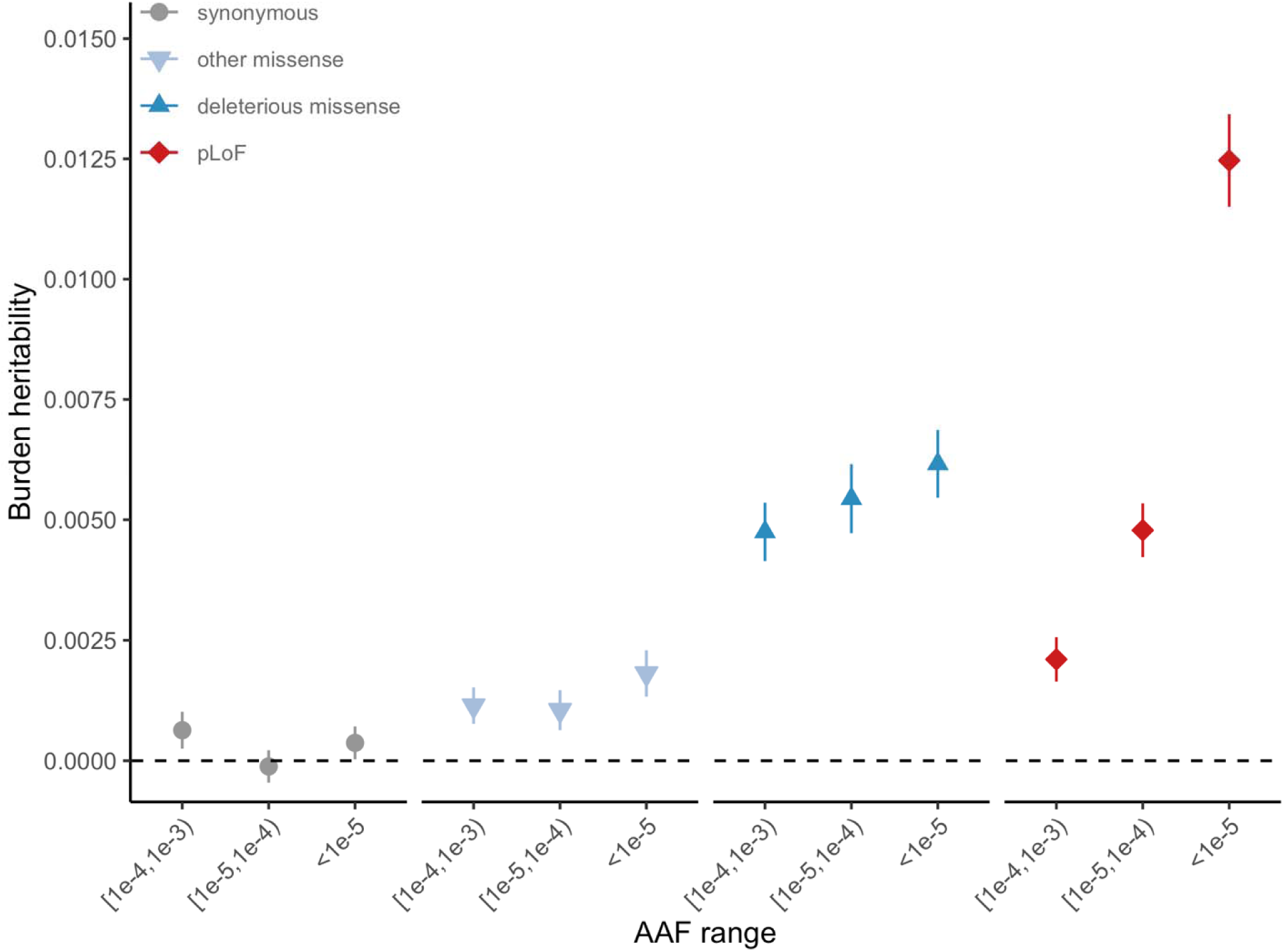
Estimated heritability of the burden of rare coding variants partitioned by alternate allele frequency (AAF) bins and annotation. Estimated using burden heritability regression (BHR). Error bars represent 95% confidence intervals around the point estimate.

**Extended Data Figure 5:**
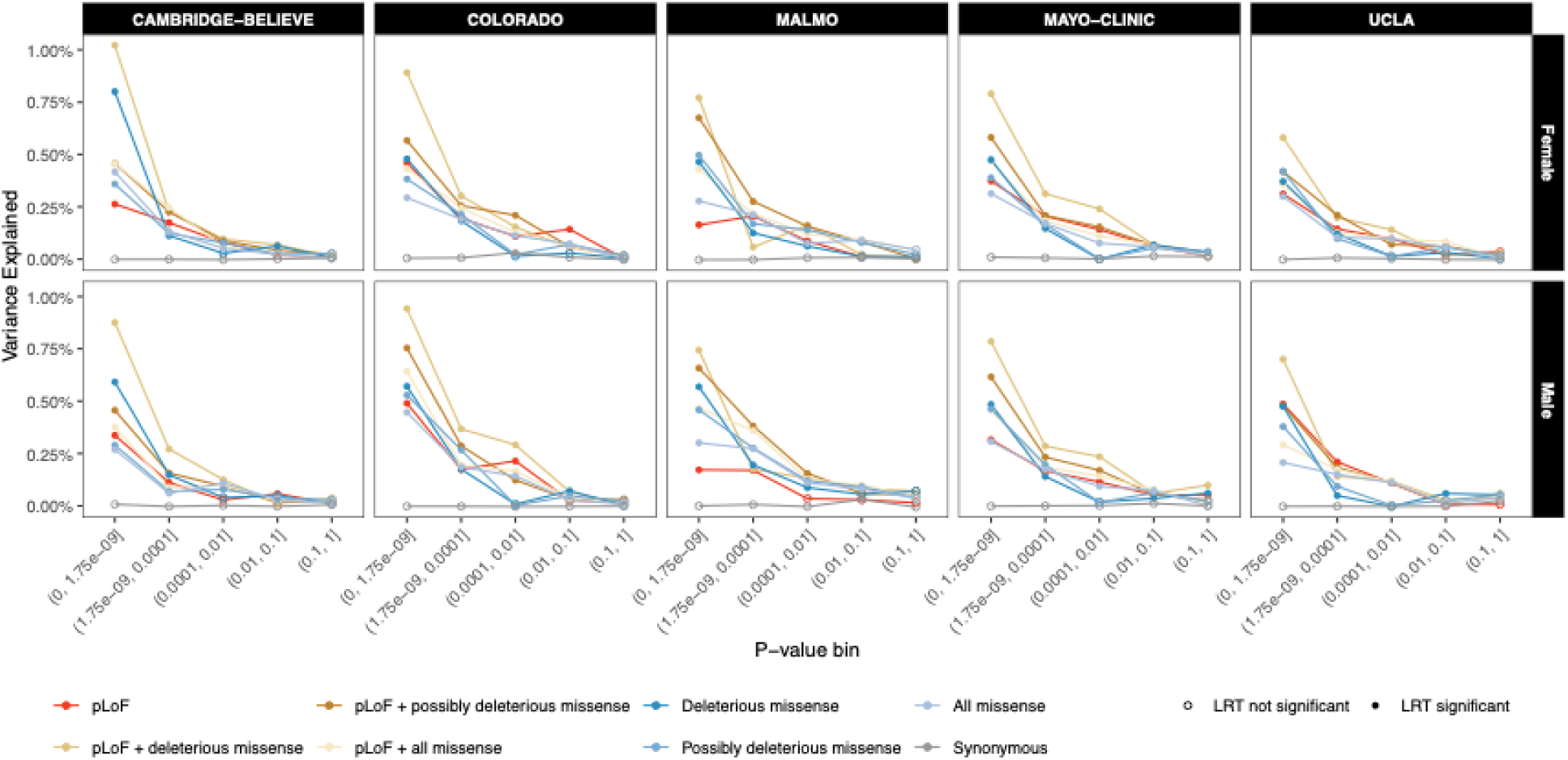
Variance explained in replication data by burden tests with AAF < 1%. Variance explained is calculated as the difference in adjusted R^2^ between nested linear models of height: one with only common variants and covariates and one that also includes a rare variant PGS. Each point represents all burdens of a given type (specified by color) that meet the given *P*-value threshold (the number of burdens in each point is given in **Extended Data** Figure 3). Likelihood Ratio Test (LRT) used to determine whether the variance explained by rare variant burdens was significantly different from zero at a conservative, Bonferroni-corrected 11 = 0.0001 to adjust for 500 tests performed.

**Extended Data Figure 6:**
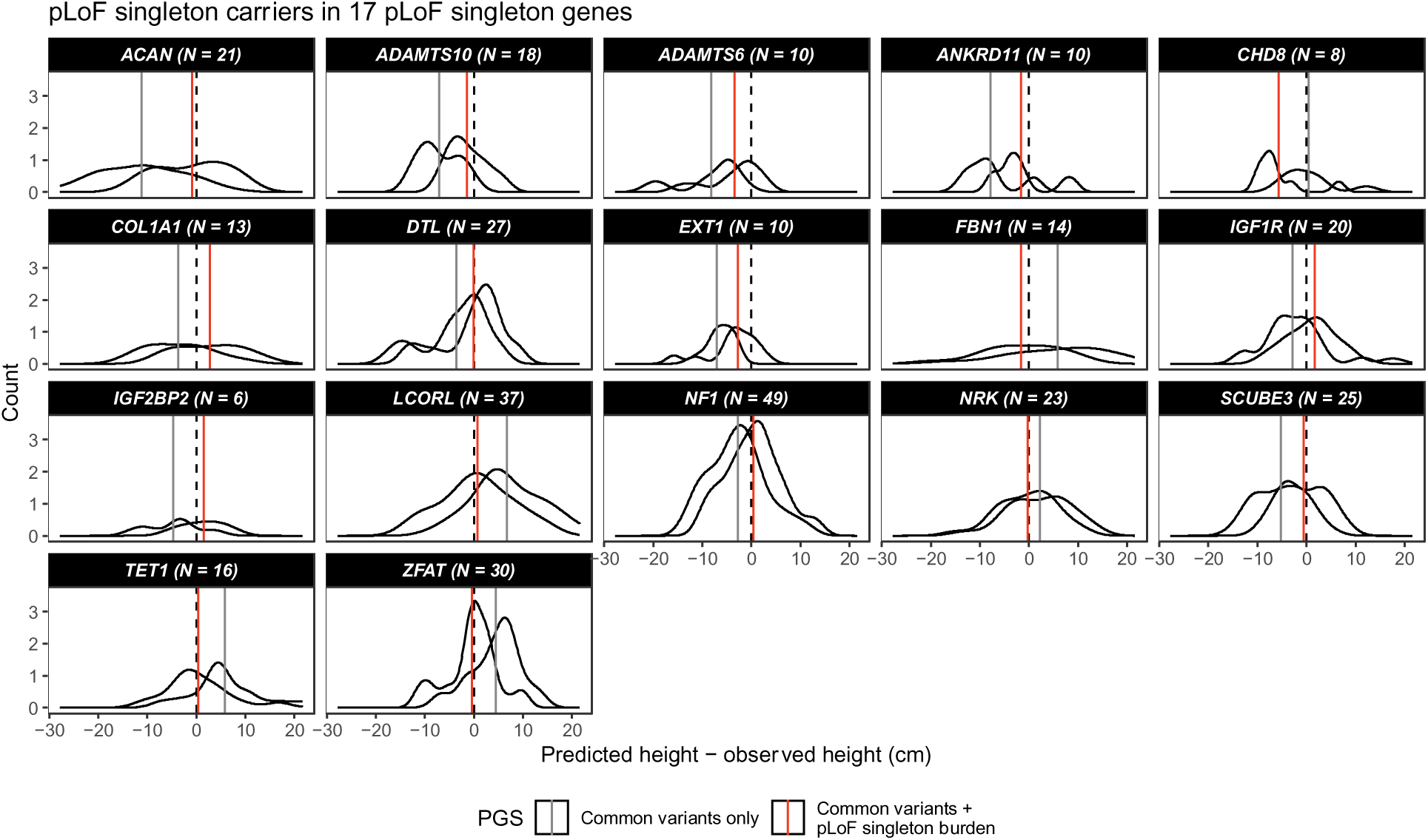
Accuracy of height prediction for carriers of pLoF singletons in 17 genes with a pLoF singleton burden significantly associated with height (**Table 1**). Solid vertical lines represent mean values for each prediction method; black dashed line indicates difference between observed and predicted height of zero. All individuals shown are in replication data.

**Extended Data Figure 7:**
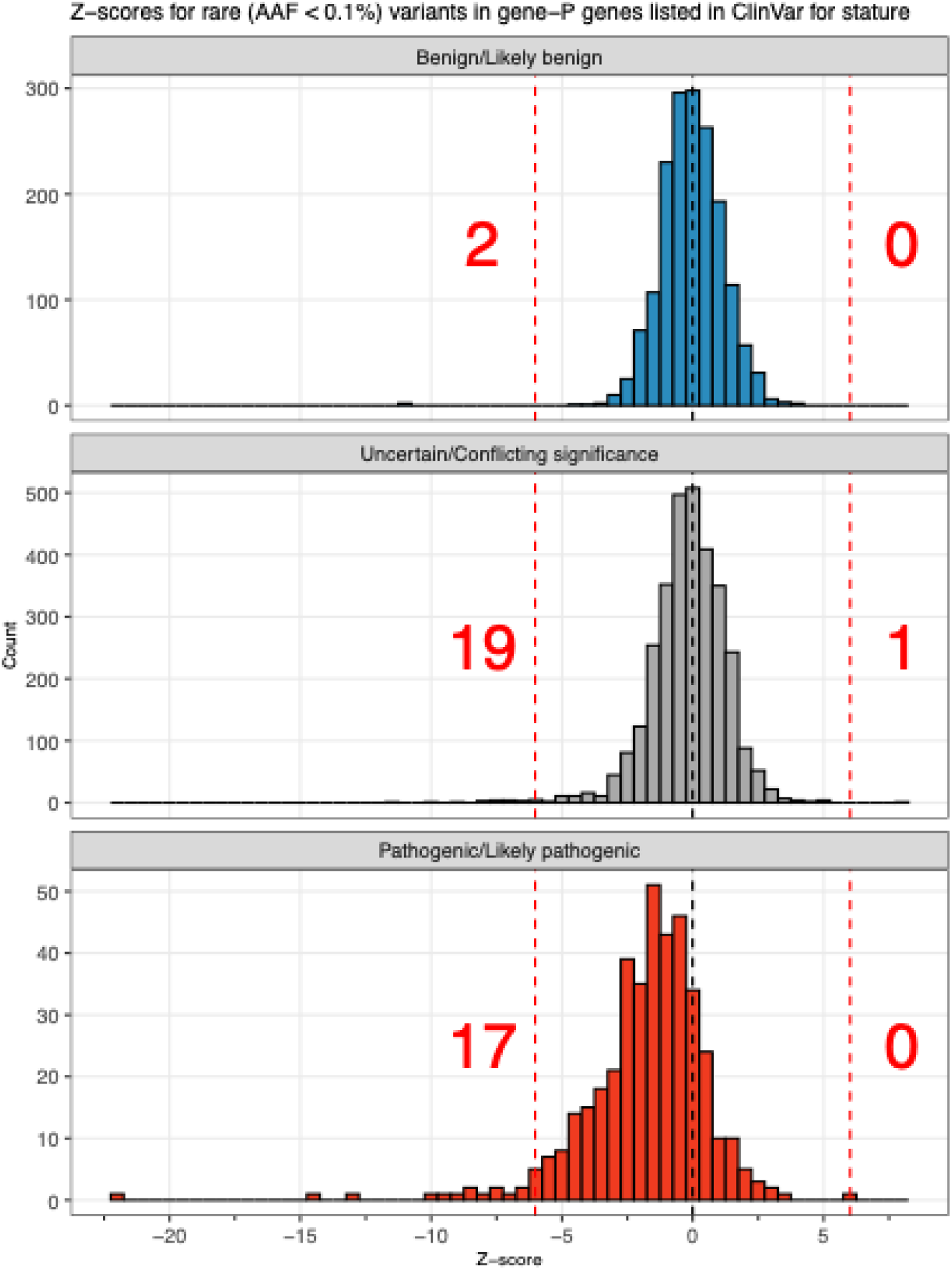
Z-scores for rare (AAF < 0.1%) variants in a gene-P gene and listed in ClinVar for a stature-related condition, separated by ClinVar pathogenicity. Black dashed lines indicate a Z-score of 0 and red dashed lines indicate the significance threshold corresponding to *P=*1.75×10^-9^ (Z = ±6.02). Red numbers represent the number of variants with Z scores more extreme than this threshold.

## Methods

### Ethics Statement

Ethical approval for the UK Biobank was previously obtained from the North West Centre for Research Ethics Committee (11/NW/0382). The work described herein was approved by UK Biobank under application number 26041. The Geisinger Health System Institutional Review Board (IRB; #2006-0258) approved all DiscovEHR analyses. For the University of Pennsylvania PennMedicine BioBank, appropriate consent was obtained from each participant regarding storage of biological specimens, genetic sequencing and genotyping, and access to all available electronic health record (EHR) data. This study was approved by the University of Pennsylvania IRB and complied with the principles set out in the Declaration of Helsinki. The Icahn School of Medicine at Mount Sinai’s IRB approved the BioMe cohort. The Mexican Ministry of Health, the Mexican National Council for Science and Technology, and the University of Oxford approved the MCPS study. For Mayo-RGC Project Generation, all subjects provided informed consent for use of specimens and data in genetic and health research and ethical approval and consent for Project Generation was provided by the Mayo Clinic IRB (#09-007763). Ethical approval and consent for the Colorado Center for Precision Medicine (CCPM) Biobank was reviewed and approved by the Colorado Multiple IRB (#15-0461). Patient recruitment and sample collection for UCLA’s Precision Health Activities was approved by the UCLA IRB (#17-001013). Lund University’s Regional Ethics Committee approved the Malmö Diet and Cancer Study (MDCS). ADAGES III approval and consent was provided by each institution’s IRB. The *All of Us* IRB follows the regulations and guidance of the NIH Office for Human Research Protections for all studies, ensuring that the rights and welfare of research participants are overseen and protected uniformly.

### Discovery cohort summaries

Our discovery analysis comprised six exome-sequenced cohorts jointly called on an IDT-exome capture as part of the RGC-ME study^1^^33^.

#### African Descent and Glaucoma Evaluation Study (ADAGES) III^134^

ADAGES III was designed to study glaucoma and recruited individuals (ages 20-96) with glaucoma, glaucoma progressors, and controls for two years (August 2014 – 2016) from five centers in Alabama, California, Georgia, New York, and Texas. All participants provided written informed consent and provided blood or saliva for DNA extraction.

#### BioMe

Charles Bronfman Institute for Personalized Medicine at the Mount Sinai Medical Center established the BioMe Biobank Program in 2007. BioMe participants comprise patients (ages 18-89) enrolled in the Mount Sinai health system who consented to participate and gave EHR access.

#### Geisinger Health System (GHS) DiscovEHR study

The GHS MyCode Community Health Initiative is a health system-based cohort from central and eastern Pennsylvania (USA) with ongoing recruitment since 2006^135^. DNA from participants was genotyped on either the Illumina OmniExpress Exome (OMNI) or Global Screening Array (GSA) and imputed to the TOPMed reference panel (stratified by array) using the TOPMed Imputation Server. Prior to imputation, we retained variants that had a MAF < 0.1%, missingness < 1% and HWE *P*-value > 10^-15^. Following imputation, data from the OMNI and GSA datasets were merged for subsequent association analyses, which included an OMNI/GSA batch covariate, in addition to other covariates described below.

#### Mexico City Prospective Study (MCPS)^39^

The MCPS is a prospective cohort comprising 159,755 adults (at least 35 years of age) recruited between 1998-2004 from the urban Mexico City districts of Iztapalapa and Coyoacán. All participants provided blood samples and signed consent forms for their data to be used for research purposes including analyses of genetic factors. DNA genotyping was performed using Illumina’s GSA and imputed to an MCPS reference panel from whole genome sequencing.

#### Penn Medicine BioBank (PMBB) study

PMBB contains ∼70,000 study participants (ages 19-90), all recruited through the University of Pennsylvania Health System (UPHS). Participants donated blood or tissue and allowed access to their EHR information^136^. DNA genotyping was performed with the Illumina GSA, and imputation performed using the TOPMed reference panel.

#### UK Biobank (UKB) study

The UK Biobank study includes approximately 500,000 adults aged 40-69 at recruitment during the period between 2006 and 2010. DNA samples were genotyped using the Applied Biosystems UK BiLEVE Axiom Array (N=49,950) or the closely related Applied Biosystems UK Biobank Axiom Array (N=438,427)^137^. Genotype data for variants not included in the arrays were inferred using the TOPMed reference panel, as described above.

### Replication cohort summaries

We performed a meta-analysis across six replication cohorts not part of the RGC-ME study that were either 1) exome sequenced using a TWIST capture (BELIEVE Study, The Biobank at Colorado Center for Personalized Medicine, Mayo Clinic Project Generation, UCLA ATLAS), 2) whole genome sequenced (*All of Us*), or 3) not able to be jointly called as part of RGC-ME due to regulatory restrictions (Malmö Diet & Cancer Study).

#### All of Us (AoU)

The National Institutes of Health’s AoU research program is a longitudinal cohort study aiming to include 1 million diverse participants across the United States of America, combining phenotypic data from various sources, including electronic health records^138^. Three sequencing centers performed the whole genome sequencing using DNA from both blood and saliva which we used batch covariates to correct for this in the analysis (**Supplementary Figure 8**). Unlike all other cohorts included in this study, participant-level phenotypic and genetic data cannot be removed from the trusted research environment and thus, we did not use AoU for height prediction analyses. All enrolled individuals provided informed consent.

#### BELIEVE Study^139^

Participants were recruited by approaching household heads of all eligible households with written and verbal information on the study. After the household head provided written consent, a structured questionnaire was administered. All participants (>10 years), who had resided in their current household for at least three years and intended to reside for an additional five years and provided both written informed consent (legal guardians in the case of non-adults) and prospective follow-up information, were eligible for recruitment.

#### The Biobank at Colorado Center for Personalized Medicine (CCPM)^140^

The CCPM biobank at the University of Colorado Anschutz Medical Campus in Aurora has enrolled >275,000 individuals. Participants consent to their biological material and medical records being used for research purposes. De-identified phenotype data was collected by Health Data Compass and comprises an individual’s entire medical record from the University of Colorado’s EHR.

#### Malmö Diet and Cancer Study (MDCS)

A population-based, prospective study from Malmö, Sweden that enrolled and consented 30,447 individuals (ages 44-73) between March 1991 and September 1996. Participants underwent a physical examination and provided information including lifestyle and clinical factors via a questionnaire.

#### Project Generation Mayo Clinic Biobank

Project Generation included 116,277 subjects (ages 18-89) from the Mayo Clinic Biobank^141^ (ongoing enrollment since 2009) and 30 disease-specific registries. Structured data in the Mayo EHR were extracted using the OMOP common data model (https://www.ohdsi.org/data-standardization/), and then de-identified with dates shifted by a random number of days to prevent re-identification.

#### University of California Los Angeles (UCLA) ATLAS Precision Health Biobank

The ATLAS Precision Health Biobank at UCLA comprises >100,000 individuals (ages 18-91) who consented for research. De-identified phenotype data comprises all hospital visits beginning in 2013 and converted to ICD-10 codes. All individuals provided written informed consent to the original recruitment of the UCLA ATLAS Community Health Initiative. All research performed in this study conformed with the principles of the Helsinki Declaration and used de-identified data (without any Protected Health Information data) with no possibility of re-identifying any of the participants. The data satisfies the HIPAA de-identification standard, and dates are shifted by a random number of days to prevent re-identification.

### Height definition

An individual’s standing height was calculated by first converting all height measurements to centimeters (cm) and removing any measurements taken before 18 years of age or <75 cm. From an individual’s remaining height measurements, we calculated their median height. Lastly, we applied a rank inverse normal transformation (RINT) separately for each sex and used this RINTed height phenotype in GWAS and exome analyses. We excluded individuals for whom their genetically predicted sex differed from their self-reported sex as well as 76 bone marrow transplant recipients for whom their DNA was collected from blood (as the DNA corresponds to the donor, but the phenotypes correspond to the recipient). To approximate height effect sizes in centimeters, we multiplied effect sizes (in SD units) by 7.994 cm, the sex-stratified, sample size weighted mean standard deviation across participants in the discovery dataset.

### Sample preparation and exome sequencing

As previously described, sample preparation and sequencing Genomic DNA libraries were created by enzymatically shearing high molecular weight genomic DNA to a 200-base pair (bp) mean fragment size^133^. Multiplexity of exome capture and sequencing was achieved by adding unique asymmetric 10-bp barcodes to the DNA fragments of single samples during library amplifications. Equal molar amounts of DNA samples were pooled for exome capture using a slightly modified version probe library of xGen exome research panel from Integrated DNA Technology. After PCR amplification and quantification of the captured DNA, samples were multiplexed and loaded to Illumina sequencing machines for sequencing to generate 75-bp paired end reads. The samples in this study were sequenced using Illumina sequencing machines including HiSeq 2500, and NovaSeq 6000 with S2 or S4 flow cells.

### Read mapping and variant calling

We used bcl2fastq (Illumina) to generate sequencing reads in the FASTQ format from Illumina image data. Following the original quality functional equivalent (OQFE) protocol^142^, BWA MEM^143^ mapped sequencing reads to GRCh38 references in an alt-aware manner, read duplicates were marked, and additional per-read tags were added. A Parabricks accelerated version of DeepVariant (v0.10) with a custom model identified single nucleotide variants and short insertion and deletions and produced a per-sample genome VCF^144^ which were aggregated with GLnexus (v1.4.3)^145^ into joint-genotyped multi-sample project-level VCF (pVCF). PLINK 1.9^146^ converted the pVCF to PLINK files for downstream analyses. Finally, we performed genotype-level quality control on the X chromosome by setting male heterozygous genotypes on the non-pseudoautosomal region of chromosome X to missing using the PLINK2^146^ option --set-hh-missing.

### Exome sequencing quality control (QC)

We implemented a support vector machine (SVM)-based QC protocol to filter likely artifactual variants as previously described^9^. Positive controls included: 1) genotype calls with ≥ 99% concordance between exome sequencing and array data; 2) transmitted singletons; and 3) an external set of likely “high quality” variants defined from 1000 genomes phase 1 high-confidence SNPs and 1000 genomes gold-standard indels, further restricted to the intersection between variants that pass QC in TOPMED Freeze 8 and gnomAD v3.1.2 genomes. Negative controls included: 1) Mendelian inconsistent variants; 2) discordant genotype calls in genomic duplicate samples; and 3) intersection of gnomAD fail variants with TOPMED Freeze 8 Mendelian or duplicate discordant variants. Prior to model training, we subset the control set of variants to the exome target capture region, binned by AAF, and then randomly sampled such that an equal number of variants were retained in the positive and negative labels. The model was then trained on up to 36 available site quality metrics (e.g., the median value for allele balance in heterozygote calls and whether a variant was split from a multi-allelic site). Even and odd chromosomes were then split into train and test sets, respectively. We performed a grid search with 5-fold cross-validation on the training set to identify the hyperparameters that return the highest accuracy during cross-validation, which are then applied to the test set to confirm accuracy (Precision=0.92, Recall=0.98, F1=0.95, AUC=0.98). While we did not filter variants based on SVM predictions, we used these predictions to create individual-level covariates for GWAS (see **Genetic association analyses**).

### Variant annotation

Variant Effect Predictor (VEP)^147^ v100.4 based on Ensembl^148^ (release 100) human protein-coding transcript models was used to annotate variants. We used a single consequence per variant based on its annotation on the canonical transcript for all the analyses described. One canonical transcript per gene was defined using a combination of MANE^148^, APPRIS^149^ and Ensembl canonical tags following the same procedure as described previously^148^.

### Genetic association analyses

We used the linear whole-genome regression test implemented in REGENIE^132^ to perform three core genetic analyses on chromosomes 1-22 and X in the discovery: 1) a TOPMed^150^ imputed common variant GWAS (AAF>0.01; N=19,527,870 variants), 2) a rare variant exome analysis (AAF≤0.01, alternate allele count [AAC]≥10; N=9,086,709 variants excluding those tested in the imputed variant GWAS), and 3) a gene-based association analysis of rare coding variation (N=19,107 gene-level omnibus tests). We set a single unified Bonferroni-adjusted significance threshold of *P*<1.75×10^-9^ across all three analyses accounting for all common, rare, and gene-based tests performed (0.05 / 28,633,686 tests). For a discussion of calibration in these results, see the **Supplementary Note**.

For the non-pseudoautosomal regions of chromosome X, we assumed a dosage compensation model where homozygous reference males are coded as 0 and hemizygous males are coded as 2 (and heterozygous males set to missing). In step 1 of REGENIE (*i.e.,* prediction of individual trait values based on the genetic data) we included directly genotyped variants with an AAF >1%, <10% missingness, Hardy-Weinberg equilibrium test *P*-value > 10^-15^ and linkage-disequilibrium (LD) pruning (1000 variant windows, 100 variant sliding windows and *r*^2^ threshold 0.1). The association model used in step 2 of REGENIE included the following covariates (**Supplementary Table 24**): age, age^2^, sex, age × sex, age^2^ × sex, the first 10 common-variant principal components (PCs) derived from the analysis of a stricter set of LD-pruned (1000-variant windows, 50 variant step size, and *r*^2^ threshold 0.1) common variants from the array data, the first 20 PCs derived from rare variants (1000-variant windows, 50 variant step size, and *r*^2^ threshold 0.1), 95 country-level fine-scale ancestry predictions (see below), sample source covariates, DNA source covariates (e.g., blood, saliva, cell-line), sequencing platform covariates, sequencing batch covariates, the total number of failed support vector machine (SVM) variants per individual, the average number of failed SVM variants per sequencing flow cell, an indicator variable signifying whether the individual was in the top 1% of individuals ranked by their total number of failed SVM sites, another indicator variable whether the individual was sequenced in the worst 50% of flow cells ranked by the average number of failed SVM variants called in individuals sequenced on the flow cell, and interaction covariates between cohort and each of: age, age^2^, sex, age × sex, and age^2^ × sex.

All rare variant and gene-based association tests were performed conditional on the 3,034 common variants including each variant in the model individually using a leave-only-same-chromosome-in (LOSCI) approach. This approach included only the subset of common variants located on the same chromosome as the variant or gene being tested to reduce the number of variables in the model.

### Fine-scale ancestry (FSA)

We used a previously described method for genetic ancestry assignment^133^. Briefly, using a diverse reference panel of 11,354 individuals from 95 populations, we calculated the proportion of haplotype sharing of each sample and assigned them to one of six genetic ancestries (AFR, AMR, EAS, EUR, MEA, SAS) if they had greater than 50% of that genetic ancestry. Individuals with no majority amongst these six genetic ancestries were assigned to an admixed group (OTH). We recognize these commonly used labels oversimplify an individual’s genetic, cultural, and environmental diversity and use them only out of convenience in properly adjusting association tests for genetic ancestry.

### Alternate allele frequencies (AAFs)

For each variant across the imputed and exome datasets, we calculated the AAF for each ancestry and to assign a single AAF to each variant, we used the ancestry maximum AAF. Using the ancestry maximum AAF in the imputed data, we found 19,514,636 well-imputed (INFO ≥ 0.3 in all discovery cohorts) variants with an AAF ≥1%, which we hereto define as common variants. Taking the maximum AAF across ancestries in the exome data, we found 44,600,231 variants with a maximum AAF < 1%, which we hereto define as rare variants. When a variant was rare (<1%) in the exome, but common in the imputed (≥1%), we elected to place those variants in the common bucket (N=335,522). REGENIE used these allele frequencies when writing burden test genotypes and running the gene-based tests. For the replication analysis, we used the same AAFs as the discovery but included an additional 12,777,247 variants that were unique to the replication data. Singletons in the replication comprised variants that were either 1) a singleton in the discovery or 2) a singleton in the replication that was absent from the discovery.

### Iterative conditional analysis

To identify a set of height-associated, conditionally independent common variants in our GWAS, we performed an iterative conditional meta-analysis on the TOPMed-imputed dosage data. Association tests were performed in REGENIE on all variants with AAF > 0.01. At each iteration, distance-based clumping was performed on all variants with *P* < 1.75×10^-9^, enforcing a minimum distance between clumps of 1 Mb. The lead variant of each clump was then added to the list of variants on which to condition in subsequent iterations. Each iteration used the leave-only-same-chromosome-in approach described above and repeated 22 times until no variants with *P* < 1.75×10^-9^ remained.

### Omnibus Gene-Based Testing

To properly adjust for multiple correlated tests across a large number of genes (in our study, only the autosomes and chromosome X), we used the gene-P^10^, an omnibus gene-based test framework, that uses ACAT^151^ to combine *P*-values from four complementary rare variants gene-based tests - BURDEN-ACAT^10^, SBAT^10^, SKAT-O^12^, and ACAT-V^11^ - into a single *P*-value per gene (**Supplementary Figure 1**). We applied a Bonferroni threshold of *P* < 1.75×10^-9^ to adjust for the number of common-variant (N=19,527,870), rare-variant (N=9,086,709), and gene-P gene-based tests (N=19,107) performed.

BURDEN-ACAT^10^ uses ACAT^151^ to combine 35 individual burden tests into a single *P*-value. Variants predicted to result in frameshift, stop gain, or alteration to essential splice-donor or splice-acceptor sites were classified as pLoF. Missense variants were then evaluated using six *in silico* pathogenicity prediction algorithms – ESM-1v^152^, SIFT^153^, PolyPhen2+HumDiv^154^, PolyPhen2+HumVar^154^, MutationTaster^155^, and LRT^156^. Variants were selected for inclusion in burden tests based on AAF, status as pLoF (stop gained, frameshift, splice donor and splice acceptor), and – for missense variants - how many of the pathogenicity prediction algorithms predicted the variant to be deleterious. Missense variants were classified as “deleterious” if predicted pathogenic by ESM-1v (using a threshold of 0.78) or all the other five algorithms; and “possibly deleterious” if predicted not pathogenic by ESM-1v but pathogenic by 1, 2, 3, or 4 of the other algorithms. Burden tests were then performed using every combination of five different alternate allele frequency thresholds (where the alternate allele frequency corresponded to the maximum across ancestries; **Methods**) – singleton (4.17×10^-7^), 0.0001, 0.001, 0.005, and 0.01 – and seven different combinations of variant classes – 1) pLoF, 2) pLoF + deleterious missense + stop lost + start lost, 3) pLoF + at least possibly deleterious missense + stop lost + start lost, 4) pLoF + all missense + stop lost + start lost, 5) deleterious missense + stop lost + start lost, 6) at least possibly deleterious missense + stop lost + start lost, and 7) all missense + stop lost + start lost – yielding 35 burden tests. These 35 *P*-values were then combined using ACAT to yield the BURDEN-ACAT *P*-value.

The Sparse Burden Association Test (SBAT)^10^ is performed by modeling the 35 burden tests jointly using non-negative least squares regression, separately in the positive direction and negative direction. ACAT^151^ combined the two directional *P*-values into a single SBAT *P*-value.

SKAT-O^12^ was performed at an AAF threshold of 0.01 using eight values of *ρ*: 0, 0.01, 0.04, 0.09, 0.16, 0.25, 0.5, and 1. For each of the seven variant class combinations listed above, the *P*-value was computed for each of the eight *ρ* values and combined using ACAT^151^. The seven variant class combination-specific *P*-values were then combined in another round of ACAT^151^ to give one per-gene SKAT-O *P*-value. Similarly, ACAT-V was performed by first using ACAT to combine single-variant *P*-values separately for each variant class combination and then combining the *P*-values for the seven variant class combinations using ACAT again. Both SKAT-O and ACAT-V grouped together all variants with AAC≤3 into a single pseudo-variant which is then included in the tests.

To provide a framework applicable to other traits, we examined the component tests underlying gene-P. After identifying genes associated via gene-P, we assessed four tests (BURDEN-ACAT, SBAT, SKAT-O, and ACAT-V) to determine which yielded significant associations (**Extended Data Figure 1A**). Genes were classified as burden-associated (*P*<1.75×10□□ in BURDEN-ACAT or SBAT) or non-burden-associated (*P*<1.75×10□□ in ACAT-V or SKAT-O and *P*≥1.75×10□□ in both BURDEN-ACAT and SBAT).

### Comparing common and rare variant burden effect sizes

To compare the effect sizes of rare pLoF variants to common variants identified by GWAS at the same genes, we linked conditionally independent GWAS variants for height to their nearest gene via genomic distance. GWAS loci were then restricted to those whose nearest gene harbored a significant singleton pLoF association with height. For genes with multiple nearby GWAS variants, we computed the mean absolute GWAS effect size. For each gene *g* with *k_g_* GWAS loci (where *k_g_* > 0), we computed the ratio of the singleton absolute pLoF effect size to the GWAS conditional joint effect size. Specifically, for gene *g* with singleton pLoF absolute effect size 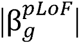 and a set of GWAS conditional joint effect sizes 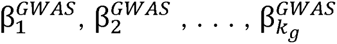, we defined the per-gene ratio as:

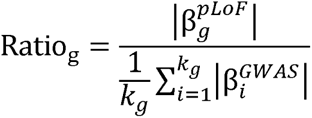

These ratios quantify the fold-increase in phenotypic effect conferred by complete gene disruption (via a singleton pLoF variant carried by a single individual) relative to the effect of common regulatory or coding variation at the same locus identified through GWAS. A ratio > 1 indicates that the singleton pLoF variant has a larger effect on height than the nearby common GWAS variant, consistent with the expectation that rare, gene-disrupting variants tend to have larger per-allele effect sizes than common variants subject to stronger purifying selection.

### Common and rare variant polygenic score (PGS)

We built a common variant PGS using PLINK^146^ from a total of 3,039 common (**Supplementary Table 8**) variants from two sources: first, 3,034 conditionally independent common variants identified via iterative conditional meta-analysis (**Methods**; **Supplementary Table 2**) and second, five common variants identified from the exome and not tested with the imputed data. The common variant PGS was computed as a weighted sum of alternate alleles, where the magnitude and direction of the weight was based on the estimated variant effect size (in SDs) from a joint model in the discovery (N=826,066; **Supplementary Table 1**) of all 3,039 variants (**Supplementary Table 8**) and the same covariates (**Supplementary Table 24**) used in the association analyses (**Supplementary Note**).

We constructed four versions of rare variant PGS (rvPGS), based on the 179 genes with significant gene-P and either a significant BURDEN-ACAT or SBAT test and weighted using effect sizes in SDs: 1) rvPGS_SNP_: all 77 significant (P < 1.75×10^-9^, AAC≥10) rare variants in these genes (after LD clumping for variants with R^2^ < 0.2), weighted by the single-variant effect size; 2) rvPGS_pLoF_: all 179 pLoF (AAF < 1%, AAC≥10) burdens weighted by the burden effect sizes; 3) rvPGS_missense_: all 179 deleterious missense (AAF < 1%, AAC≥10) burdens weighted by the burden effect sizes; and 4) rvPGS_synonymous_: all 179 synonymous (AAF < 1%, AAC≥10) burdens weighted by the burden effect sizes. When fitting prediction models based on both rvPGS_SNP_ and burden-based rvPGS, both the burden genotypes and burden effect sizes were recomputed to exclude the variants included in the single-variant rvPGS to enforce independence between the rvPGS.

### Variance explained by common and rare variants

All estimations of variance explained by the PGS were performed using genes and effect sizes discovered in the discovery (N=826,066) but estimated using the replication (N=242,679 excluding *All of Us*; **Supplementary Table 1**). First, for each cohort-sex combination (five cohorts × two sexes), a linear regression model was fit of height (in cm) on the covariates used in the replication association tests. The PGS of interest was added into the model, and the variance explained was calculated as the difference in adjusted R^2^ between the two nested models. The reported value was the mean of the ten cohort-sex subsets (five cohorts × two sexes). When computing variance explained by rare variant PGS, the null model included the 3039 common variant PGS (**Supplementary Table 8**) in addition to the covariates mentioned above.

### Prediction analyses

In replication cohorts (N = 242,679; excluding *All of Us*; **Supplementary Table 1**), we predicted an individual’s height (in cm) from a common variant PGS by fitting a linear regression model (separately by cohort and sex) in replication data with observed height (in cm) as the dependent variable and a set of independent variables comprising covariates (**Supplementary Table 24**) and the common variant PGS. Although height differs by genetic ancestry, we did not stratify predictions explicitly by genetic ancestry to avoid excluding subpopulations with few individuals (**Supplementary Table 1**). However, most of the replication cohorts primarily comprised individuals of a single genetic ancestry (excluding *All of Us*), so stratification by cohort was effectively the same as stratifying by genetic ancestry.

To predict an individual’s height using rare variants, we added various rvPGS to the same linear regression model as above. Because of the nested nature of burden tests with respect to AAF (e.g., a burden test of <1% AAF variants also includes all the variants in a singleton burden test), we restricted to only selecting a specific AAF for each class of variants. We also ensured that in any predictive models including both burdens and significant single variants, the burden effects were recomputed without any significant single variants. To assess the accuracy of any PGS, we compared the predicted height (in cm) to the individual’s observed height (**Supplementary Note**). We refer to the absolute difference in the observed and predictive height as *predictive error*.

### Enrichment analyses

To assess whether height-associated genes identified through rare variant burden testing were enriched for biologically relevant annotations, we performed a series of enrichment analyses across gene sets defined by variant consequence and allele frequency category. For each gene set, unless otherwise noted, we tested enrichment against the background set of all tested genes (n = 19,107). Annotations evaluated included OMIM disease genes (all and stature/skeletal growth-related, curated in Yengo *et al*.^2^), constrained genes (defined as the first LOEUF quintile from gnomAD v4.1), and nearest gene to a GWAS signal. For binary annotations, we used two-sided Fisher’s exact tests to estimate odds ratios with 95% confidence intervals; when odds ratios were infinite due to zero cells, we applied a continuity correction of +1 to all cells. For gene constraint, we also performed Wilcoxon rank-sum tests comparing the LOEUF rank distributions of test and background gene sets.

To address potential confounding by coding sequence length, whereby longer genes may be more likely to harbor rare qualifying variants and reach significance, we performed a canonical coding-sequence length (Ensembl release 100) matched enrichment analysis. For each gene, we identified the five closest background genes by canonical coding sequence length, sampled without replacement, to construct a matched null set and repeated all analyses described above using these CDS-matched background genes.

We used a two-sided 2×2 Fisher’s exact test to assess whether individuals carrying rare variants were more likely to be observed in the subset of the population based on various thresholds (in standard deviations [SD]) of either their *predictive error* (i.e., the absolute difference in the observed and common-variant predicted height; **Supplementary Table 18**) or their observed height (**Supplementary Table 19**). The set of genes we examined included either genes with a significant (*P*<1.75×10^-9^) burden test (e.g., 17 genes from singleton pLoF burden tests, 8 genes from singleton deleterious burden tests; **Figure 1B**; **Supplementary Table 6**) or all 179 significant gene-P genes with a significant SBAT or burden-ACAT test (**Figure 1D**; **Supplementary Table 4**). While varying the variant class, AAF threshold, gene set, and SD threshold, we kept the comparison group constant, defaulting to <0.5 SD from the mean with the format of the 2×2 table as below:

|  | Carries a rare variant | Does not carry a rare variant |
| --- | --- | --- |
| >X SDs |  |  |
| <0.5 SDs |  |  |

### Constraint analyses

LOEUF follows a uniform distribution^18^ and of the 207 genes identified in our gene-based tests, 201 had corresponding LOEUF values (gnomAD v4.1; *FGD1*, *HDAC6*, *IRS4*, *NRK*, *SCMH1*, and *SHOX* were missing in LOEUF). All analyses used the set of genes with LOEUF values. When comparing whether two gene sets significantly differed with respect to LOEUF, we used a Wilcoxon rank sum test using the LOEUF ranks. We considered genes to be LoF-constrained if they were in the first quintile of the LOEUF distribution.

### Comparison of association results with ClinVar annotations

When comparing height association results to ClinVar, height association statistics from the meta-analysis of discovery and replication cohorts (N = 1.45M) were used to maximize sample size. For consistency, the list of gene-P genes used for subsetting variants was the list of 207 genes from the discovery only (**Supplementary Table 4**). ClinVar data were downloaded in July 2025. To subset for diseases with a stature-related component, the union of three filters was used:

1. String matching of ClinVar disease column for any of the following strings: “stature”, “height”, or “chondroplasia”
2. Searching for any of the following manually curated, height-related Human Phenotype Ontology terms in the list of observed phenotype codes: HP:0004322, HP:0003510, HP:0001519, HP:0003498, HP:0003508, HP:0008909, HP:0003502, HP:0008845, HP:0008857, HP:0008848, HP:0011405, HP:0011406, HP:0011404, HP:0011407, HP:0000098, HP:0008873, HP:0003521, HP:0008921, HP:0008922, HP:0008929, HP:0008905, HP:0000002
3. Searching the observed phenotype codes for any height-related MONDO terms (**Supplementary Table 25**). This list was generated by MONDO label, definition, and synonym fields for the strings “stature”, “height”, and “chondroplasia” and removing any field with the string “normal stature.” 506 MONDO terms were identified in this way

Any ClinVar entry meeting any one of these criteria was included in the ClinVar analysis, resulting in 47,763 variants before filtering for those in gene-P genes. While not comprehensive (several variants in **Supplementary Table 21** affect known height genes but are not included), this list identifies a set of variants in ClinVar where the listed diseases are confidently determined to involve stature.

Enrichment testing for significant variants in each ClinVar pathogenicity bin (**Supplementary Table 22**) was performed using the binomial test, with the probability of success equal to 1.75×10^-9^.

## Data availability

Summary statistics from discovery analyses will be made publicly available upon publication. This study used data from the *All of Us* Research Program’s Controlled Tier Dataset v8, available to authorized researchers on the Researcher Workbench (https://www.researchallofus.org/). Data from the Mexico City Prospective Study are available to bona fide researchers for collaborative and/or open-access research purposes. The study’s Data and Sample Sharing policy can be downloaded (in English or Spanish: https://www.ctsu.ox.ac.uk/research/mcps). Available study data can be examined in the study Data Showcase (https://datashare.ndph.ox.ac.uk/mexico) and MCPS ancestry-specific allele frequencies are also available at: https://rgc-mcps.regeneron.com. The UK Biobank data are available to qualified researchers (http://www.ukbiobank.ac.uk/register-apply/). Data from the BELIEVE study are available upon reasonable request to the Data Access Committee. Further information on the application process can be found on the BELIEVE study website (https://believestudy-bangladesh.org/accessing-believe-data/). The list of 12,111 COJO SNPs and 462 OMIM height and skeletal growth related genes were downloaded from Supplementary Tables 10 and 11 of Yengo *et al.*^2^, respectively. LOEUF scores (gnomAD v4.1) for constraint were downloaded from https://gnomad.broadinstitute.org/downloads. ClinVar annotations were downloaded in July 2025 from https://ftp.ncbi.nlm.nih.gov/pub/clinvar/.

## Code Availability

Genetic data was represented in the PLINK format (v1.90b6.21) available at https://www.coggenomics.org/plink2/. REGENIE (v4.1), available at https://github.com/rgcgithub/regenie, performed all genetic analyses and wrote burden test genotype files. Meta-analyses were performed using METAL (2020-05-05) available at https://github.com/statgen/METAL. Imputation was done with Minimac4 (v1.01) available at https://github.com/statgen/Minimac4. PLINK (v2.00a3LM) built PGS. Burden heritability regression (v0.5.0-alpha) is available at https://github.com/ajaynadig/bhr. All other data analysis was performed using python (v3.11.4) and R (v4.3.2).

