## Supplementary Note for "Rare protein-coding variation and the genetic architecture of height in >1.4 million individuals"

***Table of Contents***

**Choice of common variant PGS effect sizes	3**

**The effect of conditioning gene-based tests on common genetic variation	4**

**Asymmetrical representation of OMIM height-increasing and -decreasing genes	5**

**Sensitivity analyses for gene set enrichment	6**

**Additional rare variants explaining poor height predictions	7**

**Genetic ancestry and sex analyses	8**

***CFTR* recessive association	11**

**Replication and evidence for additional genes to discover	12**

**Phenotypic variance explained by rare variants	16**

**Enrichment of rare variants in phenotypic outliers	17**

**Choice of metric for rare variant prediction	17**

**Rare variant prediction among individuals with poor common-variant prediction	18**

**Rare variant prediction among pLoF singleton carriers	19**

**Rare variant polygenic score discussion	21**

**Assessing calibration in association test results	23**

**Meta-analysis concordance	24**

**Genic constraint	25**

**Unusual distribution of variant effect sizes	26**

**Identifying rare gain-of-function variants	27**

**cvPGS x Gene Burden Interactions	27**

**List of contributors from the BELIEVE Study	29**

**List of investigators from the Colorado Center for Personalized Medicine Biobank	32**

**List of contributors from the GHS-RGC DiscovEHR Collaboration	33**

**List of investigators from the Mayo Clinic Project Generation	34**

**List of contributors from the Mexico City Prospective Study	35**

**List of contributors from the Penn Medicine BioBank	35**

**List of investigators from the Regeneron Genetics Center	36**

**List of investigators from the UCLA ATLAS Consortium	39**

**Supplementary Figures	40**

**References	56**

**Comparing common variant PGS effect sizes**

We elected to use a common variant polygenic score (PGS) constructed from effects in our discovery individuals (N = 826,066) as opposed to published effect sizes from a published study from Yengo *et al.* of approximately 5.3 million individuals^1^. This study provides two sets of PGS weights: one using approximately 12,000 variants and the other using approximately 1 million. Though the published PGSes explain slightly more variance than the one generated in this work, we elected to use the common variant results directly from the RGC-ME data for several reasons: 1) we preferred the direct iterative conditional analysis approach as opposed to approximate conditional analysis, 2) the inclusion of the X chromosome in our results, and 3) differences in population distributions. We performed analyses in our replication set (without *All of Us*) with the Yengo PGSes for comparison. The Yengo PGSes performed slightly better, however, we felt performance was similar enough to use the internally derived PGS and the impact on discovery and prediction is minimal.

| PGS | Mean R^2^ in any ancestry (N = 242,679) | Mean R^2^ in EUR only (N = 171,064) |
| --- | --- | --- |
| 3,039-variant PGS from discovery data | 0.247 | 0.284 |
| 12,111-variant PGS from Yengo, *et al*., 2022 | 0.274 | 0.327 |
| ~1 million-variant PGS from Yengo, *et al*., 2022 | 0.280 | 0.335 |

**The effect of conditioning gene-based tests on common genetic variation**

To assess the impact of conditioning the gene-based association tests on the 3,034 height-associated, conditionally independent common variants (**Supplementary Table 2**), we compared the association yield of gene-based testing with and without conditioning (**Supplementary Figure 2**; **Supplementary Table 12**). Prior to conditioning, 235 genes were significant with gene-P < 1.75×10^-9^. Of these, 26 (11%) genes with a marginal gene-P < 1.75×10^-9^ fell to more than two orders of magnitude below our Bonferroni threshold (gene-P > 1.75×10^-7^) after conditioning. Decomposing this into the component tests of gene-P, we found ACAT-V largely drives this loss of significance, where 21/120 (17.5%) of significant (*P_unconditional_* < 1.75×10^-9^) signals are not significant after conditioning, falling short of the significance threshold by over two orders of magnitude (*P_conditional_* > 1.75×10^-7^). This result is not surprising, as ACAT-V is expected to return a significant *P*-value in situations where a small number of variants in the gene are significant, rendering it susceptible to situations in which a gene-based signal is driven by a single low-frequency variant (AAF < 1%) in LD with a GWAS variant. We expect this situation describes most of the genes losing ACAT-V significance after conditioning. This is supported by the fact that burden-ACAT and SBAT based signals, which depend less on any individual variant, have the lowest relative decrease in yield post-conditioning, with 5.5% and 3.2% of significant signals lost (*P_unconditional_* < 1.75×10^-9^ to *P_conditional_* > 1.75×10^-7^), respectively. Lastly, SKAT-O fell between ACAT-V and burden with 25/193 (13%) of associated genes lost (*P_unconditional_* < 1.75×10^-9^ to *P_conditional_* > 1.75×10^-7^). Genes lost and gained from conditioning did not differ with regards to being the nearest gene to a GWAS locus (*P*=0.16; Fisher’s exact test), constraint (*P*=0.14; Fisher’s exact test), or being a height-related OMIM gene (*P*=0.60; Fisher’s exact test).

**Asymmetrical representation of height-increasing and height-decreasing genes in OMIM**

We performed additional analyses to examine possible explanations for the asymmetrical representation of height-increasing and height-decreasing genes in OMIM.

First, we examined whether height-decreasing genes were more pleiotropic, and therefore more likely to be included in OMIM for at least one of their pleiotropic trait associations. To assess the degree of pleiotropy amongst these genes, we queried published burden test results (AAF threshold < 1%) from 3981 traits (3,692 binary and 289 quantitative traits; **Supplementary Table 26**) from Backman *et al*.^2^ using the same *P-*value threshold as used in our study (1.75×10^-9^). We selected the most significant result when multiple burden tests were significant for the same gene-trait pair (this threshold and approach was also used to construct table columns indicating whether a gene was significant in the Backman study). At the *P* < 1.75×10⁻⁹ threshold, we identified 591 significant gene-trait associations (196 unique traits) amongst 64 of the 179 height-associated burden genes (**Supplementary Table 27**). Unsurprisingly, the majority of the 591 gene-trait associations involved quantitative traits (507 associations; 85.8%) compared to binary traits (84 associations; 14.2%), reflecting greater statistical power for quantitative traits. We observed that the distribution of quantitative, binary, and quantitative+binary trait associations was non-normal and zero-inflated - 115 genes (64.2%) showed no significant association for any trait at this threshold. Because of the non-normal distribution, we used a Mann-Whitney U test to compare the number of unique trait associations between gene sets. We observed on average 3.33 (95% CI: 1.10-5.56) trait associations per height-increasing gene and 3.53 (95% CI: 2.15-4.90) traits per height-decreasing gene, but the difference was not statistically significant (*P*=0.22; two-sided Mann-Whitney U test). We repeated this analysis for the 17 singleton pLoF genes (**Table 1**) and similarly did not observe a difference in the number of trait associations (*P*=0.54; two-sided Mann-Whitney U test).

We also examined whether the degree of constraint as measured by LOEUF was different between height-increasing and height-decreasing (**Methods**). We observed no difference in the LOEUF distribution between height increasing and height decreasing genes (*P*=0.27; Wilcoxon rank sum test) or when binarizing LOEUF into constrained and unconstrained gene sets (*P*=0.09; Fisher’s exact test). Similarly, restricting to singleton pLoF genes did not alter the conclusions for differences in the LOEUF distribution (*P* = 1; Wilcoxon rank sum test) or a binarized LOEUF version into constrained and unconstrained gene sets (*P*=1; Fisher’s exact test).

**Sensitivity analyses for gene set enrichment**

Coding sequence length can influence gene set enrichment analyses and thus we performed an additional set of sensitivity analyses to address this potential confounder (**Methods**). In general, we observed that controlling for coding sequence length did not result in null enrichment analyses for the nearest gene to a GWAS locus, OMIM genes, and stature/skeletal growth genes. On the other hand, enrichment of constrained genes was substantially attenuated after controlling for coding sequence length, not altogether unexpected given that the statistical test used to detect constraint has more power in longer genes as well as genes with higher mutations rates (as there are more observed mutations to compare to expectations)^3-5^.

For the 17 singleton pLoF genes (**Table 1**), all enrichments survived after controlling for coding-sequence length: nearest GWAS gene (OR=13.98; 95% CI=3.66-79.54; *P*=4.09×10^-6^; Fisher’s exact test), constraint (OR=8.47; 95% CI=1.85-79.21; *P* = 0.001; Fisher’s exact test), OMIM: (OR=3.17; 95% CI = 1.01-10.99; *P*=0.03; Fisher’s exact test), stature/skeletal growth genes (OR=5.88; 95% CI = 1.83-19.27; *P*=0.001; Fisher’s exact test). For genes associated with a <1% pLoF burden test, all enrichments but constraint survived after matching on coding sequence length: nearest GWAS gene (OR=8.95; 95% CI = 5.25-15.56; *P*=4.74×10^-18^; Fisher’s exact test), OMIM (OR=2.92; 95% CI=1.77-4.88; *P*=1.07x10^-5^; Fisher’s exact test), stature/skeletal growth genes (OR=2.49; 95% CI=1.37-4.39; *P*=0.001; Fisher’s exact test), constraint (*P*=0.04; OR=1.68; 95% CI=1.01-2.81; Fisher’s exact test). And lastly for burdens combining pLoF and missense variants, they all survived except for constraint: nearest GWAS gene (OR=8.10; 95% CI=5.63-11.72; *P* = 1.46×10^-31^; Fisher’s exact test), OMIM genes (OR=2.41; (95% CI=1.7-3.42; *P* = 3.5×10^-7^; Fisher’s exact test), OMIM stature/skeletal growth genes (OR=2.18; 95% CI=1.39-3.37; *P=*5.33×10^-4^; Fisher’s exact test), and constraint (*P*=0.16; Fisher’s exact test).

**Additional rare variants explaining poor height predictions**

Given our observation of large effect *FGFR3* and *PTPN11* missense variants that explained some individuals whose |predictive error| for height were more than 3 SDs, we examined whether rare variants not meeting our minimum alternate allele count threshold (AAC>9) might also explain some of the poor performance for the rvPGS. Given the predicted height for these individuals were off by >20 cm, we knew for a single variant to drive that difference, it must have a large $\beta$. With that in mind, we examined rare variants that, while statistically significant at the Bonferroni *P*-value threshold of 1.75x10^-9^, they were not reported due to low AAC and found 13 rare nonsynonymous variants (**Supplementary Table 14**). Of these 10 rare nonsynonymous variants with 1<AAC<10, three - 4:1805644:C:A (p.Asn540Lys) in *FGFR3*, 17:63917337:C:T (p.Arg290His) in *GH1*, 18:51078294:C:T (p.Arg496Cys) in *SMAD4 -* were present in the replication analysis and explain the large |predictive error| for the carriers (**Supplementary Figure 9**).

**Genetic ancestry and sex analyses**

When analyzing each genetic ancestry group separately, we did not identify any additional gene-based associations specific to non-EUR genetic ancestries. However, we did identify four genes that were significantly associated with height (gene-P < 1.75×10^-9^) in EUR samples when analyzing EUR alone, but not in the full cohort. These genes are shown in the table below:

| **Gene symbol (Ensembl ID)** | **gene-P, EUR only** | **gene-P, combined analysis** |
| --- | --- | --- |
| *DNMT1* (ENSG00000130816) | 2.74×10^-10^ | 4.35×10^-9^ |
| *SPIN4* (ENSG00000186767) | 7.32×10^-10^ | 1.46×10^-6^ |
| *WWC2* (ENSG00000151718) | 1.39×10^-9^ | 2.74×10^-9^ |
| *GRK5* (ENSG00000198873*)* | 1.60×10^-9^ | 4.15×10^-7^ |

Among the most-strongly associated burden masks per gene, we did not detect any significant heterogeneity in effect sizes across genetic ancestries after Bonferroni correction for the 19,105 genes tested (Cochran’s Q-test; ⍺ = 2.62×10^-6^).

Using an AAC threshold of 10, we also detected no new gene-based associations when analyzing sexes separately and similarly detected no heterogeneity in effect sizes across sexes.

We identified a small number of single variants that were significant (⍺ = 1.75×10^-9^) in one sex or genetic ancestry group (AAC > 9), but not in the combined analysis. As in the gene-based results, most of these genetic ancestry group-specific associations appeared in the EUR-only analysis because that group had the largest sample size. There were four EUR-only single-variant associations, comprising two missense, a 3’ UTR, and a synonymous variant (**Supplementary Table 15**). Two single variants were specific to the AMR-only analysis, and one more was specific to the female-only analysis (**Supplementary Table 15**). No single variant associations were identified that were significant only in the male-only analysis or any other ancestry-specific analysis with AAC > 9. Furthermore, no evidence of single-variant heterogeneity across sexes or genetic ancestries was detected among nominally significant (P < 0.05) exome variants.

One notable variant is *FBN1* p.E1297G (15:48481729:T:C), a missense variant that is enriched in the AMR genetic ancestry and has been independently shown to both be associated with reduced height in Peruvian individuals and demonstrate patterns of positive selection^6^. While this variant is associated in the AMR-only analysis more strongly than in any other single genetic ancestry group (**Supplementary Table 16**), it is even more strongly associated in the combined analysis. This suggests that while this variant is highly enriched in AMR individuals (246-fold relative to EUR and 59-fold relative to all non-AMR), the functional impact is consistent across ancestries. Although the second highest population-specific AAF for this variant was in admixed individuals with no majority ancestry (OTH; **Methods**), these individuals have high estimated global proportions of AMR ancestry (**Supplementary Figure 4**, see note below). Furthermore, as over 97% of carriers in the discovery data come from the MCPS cohort, the local ancestry inferred in that previously published study^7^ indicates that virtually all haplotypes carrying this variant have a high likelihood of indigenous origin (**Supplementary Figure 4**).

***CFTR*** 𝚫F508 **recessive association**

We investigated whether a recessive model could identify associations missed by the additive model, but only found one in the discovery data: the *CFTR* 𝚫F508 (7:117559590:ATCT:A) mutation that causes cystic fibrosis ($\beta_{rec}$=-0.73; 95% CI = [-0.97, -0.50]; $P_{rec}$ = 5×10^-10^) and also replicated it in an additional 624,567 individuals ($\beta_{rec}$ = -0.77 SD; 95% CI = [-0.94, -0.60]; *P* = 2.3x10^-18^).

Given the recessive nature of diseases like cystic fibrosis, we investigated how cohort composition – the enrichment or depletion of rare diseases like cystic fibrosis – could affect the discovery of non-additive associations. To this end, we examined the frequency of the variant (i.e., the AAF) and the homozygote alternate genotype frequency across cohorts (**Supplementary Table 28**). While the 𝚫F508 AAF varies between cohorts in accordance with ancestry proportions -- >1% in cohorts comprised of a majority of individuals of European descent (e.g., UKB, GHS, Penn) and lowest (<0.2%) in non-EUR majority cohorts (e.g., Mexico City Prospective Study or BELIEVE) -- the frequency of 𝚫F508 homozygotes differed considerably. Though the ΔF508 allele frequency is highest in UKB (0.0153), there are only two homozygous individuals compared to ~106 expected under Hardy-Weinberg equilibrium (observed/expected ratio = 0.02). This ~50-fold depletion is consistent with UKB's well-documented healthy-volunteer bias^8^, as individuals with cystic fibrosis – a severe, life-shortening disease – would be far less likely to participate in a general population study. Furthermore, the minimum age to participate in UKB was 40, and the median age of death for cystic fibrosis patients didn’t exceed 40 until 2014^9^. In contrast, Penn Medicine Biobank and All of Us have a ~7.5-fold and ~2.3-fold excess of ΔF508 homozygotes, respectively, likely due to being hospital-based cohorts. This creates a bidirectional ascertainment spectrum: health-system biobanks can enrich for disease while volunteer biobanks like UKB deplete it. Importantly, while homozygote counts vary dramatically, the variant AAFs are relatively stable across EUR-majority cohorts (0.011–0.015), suggesting that ascertainment bias primarily affects the frequency of homozygous carriers.

**Replication and evidence for additional genes to discover**

We meta-analyzed association results from exome or whole genome sequence data in 624,567 individuals (240,159 non-EUR, 38.4%) from six additional cohorts (**Supplementary Table 1**) to evaluate the robustness of our gene-based and single-variant association results. Ignoring power, 203 of the 207 gene-based associations (98.1%) replicate at a nominal gene-P (⍺ = 0.05; **Supplementary Table 4**; **Supplementary Figure 5A**). Replication rates were consistent across the different gene-based tests as well: BURDEN-ACAT: 98.1% (155/158), SBAT: 93.8% (166/177), SKAT-O: 95.3% (164/172), and ACAT-V: 94.9% (94/99; **Supplementary Figure 5B**-**E**).

We also assessed replication in the 126 individually significant exome variants (**Supplementary Table 3**). Of the 126 variants, 125 were also present in the replication data; without taking statistical power into account, the replication rate was 90.4% (113/125; **Supplementary Figure 5F**). Only one missense variant, p.Ala1225Ser in *UBR1* (15:42998252:C:A), had inconsistent effect directions between the discovery ($\beta$=-0.76 cm; *P*=4.30x10^-10^) and replication analysis ($\beta$=0.03 cm; *P*=0.88). After accounting for variants with >80% power, our replication rate increased to 96.1% (98/102; **Supplementary Table 3**).

While we used a stringent Bonferroni significance threshold that covers genetic associations across common, rare, and gene-based tests (**Methods**), we recognize others may opt to include in their Bonferroni correction only a subset of these tests (e.g., only gene-based tests). To this end, we repeated the replication analysis for gene-based tests (**Supplementary Figure 5A-E**), individual exome variants (**Supplementary Figure 5F**), and burden tests (**Supplementary Figure 6**) and assessed the replication rate at a nominal *P*-value threshold of 0.05. For gene-P and its four-underlying gene-based tests, all but SBAT maintained an acceptable false discovery rate of < 0.05 up to *P* < 1×10^-7^, suggesting that not only were we conservative in this study with respect to *P*-values, but that there are at least another 58 genes we could have claimed as significant via gene-P (265 genes discovered at gene-P < 1×10^-7^). Furthermore, all gene-based tests maintained a replication rate above 50% at *P* < 0.001. For those researchers interested in using a Bonferroni threshold that roughly corresponds to correcting for the ~20,000 genes in the genome (~2.5×10^-6^), replication rates remained respectable: 91.6% for gene-P (337/368 genes), 93.5% for BURDEN-ACAT (272/291), 84.0% for SBAT (272/324), 88.0% for SKAT-O (278/316), and 87.4% for ACAT-V (167/191). These results indicate most gene-based associations identified at or below *P* < 2.5×10^-6^ represent likely genuine signals and furthermore we would have discovered 77.8% more genes (368 vs. 207). While the replication rates across *P-*value thresholds for the gene-based tests were all very similar (all pairwise Pearson R>0.988) we observed that gene-P and Burden-ACAT consistently achieved the highest replication rates across all *P*-value thresholds, reflecting the ability of these omnibus-style tests to capture robust signals across multiple underlying groups. SKAT-O and ACAT-V, which are designed to be more powerful in the presence of variants with mixed directions of effect or when only a small fraction of variants are causal, showed slightly lower, but still high replication rates (**Supplementary Figure 5D** and **5E**). SBAT exhibited the lowest replication rates, though its performance remained strong at stringent thresholds (**Supplementary Figure 5C**).

For individual exome variants, replication rates followed the same monotonic trend observed for gene-based tests (**Supplementary Figure 5F**). The number of variants reaching significance also decreased markedly with more stringent thresholds: 62,564 variants were identified at *P* < 0.01, compared to 629 at *P* < 1×10^−5^ and 125 at *P* < 1.75×10^−9^. At the exome-wide significance threshold (*P* < 1.75×10^−9^), 90.4% of variants replicated (113 of 125 variants). The replication rate was 88.1% at *P* < 1×10^−8^ (140 of 159), 85.2% at *P* < 1×10^−7^ (184 of 216), and 74.4% at *P* < 1×10^−6^ (253 of 340). At lenient thresholds (*P* < 0.01), only 7.4% of variants replicated (4,620 of 62,564), consistent with the expectation that the vast majority of variants at this threshold being false positives.

We further examined the replication rates of burden tests stratified by variant functional annotation (mask) and AAF category (**Supplementary Figure 6**). Burden tests were evaluated across seven mask categories (missense, possibly deleterious missense, deleterious missense, pLoF, pLoF+missense, pLoF+possibly deleterious missense, and pLoF+deleterious missense) and five AAF thresholds (singleton, <0.01%, <0.1%, <0.5%, and <1%). Singleton burden test replication rates remained nearly unchanged from 1.75×10^-9^ to 1.75×10^-6^. Furthermore, at *P<*1.75×10^-6^ nearly twice as many pLoF singleton burden tests were discovered (33) when compared to our Bonferroni threshold of 1.75×10^-9^, again confirming both that we were extremely strict and that more genes are (unsurprisingly) left to be identified. However, at more relaxed thresholds (*P* < 0.01), singleton-only analyses showed notably lower replication rates (14.5–36.0% depending on the mask) compared to higher AAF thresholds reflecting the inherent noise and low power of singleton-driven signals at lenient *P*-value thresholds. This is expected, as higher AAF thresholds (≤0.5% and ≤1%) aggregate more common variants, yielding larger cumulative allele counts, more stable effect estimates, and greater statistical power in replication.

Across mask categories, the inclusion of pLoF variants—either alone or in combination with missense variants—generally yielded replication rates comparable to or higher than missense-only masks at matched AAF thresholds. Masks incorporating functional annotation to prioritize deleterious missense variants did not consistently improve replication rates compared to broader missense masks, suggesting that at the sample sizes and thresholds examined, the increased specificity of more restrictive masks is offset by reduced variant counts.

As an alternative analysis to examine whether genes that did not achieve significance in our discovery experiment with our conservative ⍺=1.75×10^-9^ threshold are truly associated with height, we leveraged the observation that approximately 2/3 of both our gene-based and single variant associations affect height in a negative direction. This excess of negative effects allows us to see evidence of true associations among genes with subthreshold significance. To this end, we quantified the proportion of negative burden signals for each variant class at AAF < 1% across several *P*-value bins (**Extended Data Figure 5**). The degree and persistence of the excess of negative effect estimates down to *P*<0.01 is consistent with estimates of phenotypic variance explained, both suggesting there is substantial additional signal left to discover.

**Phenotypic variance explained by rare variants**

With rare variants frequently offered as the answer to the “missing heritability” problem in complex trait genetics, we sought to estimate the cumulative impact of rare coding variation on height at both the population and individual level. We used burden heritability regression (BHR)^10^ to estimate the burden heritability, $h_{burden}^{2}$, which is defined as the fraction of phenotypic variance explained by the rare alternative allele burden of pLoF, missense, and synonymous variants in each gene. It is important to note that $h_{coding}^{2}\geq h_{burden}^{2}$^10^. We found $h_{burden}^{2}$ decreased with decreasing variant deleteriousness: pLoFs, being the most deleterious variant class with the largest effect sizes, explained the highest $h_{burden}^{2}$ at 1.94%, followed by deleterious missense variants at 1.63%, remaining missense variants at 0.41%, and synonymous variants at 0.09% (**Extended Data Figure 3**). Furthermore, we found that very rare pLoFs (AAF<1×10^-5^) explained more $h_{burden}^{2}$ than any other category (**Extended Data Figure 3**) and this trend extends to missense, but not synonymous variants. Collectively, rare nonsynonymous variants explain 3.97% of the phenotypic variation in height (95% CI=[3.77%, 4.17%]), nearly identical to the estimate of 3.82% (95% CI=[3.55%, 4.09%]) published by Weiner *et al*^10^. Furthermore, the 179 burden associations we identify here explain ~1.1% of the phenotypic variance in height in the replication data (**Methods**; **Extended Data Figure 4**).

To assess the extent to which heritability exists below our genome-wide significance threshold, we binned genes by gene-P and calculated the variance explained by each bin. While the most phenotypic variance could be explained by genome-wide significant associations, we found significant (likelihood ratio test, ⍺=0.0001 to adjust for 500 tests) phenotypic variability could be attributed to genes with gene-P<0.01 across nearly all variant classes except for synonymous variants (**Extended Data Figure 4**). In contrast to prediction from BHR, we estimate gene tests combining pLoF and deleterious missense variation explain the largest portion of phenotypic variability among rare nonsynonymous variation (mean variance explained across cohort-sex subsets of 0.82%).

**Enrichment of rare variants in phenotypic outliers**

It is often assumed individuals at the extremes of a phenotypic distribution are more likely to harbor rare variants of large effect and as such, studies are sometimes carried out specifically in these individuals. We sought to determine whether individuals with large predictive errors were more likely to harbor large effect rare variants than individuals at the extremes of the phenotype distribution.

As discussed in the main text, we observed that individuals with |predictive error| >2.5 SD are 22.7-fold enriched (95% CI = [11.3, 41.7]; *P* = 1.29×10^-12^; Fisher’s exact test) for singleton pLoFs in 17 singleton pLoF genes (**Figure 2A**; **Supplementary Table 18**). In comparison, selecting individuals with an observed height >2.5 SD (~26.75 cm) away from the mean were 4.8-fold enriched (95% CI = [2.39, 8.72]; *P*=2.14×10^-5^; Fisher’s exact test; **Supplementary Table 19**) more likely to carry a singleton pLoF variant in one of the 17 significant singleton pLoF burden genes than individuals <0.5 SD from the mean (**Methods**).

**Choice of metric for rare variant prediction**

We considered at length when deciding how best to present our results in an interpretable way. While previous results have been reported in terms of R^2^, we chose the average absolute residual of the height prediction model for two reasons:

1. Numerical interpretability: the average reader may find it easier to interpret a number that represents how far, on average, a prediction is from the correct value as opposed to R^2^, which measures explained variance.
2. Small sample size: some subpopulations in which we evaluated predictions were too small for R^2^ to be useful measure of prediction performance. This is a consequence of our focus on rare variants, which necessarily have few carriers. For example, in the bottom panel of **Figure 3D**, all carrier subpopulations evaluated had between 3 and 8 carriers. The subpopulations in the top panel were also small (counts shown in **Extended Data Figure 6**). In order to allow fair comparisons in prediction between the subpopulations in the text, we elected to use the average absolute residual everywhere.

However, we do report prediction results in R^2^ in **Figure 3B** and **Supplementary Figure 10B**.

**Rare variant prediction among individuals with poor common-variant prediction**

Given that deleterious rare variants in the genes we discovered are enriched among individuals with poor predictions from common variants (**Figure 3A**), we attempted to leverage this enrichment to improve predictive performance among these individuals. We explored various strategies to include rare variant PGS (rvPGS) in prediction (**Methods**), including adding rvPGS for significant rare variants only, including rvPGS for burden effects, and including both kinds of rvPGS (with significant individual rare variants excluded from the burdens). Overall, we found that including both kinds of rvPGS performed the best (**Supplementary Figure 7**) as it allowed for improved prediction both for those who carry rare, deleterious variants achieve significance in our study and those carrying variants too rare for significance (but where the affected gene is discovered by a burden test). Still, to understand why height predictions improved by only 0.37 cm on average among these 601 individuals for whom common variants predict height poorly (predictive error from common variants > 2.5 SD), we examined the distribution of prediction improvement achieved by adding rare variants (that is, predictive error_common_ - predictive error_common+rare_). For six individuals, incorporating rare variants made height prediction more than 20 cm more accurate than common variants alone (**Supplementary Figure 10**).

**Rare variant prediction among pLoF singleton carriers**

Even among individuals carrying pLoF singleton variants among the 17 significant pLoF singleton genes (**Table 1**), the mean improvement in prediction error when adding in pLoF singletons into the predictive model was modest at 1.8 cm (**Figure 3C**). This initially surprising result is likely due to two main factors: allelic heterogeneity and winner’s curse. Carriers in replication data were chosen based on carrying a gene-specific burden, annotation as a pLoF, and allele frequency. However, there is no guarantee that the individual variants carried by participants in replication data are included in the discovery data from which effects are estimated (even though we included all singleton variants in discovery as “singletons” in replication analysis regardless of their replication AAC, see **Methods**), particularly as sample size was not equivalent. For example, in *CHD8*, where height predictions in replication data become *worse* after adding pLoF singleton burdens in the prediction model (**Figure 3D**), there is no overlap between the pLoF singletons carried in the two datasets. As a result, unlike the *CHD8* pLoF singletons observed in discovery data, those observed in replication data may not result in loss-of-gene function despite their annotations.

As shown in **Figure 3D** and **Extended Data Figure 6**, however, this occurs in a minority of genes, and most genes do demonstrate substantial improvement in prediction when modeling rare variants in this way. This mean of 1.15 cm reflects an average across genes that do benefit from modeling rare variant burdens (e.g. *ACAN*) and those that do not (e.g. *CHD8*) and thus may be deflated relative to our expectations. Furthermore, this summary metric represents the mean value weighted by the number of carriers of each pLoF singleton burden (**Extended Data Figure 6**). To illustrate, *NF1* -- with 49 pLoF singleton carriers having a mean improvement of only 0.64 cm – carries more than twice as much weight as *ACAN*, whose 21 carriers improve by 5.60 cm on average, due to the number of carriers (**Figure 3D**).

Similarly, winner’s curse is likely a factor: effect sizes from discovery data may be overestimated for these rare variants where we were underpowered relative to the frequency in that dataset. Effect sizes are often attenuated when applied to a new dataset.

Some other potential explanations include:

1. The replication dataset in which we identified these pLoF singleton carriers (N = 242,679) is substantially smaller than the discovery dataset where these genes were discovered (N = 826,066), particularly as the replication data used for prediction excluded the *All of Us* cohort. As a result, the replication data are less well-powered to find rare functional variants in these largely constrained genes. Increased replication sample size might enable observation of more rare functional variation in these 17 genes. This would, in turn, increase the average prediction improvement from modeling pLoF singleton burdens.
2. Potential errors in phenotyping are substantially more problematic when focusing on a small number of individuals
3. Rare pLoF singletons observed only in replication data may have lower penetrance than those observed in discovery data.

**Rare variant polygenic score discussion**

Rare variant polygenic scores (rvPGSes) are still in their nascency with many unanswered questions regarding best practices. In attempting to implement rvPGS, we encountered several decision points, caveats, and limitations that we hope will be addressed by future researchers.

One of the first decision points was whether to use the associations from gene-based tests or from individual variants. As most rare variants from sequencing studies are too rare to be significant on their own (e.g., singletons)^3,11-13^ and their effect size estimates are poorly estimated^2^, we initially opted for using the gene-based test associations. However, this decision to use gene-based test effect sizes had four main caveats.

**Not all gene-based tests estimate a gene-based effect size.** Variants included in a PGS require an effect size (or at least an effect direction) to weight the genotypes, yet only burden tests compute effect sizes at the gene level. Thus, we could not include 28 genes identified in only ACAT-V^14^ or SKAT-O^15^ (**Figure 1D**) in the PGS.

**Burden tests using nested AAF thresholds are correlated and require selecting one burden test per gene.** The burden tests performed with gene-P all have nested AAF thresholds (e.g., singleton pLoFs are present in all pLoF burden tests regardless of AAF threshold). As such, for a given gene and variant class, burden test effect sizes are correlated across AAF thresholds, and carriers of a given variant may contribute to separate burden tests of the same gene. This is unfortunate as one may want to capture an accurate contribution of singleton variants (with a high average effect size) but that only apply to a small subset of individuals, while also capturing the higher frequency rare variants with weaker effect sizes but contribute to a larger proportion of the population. For our enrichment and prediction analyses, we had to arbitrarily choose between using rarer and less powered, but higher effect size, burden tests compared to more common and better powered, but lower effect size, burden tests. This decision led to either rvPGSes that were more sparse but larger in magnitude or slightly less sparse but weaker in magnitude.

**Burden tests of separate genes are not necessarily independent w**hen nonsynonymous variants reside in multiple canonical transcripts. This becomes most notable when read-through transcripts are tested. Three of our 207 height-associated genes are read-through transcripts (*AC073896.1*, *AC127029.3*, *AC004922.1*) which include most of the same exons as the canonical transcripts of *CNPY2*, *GH1*, and *ARPC1B*, respectively; in each of these three cases, both the read-through transcript and the gene are both statistically significant and included in the PGS, resulting in double-counting shared nonsynonymous variants in each gene.

**Burden tests average the effect sizes across contributing variants**^13,16,17^. While averaging the effect size across contributing variants is fine for most genes and variant classes (**Supplementary Figure 11**), in extreme cases this can result in the burden effect size significantly underperforming individual variants (**Extended Data Figure 5**). We encountered two instances – *FGFR3* and *PTPN11* – where the magnitude of the strongest burden test’s effect size was three to ten times weaker than that of statistically significant rare variants included in the gene burden.

With the caveats of using gene-base effect size estimates in a rvPGS, one might opt to use the individual rare variants instead. However, this approach is also not without issues – namely, our analysis identified fewer significant rare variants than gene-based tests after conditioning on common variants because single variants have less statistical power on average. Furthermore, not all rare variants are independent which further reduces the number of rare variants included in the PGS.

Another issue we encountered was that because one likely wants to examine the effects of the rvPGS in individuals at the phenotypic extremes, accurate phenotypes are extremely important. For quantitative traits that can be measured in different units, mixing units can lead to individuals who might normally reside in the center of the distribution ending up in the tails. Additionally, a recent study demonstrated 0.15% of UKB participants misrepresented their height in self-reports and these individuals are enriched for congenital malformations and rare pLoFs in Mendelian growth disorders^18^. While we did not use self-reported height in UKB, we cannot rule out that some of the height data collected in the various cohorts of this study included self-reported heights. While incorrect phenotypes certainly affect association results and reduce statistical power, our association analyses are not exclusively focused on phenotypic extremes and thus they are not affected to the same degree as PGSes focused on phenotypic extremes.

**Assessing calibration in association test results**

As has been extensively described over the years^1,19,20^, inflation in λ_GC_ can be due to true polygenicity, as for highly polygenic traits many variants have true effects. This raises the genome-wide median test statistic, so λ_GC_ > 1 can reflect true signal, not confounding. λ_GC_ scales with sample size; even perfectly calibrated, confounder-free studies often show λ_GC_ > 1 at large N. Nonetheless, we have also added λ_GC_ for several subsets of our analysis in a separate section of **Supplementary Table 16**. In conjunction with the more informative LDSC/BHR results above, we do not find these λ_GC_ values to signal anything of obvious concern.

Common variant λ_GC_ = 3.57

gene-P synonymous λ_GC._ = 1.732

gene-P nonsynonymous λ_GC._ = 2.684

gene-P pLoF λ_GC._ = 1.676

all burden tests λ_GC._ = 1.308

**Meta-analysis concordance**

To evaluate our mega-analysis approach (i.e., performing association tests using a multi-cohort joint callset), we compared our single-variant (**Supplementary Figure 12**) and gene-based association results to those yielded by a meta-analysis approach (i.e., performing association tests separately by cohort-ancestry group and combining summary statistics). We performed fixed-effects meta-analysis across 24 cohort-ancestry groups using METAL^21^, excluding cohort/ancestry groups with fewer than 485 samples to avoid fitting models with more covariates than samples. Overall, the results were highly concordant, with -log_10_*P* and effect sizes for single-variant tests showing adjusted R^2^ of 0.93 and 0.97, respectively (**Supplementary Figure 13**). Furthermore, pLoF (AAF < 1%) -log_10_*P* and effect sizes showed adjusted R^2^ of 0.96 and 0.98 (**Supplementary Figure 14**). One notable difference was the number of significant gene-based results; whereas the meta-analysis yielded 180 and 80 genes with gene-P or pLoF (AAF < 1%) burden *P* < 1.75×10^-9^, respectively, the mega-analysis yielded 207 and 81. This finding underscores the benefit of access to individual-level genotype data and joint analysis, as opposed to summary-level data only.

**Genic constraint**

We observed an enrichment of constrained genes amongst the 17 singleton pLoF genes (based on loss-of-function observed/expected upper-bound fraction [LOEUF]^11^ from gnomAD v4.1; **Table 1**; **Supplementary Table 5**).

Given these observations, we investigated whether genes associated with human height identified using gene-based tests were enriched for constrained genes using LOEUF^11^. We observed that height associated genes are enriched in genes under constraint (*P*=3.85×10^-11^; 𝛸^2^=67.94; 9 degrees-of-freedom chi-square test). LOEUF correlates with both singleton pLoF and singleton deleterious missense effect sizes in both directions and explains 15% and 25% of the variance in effect sizes for singleton pLoF burden tests with a positive or negative effect size, respectively (**Supplementary Figure 15).**

**Unusual distribution of variant effect sizes**

We observed an order-of-magnitude disparity between the *FGFR3* missense burden test with the largest effect size ($\beta$=-0.29 SD; 95% CI=[-0.33,-0.25]; *P*=2×10^-41^; deleterious missense variants; AAF<1×10^-4^; **Supplementary Table 13**) and the ClinVar pathogenic, gain-of-function missense variant for achondroplasia^22^, 4:1804392:G:A (*P*=3.22×10^-64^; $\beta$=-3.73 SD; 95% CI=[-4.16, -3.29]; **Supplementary Table 3**). Such a large difference in effect sizes between individual variants and the burden test suggests a wide distribution of single-variant effect sizes within the gene, as the burden effect size is effectively a weighted average of the individual variant effects. Given this observation, we investigated whether any other genes in the genome might also harbor a small number of variants with outlier effect sizes and thus have discordance between burden test effect sizes and those estimated from the contributing variants. To search for such genes, we measured the standard deviation of the effect size distribution per-gene, per-variant class (e.g., synonymous, missense, pLoF) of rare (AAF<0.01; alternate allele count > 9) variants and restricted to genes with at least 10 separate variants. For each variant class, we examined the distribution of standard deviations (**Supplementary Figure 11**) and only two genes were 12 SDs from the mean: *FGFR3* ($\sigma_{\beta}$=0.66) and *PTPN11* ($\sigma_{\beta}$=0.54). An additional three genes, *FGD1* ($\sigma_{\beta}$=0.40), *IGF2BP2* ($\sigma_{\beta}$=0.42), and *NPR2* ($\sigma_{\beta}$=0.46) were 10 SDs from the mean.

**Identifying rare gain-of-function variants**

We also examined whether any genes harbored rare missense variants with effects opposite their pLoF burden test (*P* < 1.75×10^-9^), as this might suggest the missense variant acted via a gain-of-function mechanism. However, we only found one: *FBN1*, where missense variant p.Tyr1696His (15:48463220:A:G) was associated with -7.77 cm lower height (*P*=4.3×10^-10^) while both the pLoF burden (singleton $\beta$=11.14 cm; *P*=1.45×10^-23^) and the deleterious missense burden (singleton $\beta$=8.06 cm; *P*=3.49×10^-23^) were associated with increased height.

This result raised the question of whether our results could provide an estimate on the proportion of missense variants with a direction of effect consistent with the estimated direction of effect from pLoFs. Again, setting our search space to genes with both a statistically significant missense variant and pLoF burden left us with 14 genes collectively harboring 21 missense variants. As discussed above, all but the *FBN1* missense variant had a consistent direction of effect with the pLoF burden (i.e., 20/21 missense variants), placing a very rough estimate of 95.2% of missense variants acting in the same direction as pLoF burden tests.

**cvPGS x Gene Burden Interactions**

We next sought to detect interactions between rare variant burdens and common variant PGS to understand whether variation in any of our discovered genes modify the effect of common height associations genome-wide. Because we limited our search to the 179 discovered by burden tests, we set a conservative Bonferroni significance threshold of $\alpha=\frac{0.05}{35*179}=8.0\times{10}^{-6}$ to account for all 35 burden tests performed per gene. At this threshold, two genes had burdens that demonstrated significant interactions with cvPGS: *GLS* and *PTPN11*; rare variants in each gene were associated with a smaller effect of cvPGS on height in discovery data (**Supplementary Table 29; Supplementary Figure 16**). While these interaction signals did not replicate in independent data (**Supplementary Table 29**), this is likely due to winner’s curse and low statistical power resulting from 1) modest interaction effect sizes and 2) the smaller sample size of our replication dataset. In fact, it is likely that there are other true interactions that even our discovery dataset does not have statistical power to detect. Suggestive evidence of this comes from the interaction effect between the cvPGS and a burden of pLoF singletons in *FGFR3*, the gene responsible for achondroplasia^22^, which is twice as large as that of *PTPN11*, but not significant (β = -0.45 height SD (*FGFR3* allele × cvPGS unit), *P* = 0.119).

*GLS* codes for glutaminase, an important enzyme for mitochondrial energy generation via the Krebs cycle^23^, and which has several Mendelian syndromes reported in OMIM. While the interaction signal in our data is strongest from the pLoF + all missense test with AAF < 1%, over 93% of carriers of this burden carried a missense variant, and the missense-only burden has a nearly identical interaction effect. This may suggest a gain-of-function (GoF) mechanism interacting with the cvPGS, and a rare missense GoF variant has been reported in this gene^24^.

*PTPN11* codes for protein tyrosine phosphatase non-receptor 11, and rare variants in this gene have been reported in OMIM to be associated with Noonan syndrome, of which short stature is a component^25^. The strongest cvPGS interaction in *PTPN11* is with rare deleterious missense variants, which agrees with reports of (potentially GoF) missense variants in this gene causing Noonan Syndrome^26^.

**List of contributors from the BELIEVE Study** (listed alphabetically within each category)

**Recruitment Coordinators and Field Supervisors**: Israt Akter^10^, Laboni Akter^10^, Md Shehab Uddin Al-Abid^10^, Arnab Aditya Das^10^, Khairul Islam^10^, Mir Shahadul Islam^10^, Shoriful Islam^10^, Mohammad Kamruzzaman^10^, Md Taslim Uddin Miah^10^, Monalisa Moni^10^, Sabrina Monsur^10^, Md Mostafa Monower^10^, K M Thouhidur Rahman^10^, Anjuman Ara Rahman^10^, Mantaka Rahman^10^, A H M Rezwan^10^, Jayashree Saha^10^, Sharraf Samin^10^, Kazi Nazmus Saqeeb^10^, Monjeline Sultana^10^, Ishrat Tasmin^10^

**Study Site Support Workers**: Md Rezaul Karim Akanda^10^, Jesmin Akhter^10^, Ayasha Akter^10^, Bakul Akter^10^, Jharna Akter^10^, Jesmin Akter^10^, Juba Akter^10^, Khadiza Akter^10^, Khadija Akter^10^, Lipi Akter^10^, Maksuda Akter^10^, Mousumi Akter^10^, Mst Lovely Akter^10^, Nahida Akter^10^, Nasima Akter^10^, Samima Akter^10^, Sema Akter^10^, Shahida Akter^10^, Shahnaj Akter^10^, Shamima Akter^10^, Taslima Akter^10^, A T M Zorjis Alam^10^, Mahmuda Atique^10^, Lutfa Begum^10^, Mst Nazma Begum^10^, Farjana Choudhury^10^, Md Zakir Hossain Chowdhury^10^, Mitali Paul Chowdhury^10^, Robin Reza Chowdhury^10^, Mukul Rani Debnath^10^, Kaniz Fatema^10^, Nahid Ferdash^10^, Naima Ferdous^10^, Md Rakib Al Hasan^10^, Khandaker Hashanuzzaman^10^, Shamima Haq^10^, Md Riazul Haque^10^, Alamgir Hossain^10^, Md Ibrahim^10^, Nusrat Jahan^10^, Shahi Israt Jahan^10^, Israt Jahan Jarin^10^, Zohora Pradhan Jonaki^10^, Asik Kabir^10^, Tonema Kader^10^, Md Mostafa Kamal^10^, Sayed Kamruzzaman^10^, Sadik Fatima Kanon^10^, Nazmul Karim^10^, Shamsul Karim^10^, Tanuja Khanom^10^, Shamim Ara Khatun^10^, Badrun Nahar Lorin^10^, Mst Sirajum Manira^10^, Farhana Jahan Mary^10^, Kazi Dilruba Mita^10^, Lipi Mitra^10^, Basudeb Mollik^10^, Kamrun Nahar^10^, Sudipta Nargis^10^, Nusrat Alam Nawmee^10^, Esrat Zahan Nesa^10^, Mahmuda Akter Nipa^10^, Mehenaz Parvin^10^, Sanjida Parvin^10^, Shahnaj Parvin^10^, Suraiya Parvin^10^, Kaniz Fatema Priya^10^, Golam Mostafa Quadrey^10^, Nayan Rabidash^10^, Rulia Rahman^10^, Madhabi Rani^10^, Shahjalal Sarker^10^, Smriti Sarker^10^, Razia Sultana Shathi^10^, Fatema Shelly^10^, Ireen Sultana^10^, Rovaiya Sultana^10^, Israt Jahan Sumi^10^, Sharmin Tamanna^10^, Khadija Akter Topy^10^, Umme Habiba Urmee^10^, Suraya Yesmin^10^, Shafia Zerin^10^

**Data Management Team**: Julianne Halley^10^, Catherine Perry^10^, Sarah Spackman^10^, Charlotte van Coeverden^10^, Matthew Walker^10^

**Laboratory Team**: Mahmuda Akther Akhi^10^,Asia Akter^10^, Labony Akter^10^, Ms Mili Akter^10^, Mst Shamima Akter^10^, Sabina Akter^10^, Salma Akter^10^, Setu Akter^10^, Tahmina Akter^10^, Md Sabdar Ali^10^, Mst Jesmin Ara^10^, Edyta Bujnik^10^, Jason Crawte^10^, Apu Chandra Das^10^, Many Das^10^, Samantha Farrow^10^, Nurjahan Fatema^10^, Md Riyad Hasan^10^, Md Saimul Islam^10^, Soniya Jannat^10^, Mst Amena Khatun^10^, Most Nurnahar Khatun^10^, William Mossman^10^, Robyn Murdoch^10^, Zannaton Naeem^10^, Danh Nguyen-Murray^10^, Evrikleia Ntasi^10^, Silvia Alonso Rodriguez^10^, Most Rezina Akter Roma^10^, Tripty Roy^10^, Most Abida Sultana^10^, Sharmin Akter Samia^10^

**Study Administration Team**: Nazneen Ali^10^, Laryssa Amado^10^, Tanya Braune^10^, Eilidh Cowan^10^, Steve Ellis^10^, Ellie Farrow^10^, Richard Houghton^10^, Md Zahidul Islam^10^, Giulia Loffreda^10^, Hannah Lombardi^10^, Ank Michielsen^10^, Niko Ovenden^10^, Tamara Sabri^10^, Karen Saunders neé Heasley^10^, Md Rafiqul Islam Rabbi^10^, Hosne Ara Rekha^10^, Valerie Rhenius^10^, Md Khalid Sultan^10^, Sophie Weston^10^, Hannah Williams^10^

**Scientific Investigators and Collaborators:** Tahmeed Ahmed^10^, Jim Ajioka^10^, Khondker Abdul Awal^10^, Aytalina Azarova^10^, Arul Baradi^10^, Adam S Butterworth^10^, Evangelia Chatzidiakou^10^, Sohel Reza Choudhury^10^, Rajiv Chowdhury^10^, Mayank Dalakoti^10^, John Danesh^10^, Emanuele Di Angelantonio^10^, Camilla Faidutti^10^, Joerg Feldmann^10^, Richard Fenner^10^, Meerjady Sabrina Flora^10^, Simon Griffin^10^, Louise Hair^10^, Sharifuddin Hasnat^10^, Sarah Hawkes^10^, Shahid Akhter Hossain^10^, Tuhin Haque^10^, Tafsir Hassan^10^, Md Mominul Islam^10^, Shafiul Islam^10^, Maria L C Iurilli^10^, Syed Shariful Islam^10^, Roderic L Jones^10^, Stephen Kaptoge^10^, Md Khalequzzaman^10^, Md AlfazalKhan^10^, Kamrul Hasan Khan^10^, Nusrat Khan^10^, Lawrence King^10^, Joe Lavallée^10^, Shammi Luhar^10^, Abdul Malik^10^, Fazila Tun-Nesa Malik^10^, Nick Mascie-Taylor^10^, Md Mostafa Monower^10^, Ruchira Tabassum Naved^10^, Md Sirajul Islam^10^, Aliya Naheed^10^, Anisur Rahman^10^, Lisa Pennells^10^, Olalekan Popoola^10^, Mahbubur Rahman^10^, Rubhana Raqib^10^, Laurie Savage^10^, Sara Shazad^10^, Tahmina Shirin^10^, Lalitha Sundaram^10^, Stephen Sutton^10^, Henry Taylor^10^, Aloka Tulukdar^10^, Kim Van Daalen^10^, Angela Wood^10^, Luisa Zuccolo^10^

**Fellows and Trainees:** Sadika Akhter^10^, Tanvir Chowdhury^10^, Nurul Huda^10^, Samia Naz Isha^10^, Riaz Hossain Khan^10^, Samsad Rabbani Khan^10^, Md Mostafa Monower^10^, Aliva Salmeen^10^, Zeeba Zahra Sultana^10^, Animesh Talukder^10^, Renesa Tarannum^10^

**External Advisory Board (CAPABLE Initiative):** Colin Baigent^10^, Abbas Bhuiya^10^, Shahida Parvin^10^

**Steering Committee & Data and Sample Access Committee:** Sohel Reza Choudhury^10^, John Danesh^10^, Emanuele Di Angelantonio^10^, Simon Griffin^10^, Md Khalequzzaman^10^, Md Alfazal Khan^10^, Nick Mascie-Taylor^10^

^10^BELIEVE Study, Dhaka, Bangladesh

**List of investigators from the Colorado Center for Personalized Medicine – RGC Collaboration**

All authors are listed in alphabetical order.

Heather D. Anderson^11,12^, Christina L. Aquilante^11,13^, Kelsey Arbogast^11^, Ian M. Brooks^11,14,15^, Elizabeth E. Burke^11,16^, Emily M. Casteel^11^, Joanne B. Cole^11,14^, Curtis R. Coughlin II^11,17^, Jacob Crawford^11^, Kristy Crooks^11,18^, Erin Culver^11^, Matthew J. Fisher^11^, Teresa C. Frye^11^, Hunter George^11^, Chris R. Gignoux^11,14^, Elizabeth K. Gilliland^11^, Casey S. Greene^11,14^, Emily Hearst^11,16^, Audrey E. Hendricks^11,14,19^, Randi K. Johnson^11,14,20^, Shelby Jones^11^, Dave Kao^11,21,16^, Gabrielle A. Knortz^11^, Danielle Koffenberger^11^, Santhanagopalan Krishnamoorthy^11^, Lisa Ku^11,22^, Elizabeth L. Kudron^11,14,17^, Rashawnda Lacy^11,15^, Ethan M. Lange^11,14^, Joe A. Lesny^11^, Meng Lin^11,14^, James L. Martin^11^, Nicole L. McDaniel^11,12^, Jack Pattee^11,23^, Nikita Pozdeyev^11,14,24^, Alaa Radwan^11,12^, Nick Rafaels^11^, Sridharan Raghavan^11^, Neda Rasouli^11,24^, Carolina Sanchez-Wild^11^, Elise L. Shalowitz^11^, Hoda Sherif^11^, Johnathan A. Shortt^11,14^, Adrian M. Stewart^11^, Carolyn T. Swartz^11,25^, Anna Tanaka^11,16^, Emily B. Todd^11,14^, Katy E. Trinkley^11,26^, Vendant Vohra^11^, Laura K. Wiley^11,14^

^11^Colorado Center for Personalized Medicine, Aurora, CO, USA

^12^Department of Clinical Pharmacy, University of Colorado Skaggs School of Pharmacy and Pharmaceutical Sciences, Anschutz Medical Campus, Aurora, CO, USA

^13^Department of Pharmaceutical Sciences, University of Colorado Skaggs School of Pharmacy and Pharmaceutical Sciences, Anschutz Medical Campus, Aurora, CO, USA

^14^Department of Biomedical Informatics, University of Colorado School of Medicine, Anschutz Medical Campus, Aurora, CO, USA

^15^Health Data Compass, Office of the Vice Chancellor for Health Affairs, Anschutz Medical Campus, Aurora, CO, USA

^16^CARE Innovation Center, UCHealth, Anschutz Medical Campus, Aurora, CO, USA

^17^Department of Pediatrics, University of Colorado School of Medicine, Anschutz Medical Campus, Aurora, CO, USA

^18^Department of Pathology, University of Colorado School of Medicine, Anschutz Medical Campus, Aurora, CO, USA

^19^Department of Mathematical and Statistical Sciences, College of Arts and Sciences, University of Colorado Denver Campus, Aurora, CO, USA

^20^Department of Epidemiology, Colorado School of Public Health, Anschutz Medical Campus

^21^Division of Cardiology, Department of Medicine, University of Colorado School of Medicine, Anschutz Medical Campus, Aurora, CO, USA

^22^Hereditary Cancer Clinic, UCHealth, Anschutz Medical Campus, Aurora, CO, USA

^23^Department of Biostatistics and Informatics, Center for Innovative Design and Analysis, Anschutz Medical Campus, Aurora, CO, USA

^24^Division of Endocrinology, University of Colorado School of Medicine, Anschutz Medical Campus, Aurora, CO, USA

^25^UCHealth Epic IT Department, Aurora, CO, USA

^26^University of Colorado Department of Family Medicine, University of Colorado School of Medicine, Anschutz Medical Campus, Aurora, CO, USA

**List of investigators from the GHS-RGC DiscovEHR collaboration**

All authors are listed in alphabetical order.

Adam Buchanan^27^, David J. Carey^27^, Christa L. Martin^27^, Michelle Meyer^27^, Kyle Retterer^27^, David Rolston^27^.

^27^Geisinger Health System, Danville, PA, USA

**List of investigators from the Mayo Clinic Project Generation (PG)**

All authors are listed in alphabetical order.

**PG Leadership Team**

James R. Cerhan^28^, Fergus J. Couch^28^, Janet E. Olson^28^

**Statistical Genetics and Bioinformatics**

Nicholas B. Larson^28^, Zachary S. Fredericksen^28^

**Registry Principal Investigators**

J. Eric Ahlskog^28^, Samuel O. Antwi^28^, Andrew D. Badley^28^, Jamie N. Bakkum-Gamez^28^, Suzette J. Bielinski^28^, Joanna M. Biernacka^28^, Lisa A. Boardman^28^, James H. Bower^28^, James R. Cerhan^28^, Mine Cicek^28^; Fergus J. Couch^28^, Angela Dispenzieri^28^, Sean C. Dowdy^28^, Jeanette E. Eckel Passow^28^, Gretchen E. Glaser^28^, Ellen L. Goode^28^, Peter C. Harris^28^, W. Michael Hooten^28^, Robert B. Jenkins^28^, Scott H. Kaufmann^28^, Richard B. Kennedy^28^, Iftikhar J. Kullo^28^, John C. Lieske^28^, Minetta C. Liu^28^, Daniel J. Ma^28^, Sarah A. McLaughlin^28^, Robert W. Mutter^28^, Janet E. Olson^28^, Naveen L. Pereira^28^, Vijay K. Ramanan^28^, Celine M. Vachon^28^, Prashanthi Vemuri^28^, Ping Yang^28^

**Management**

Lisa K. Colborn^28^, Andrew J. Danielsen^28^, Jonathan J. Harrington^28^, Jennifer M. Kushwaha^28^

^28^Mayo Clinic, Rochester, MN, USA

**List of investigators from the Mexico City Prospective Study**

Jason Torres^6^, Pablo Kuri-Morales^7,8^, Jaime Berumen^9^, Jesús Alegre-Díaz^9^, Rory Collins^30^, Roberto Tapia-Conyer^31^, Jonathan R. Emberson^30^

^6^Clinical Trial Service Unit and Epidemiological Studies Unit, Nuffield Department of Population Health, University of Oxford, Oxford, UK

^7^Instituto Tecnológico y de Estudios Superiores de Monterrey, Monterrey, Mexico

^8^Faculty of Medicine, National Autonomous University of Mexico, Mexico City, Mexico

^9^Experimental Research Unit from the Faculty of Medicine, National Autonomous University of Mexico, Mexico City, Mexico

^30^University of Oxford, Oxford, UK

^31^National Autonomous University of Mexico (UNAM), Mexico City, Mexico

**List of contributors from the Penn Medicine BioBank (PMBB)**

**PMBB Leadership Team**

Daniel J. Rader^32^, Marylyn D. Ritchie^32^

**Patient Recruitment and Regulatory Oversight**

JoEllen Weaver^32^, Nawar Naseer^32^, Giorgio Sirugo^32^, Afiya Poindexter^32^, Yi-An Ko^32^, Kyle P. Nerz^32^, Jenna Dever^32^, Aidan Harvey^32^, Sydney Linn^32^

**Lab Operations**

JoEllen Weaver^32^, Meghan Livingstone^32^, Fred Vadivieso^32^, Stephanie DerOhannessian^32^, Teo Tran^32^, Julia Stephanowski^32^, Salma Santos^32^, Ned Haubein^32^, Joseph Dunn^32^

**Clinical Informatics**

Anurag Verma^32^, Colleen Morse Kripke^32^, Marjorie Risman^32^, Renae Judy^32^, Colin Wollack^32^

**Genome Informatics**

Anurag Verma^32^, Shefali S. Verma^32^, Scott Damrauer^32^, Yuki Bradford^32^, Scott Dudek^32^, Theodore Drivas^32^

^32^Department of Genetics, Perelman School of Medicine, University of Pennsylvania, Philadelphia, PA, USA.

**List of contributors from the Regeneron Genetics Center**

**RGC Management & Leadership Team**

Aris Baras^1^, Gonçalo Abecasis^1^, Adolfo Ferrando^1^, Giovanni Coppola^1^, Andrew Deubler^1^, Luca A Lotta^1^, John D Overton^1^, Alan Shuldiner^1^, Katherine Siminovitch^1^, Jason Portnoy^1^, Marcus B Jones^1^, Lyndon Mitnaul^1^, Alison Fenney^1^, Jonathan Marchini^1^, Manuel Allen Revez Ferreira^1^, Maya Ghoussaini^1^, Mona Nafde^1^, William Salerno^1^, Cristen Willer^1^, Lourdes Crane^1^.

**Sequencing & Lab Operations**

John D Overton^1^, Christina Beechert^1^, Erin Fuller^1^, Laura M Cremona^1^, Eugene Kalyuskin^1^, Hang Du^1^, Caitlin Forsythe^1^, Zhenhua Gu^1^, Kristy Guevara^1^, Michael Lattari^1^, Alexander Lopez^1^, Kia Manoochehri^1^, Prathyusha Challa^1^, Manasi Pradhan^1^, Raymond Reynoso^1^, Ricardo Schiavo^1^, Maria Sotiropoulos Padilla^1^, Chenggu Wang^1^, Sarah E Wolf^1^

**Genome Informatics & Data Engineering**

Mona Nafde^1^, Manan Goyal^1^, George Mitra^1^, Sanjay Sreeram^1^, Rouel Lanche^1^, Vrushali Mahajan, Sai Lakshmi Vasireddy^1^, Gisu Eom^1^, Krishna Pawan Punuru^1^, Sujit Gokhale^1^, Benjamin Sultan^1^, Pooja Mule^1^, Mudasar Sarwar^1^, Muhammad Aqeel^1^, Xiaodong Bai^1^, Lance Zhang^1^, Sean O'Keeffe^1^, Razvan Panea^1^, Evan Edelstein^1^, Ayesha Rasool^1^, William Salerno^1^, Evan K Maxwell^1^, Boris Boutkov^1^, Alexander Gorovits^1^, Ju Guan^1^, Lukas Habegger^1^, Alicia Hawes^1^, Olga Krasheninina^1^, Samantha Zarate^1^, Adam J Mansfield^1^, Lukas Habegger^1^.

**Analytical Genetics and Data Science**

Gonçalo Abecasis^1^, Keyrun Adhikari^1^, Manuel Allen Revez Ferreira^1^, Joshua Backman^1^, Suganthi Balasubramanian^1^, Suying Bao^1^, Kathy Burch^1^, Adrian Campos^1^, David Corrigan^1^, Olivier Delaneau^1^, Liron Ganel^1^, Sheila Gaynor^1^, Benjamin Geraghty^1^, Arkopravo Ghosh^1^, Maya Ghoussaini^1^, Christopher Gillies^1^, Lauren Gurski^1^, Eric Jorgenson^1^, Tyler Joseph^1^, Michael Kessler^1^, Jack Kosmicki^1^, Alexander Lachmann^1^, Karl Landheer^1^, Adam Locke^1^, Priyanka Nakka^1^, Jonathan Marchini^1^, Anthony Marcketta^1^, Joelle Mbatchou^1^, Jonathan Ross^1^, Dhruv Shidhaye^1^, Carlo Sidore^1^, Eli Stahl^1^, Kathie Sun^1^, Mira Tang^1^, Rujin Wang^1^, Kuan-Han Wu^1^, Chen Wang^1^, Kyoko Watanabe^1^, Mark Weiner^1^.Frank Wendt^1^, Sean Yu^1^, Blair Zhang^1^, Chuanyi Zhang^1^, Andrey Ziyatdinov^1^, Yuxin Zou^1^

**Therapeutic Area Genetics**

Adolfo Ferrando^1^, Giovanni Coppola^1^, Luca A. Lotta^1^, Alan Shuldiner^1^, Katherine Siminovitch^1^, Kimberly Skead^1^, Brian Hobbs^1^, Jon Silver^1^, William Palmer^1^, Rita Guerreiro^1^, Amit Joshi^1^, Antoine Baldassari^1^, Cristen Willer^1^, Sarah Graham^1^, Ernst Mayerhofer^1^, Erola Pairo Castineira^1^, Mary Haas^1^, Niek Verweij^1^, George Hindy^1^, Jonas Bovijn^1^, Tanima De^1^, Luanluan Sun^1^, Olukayode Sosina^1^, Arthur Gilly^1^, Peter Dornbos^1^, Moeen Riaz^1^, Manav Kapoor^1^, Gannie Tzoneva^1^, Anna Alkelai^1^, Sahar Gelfman^1^, Vijay Kumar^1^, Jacqueline Otto^1^, Jose Bras^1^, Silvia Alvarez^1^, Jessie Brown^1^, Hossein Khiabanian^1^, Joana Revez^1^, Jae Soon Sul^1^, Lei Chen^1^, Sam Choi^1^, Amy Damask^1^, Nan Lin^1^, Charles Paulding^1^, Sameer Malhotra^1^, Joseph Herman^1^.

**Research Program Management & Strategic Initiatives**

Marcus B. Jones^1^, Michelle G. LeBlanc^1^, Nadia Rana^1^, Jennifer Rico-Varela^1^, Jaimee Hernandez^1^, Larizbeth Romero^1^, Ashley Paynter^1^.

**Senior Partnerships & Business Operations**

Randi Schwartz^1^, Lourdes Crane^1^, Alison Fenney^1^, Jody Hankins^1^, Anna Han^1^, Samuel Hart^1^, Ryan Smith^1^.

**Business Operations & Administrative Coordinators**

Ann Perez-Beals^1^, Gina Solari^1^, Johannie Rivera-Picart^1^, Michelle Pagan^1^, Sunilbe Siceron^1^.

^1^Regeneron Genetics Center, 777 Old Saw Mill River Rd., Tarrytown, New York, USA

**List of contributors from the UCLA-RGC ATLAS Collaboration**

**UCLA Precision Health ATLAS**

Alex Bui^33^, Antonia Petruse^33^, Arash Naeim^33^, Chris Denny^33^, Clara Lajonchere^33^, Clara Magyar^33^,

Dan Geschwind^33^, Maryam Ariannejad^33^, Paul Boutros^33^, Paul Spellman^33^, Sarah Dry^33^, Stan Nelson^33^.

**UCLA Precision Health Data Discovery**

Albert Duntugan^33^, Alex Bui^33^, Bogdan Pasaniuc^33^, Chris Denny^33^, Clara Lajonchere^33^, Dan Geschwind^33^, Paul Boutros^33^, Paul Tung^33^, Tim Chang^33^, Yael Berkovich^33^.

**UCLA IT High Performance Computing**

Albert Duntugan^33^, Ankur Jain^33^, Danielle Martinez^33^, Ghouse Mohammed^33^, Lora Illiev^33^, Michael Broudy^33^, Paul Boutros^33^, Paul Tung^33^, Roni Haas^33^, Shaiful Alam^33^, Taka Yamaguchi^33^, Yael Berkovich^33^, Yash Patel^33^

^33^Institute of Precision Health, University of California, Los Angeles, Los Angeles, CA, USA

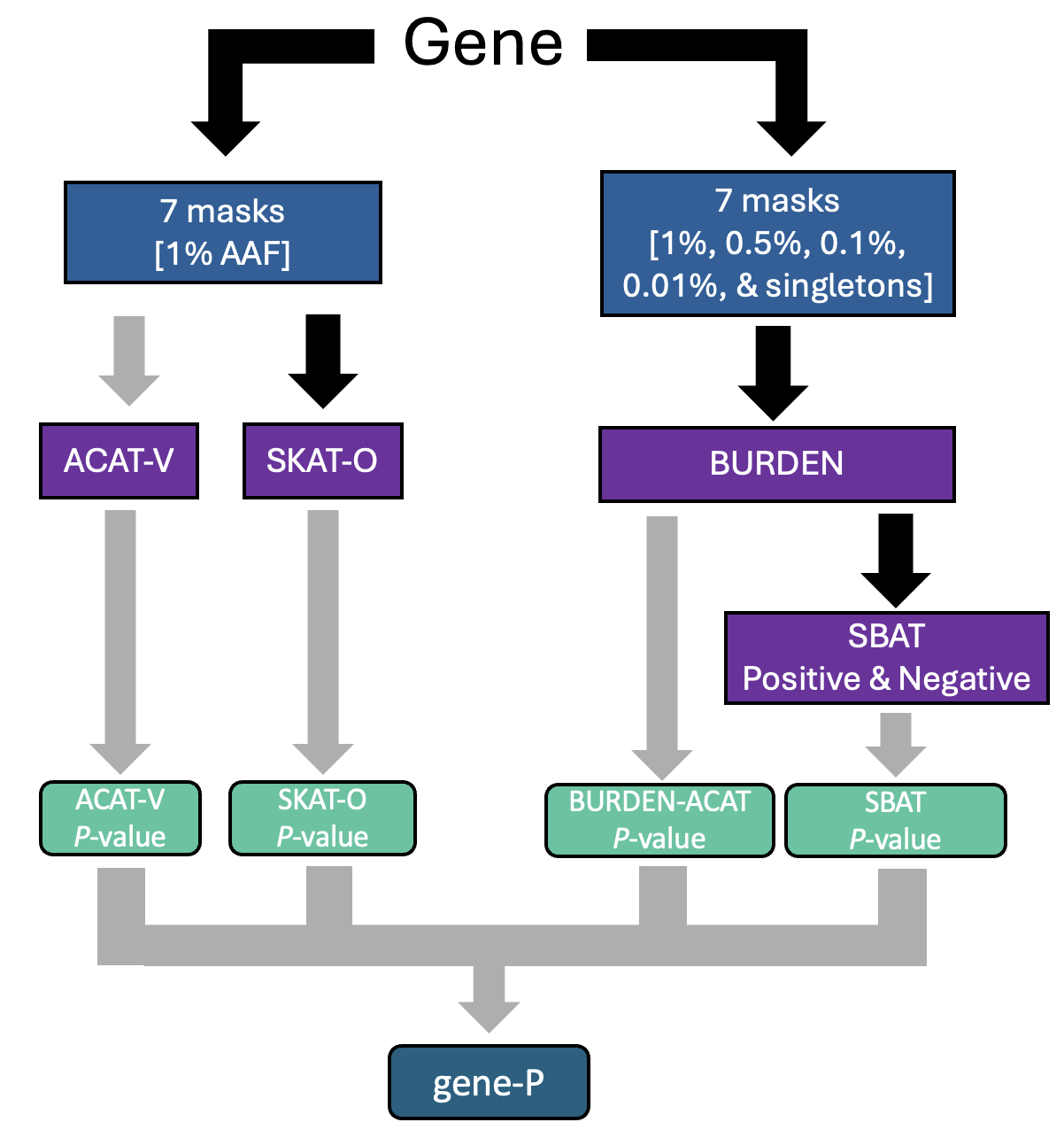

**Supplementary Figure 1.** Unified gene-P test flowchart. Gene-based test associations focused on a single, unified *P*-value per gene (the gene-P *P*-value), which aggregates across multiple gene-based test methods, alternate allele frequencies, and masks comprising different combinations of variant classes. The flowchart visualizes how, for a given gene, individual variants are combined and tested to yield a single *P*-value. Gray arrows indicate aggregation by ACAT^14^.

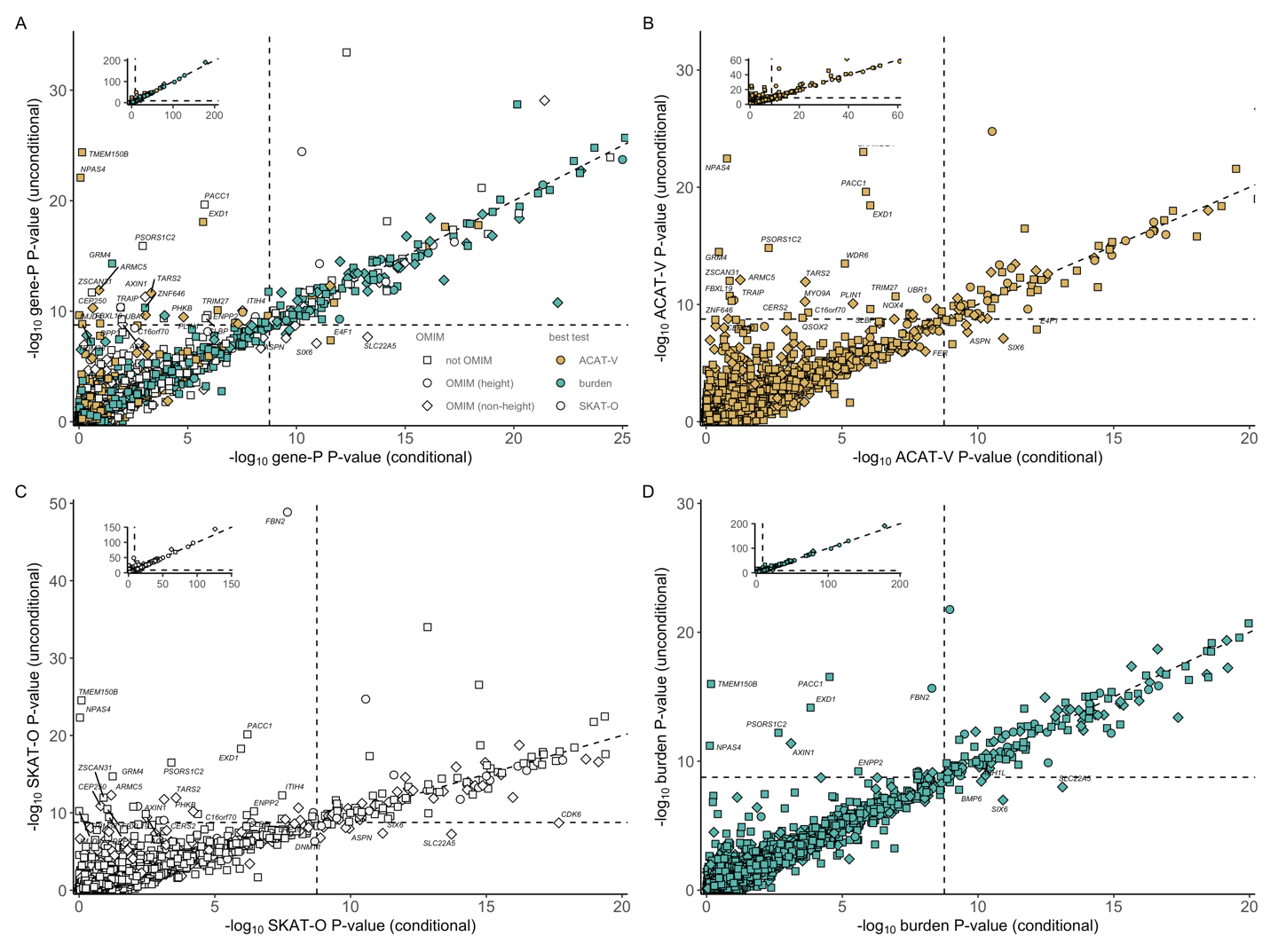

**Supplementary Figure 2**: **A**) -log_10_ gene-P *P*-values for all genes pre- and post-conditioning on 3034 independent common variants (**Supplementary Table 2**). Genes are colored based on which of the gene-based tests that comprise gene-P was most significant. **B)** -log_10_ ACAT-V *P*-values for all genes pre- and post-conditioning. **C**) -log_10_ SKAT-O *P*-values for all genes pre- and post-conditioning. **D**) -log_10_ burden test (the minimum of SBAT and burden ACAT) for all genes pre- and post-conditioning. The full distribution is present in the inset. Each point represents a gene and dashed lines demarcate the Bonferroni significance threshold of 1.75x10^-9^ throughout **A**-**D**.

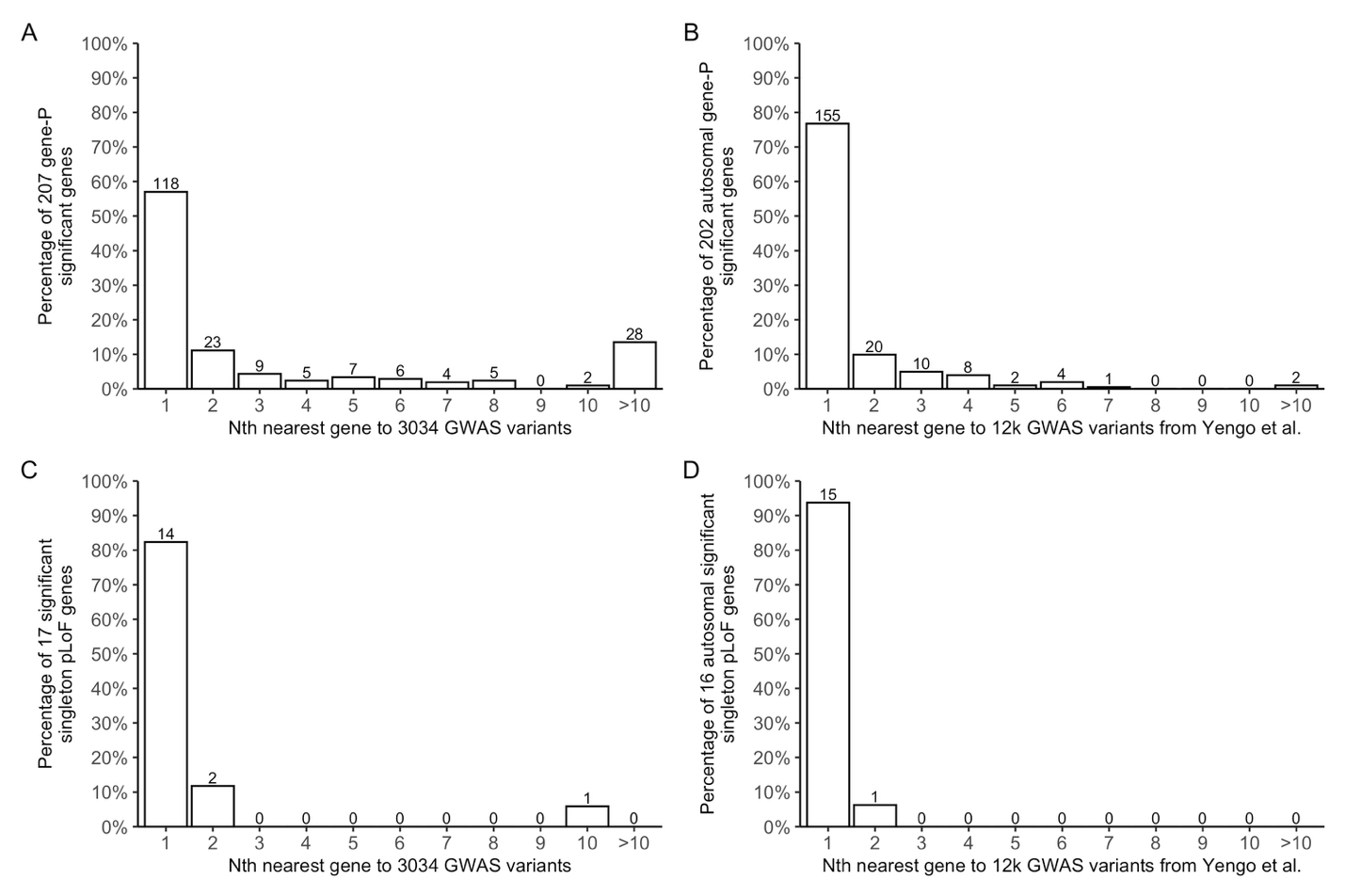
**Supplementary Figure 3**. Proximity of genes discovered via rare variant gene-based tests to GWAS variants. Panels **A** and **C** use 3034 conditionally independent GWAS loci discovered in this study (**Supplementary Table 2**) and panels **B** and **D** use 12,111 autosomal variants from the largest GWAS of height^1^. The 207 gene-P genes (**Supplementary Table 4**) are depicted in panel **A** and the 202 autosomal gene-P genes are in panel **B** (as the Yengo *et al.* GWAS only covered the autosomes). The 17 genes with a burden of singleton pLoFs (**Supplementary Table 5**) are depicted in **C** and **D** (*NRK* is excluded from panel **D** due to being on the X-chromosome as Yengo *et al*. GWAS only report autosomal loci).

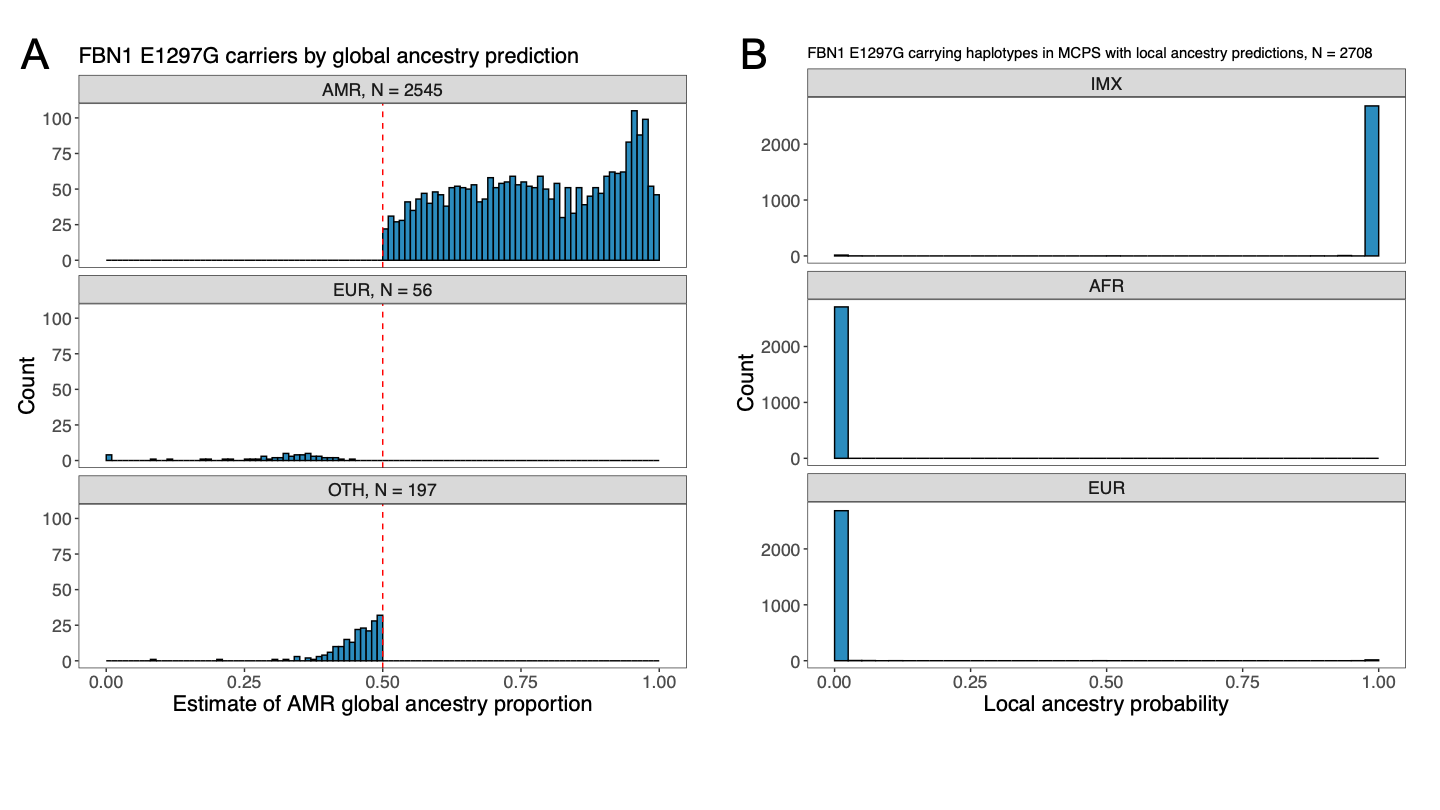

**Supplementary Figure 4**. **A)** Global AMR ancestry proportion estimates among 2798 carriers of *FBN1* p.E1297G (15:48481729:T:C) in discovery data. Individuals were assigned to a genetic ancestry group using a threshold of 50% for any one continental category (**Methods**). Those not reaching that threshold in any category were assigned to the OTH group. The red dashed line represents the 50% threshold. **B)** Local ancestry proportions of *FBN1* p.E1297G carriers in MCPS from Ziyatdinov *et al.*^7^ where such data were available. These predictions are made on phased haplotype data, so each haplotype is treated separately (22 homozygotes were observed). AMR – Admixed American, EUR – European, OTH – admixed, IMX – indigenous Mexican.

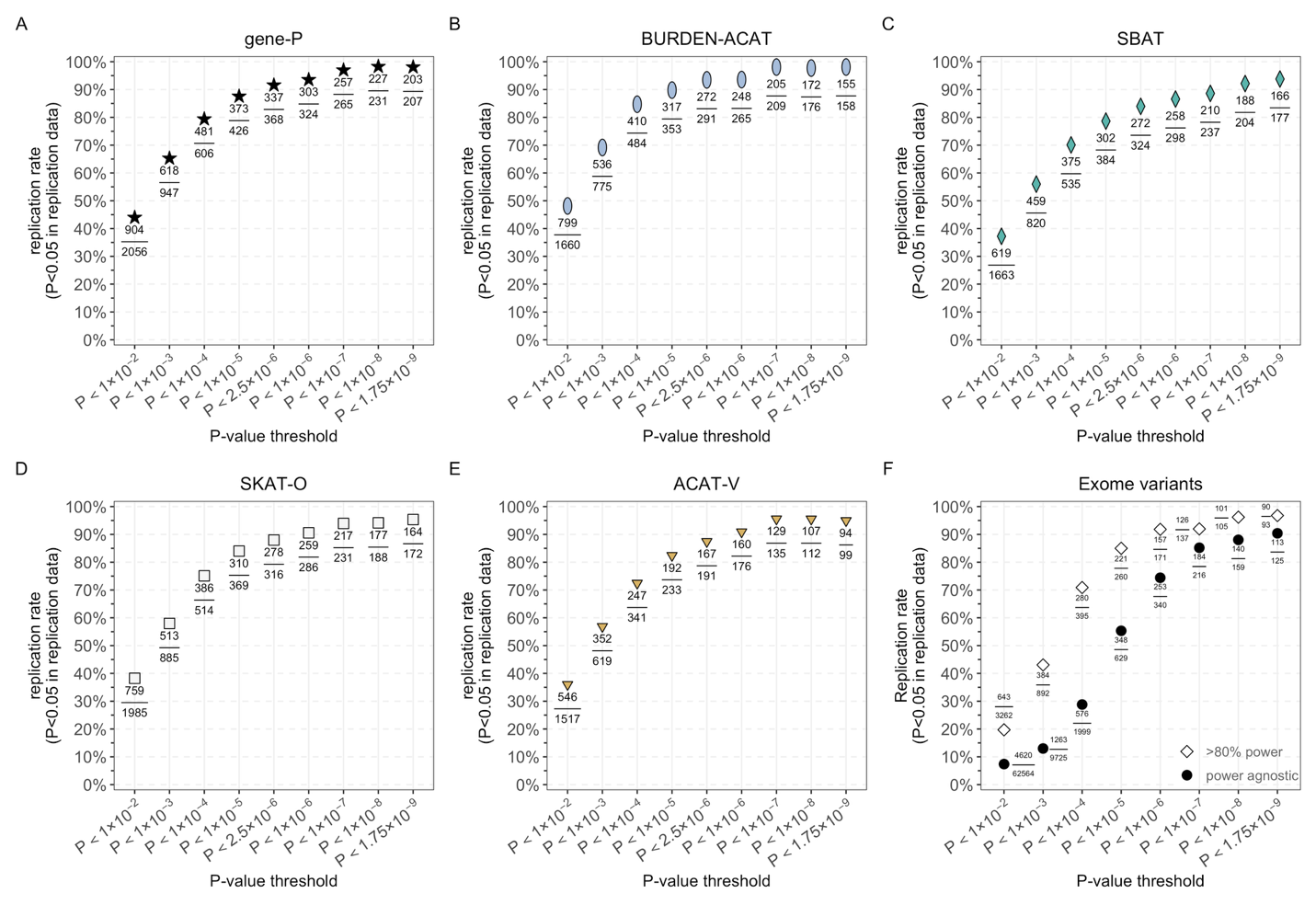

**Supplementary Figure 5**. Number of associations identified (numbers presented next to each point) in the discovery dataset (N=826,066) across *P*-value thresholds and replication rate (points; *P* < 0.05 in N=624,567) at each *P*-value threshold for gene-based tests: (**A**) gene-P, (**B**) Burden-ACAT, (**C**) SBAT, (**D**) SKAT-O, and (**E**) ACAT-V. (**F**) The replication rate for individual exome variants (AAC>9; in the exome target region; not imputed) present in at least one individual in the replication cohort are represented by black circles. The subset of variants with >80% power is depicted by white diamonds.

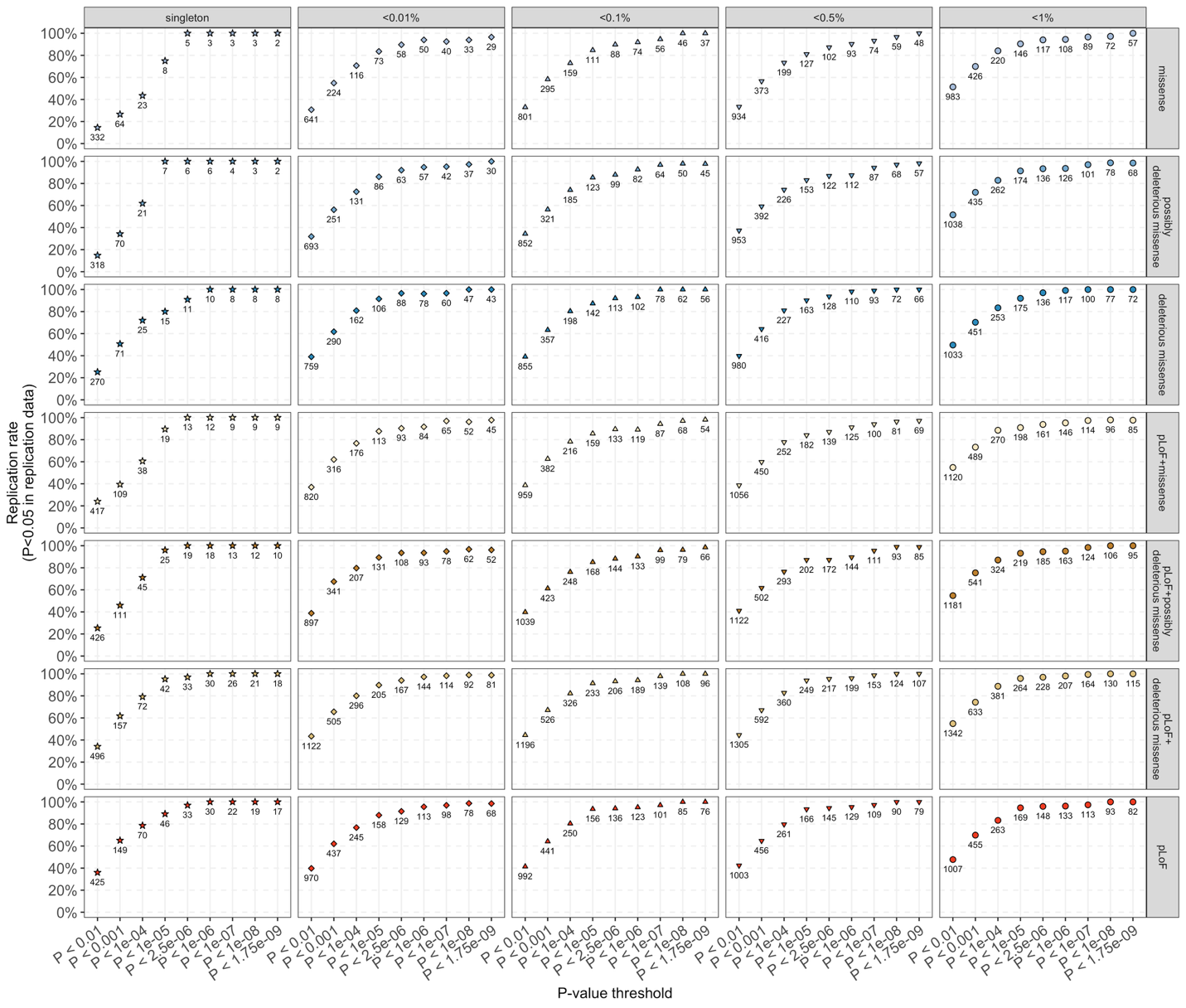

**Supplementary Figure 6**: Number of burden tests identified (numbers presented next to each point) in the discovery (N=826,066) and replication rate (points; *P* < 0.05 in N=624,567) across *P*-value thresholds for 35 sets of burden tests (seven combinations of variant classes and five AAF thresholds).

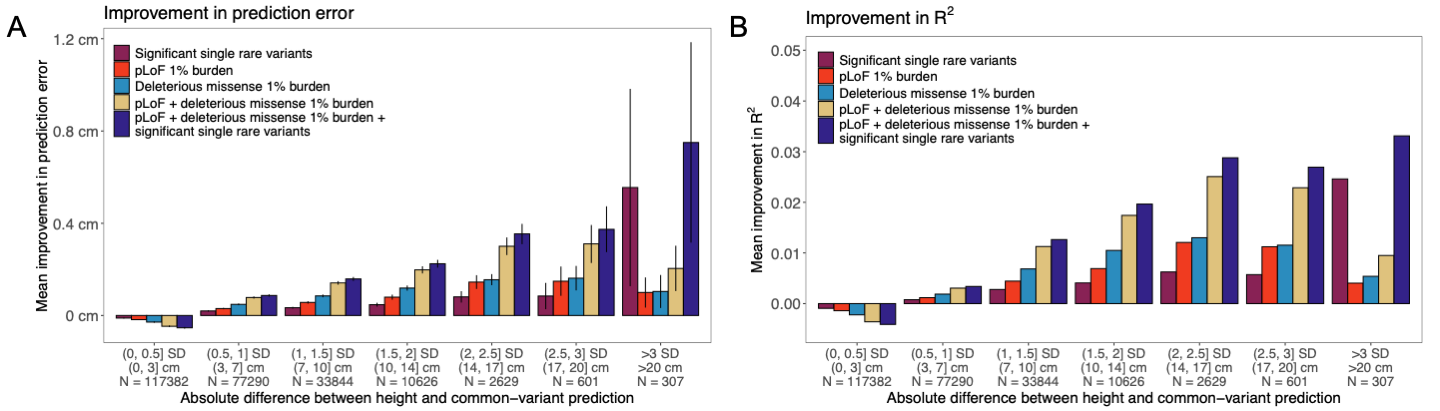

**Supplementary Figure 7**: Average improvement in height prediction when adding rare variants. **A)** difference (cm) between the predictive error from common variants and the predictive error including rare variants. **B)** Difference between R^2^ from common variants and R^2^ when using common and rare variants. R^2^ is calculated for the univariate linear regression of observed height on predicted height. In both panels, X axes are split based on the same common-variant prediction bins as in **Figure 3A**. All predictions are performed using common variants, rare variants, and covariates (**Methods**).

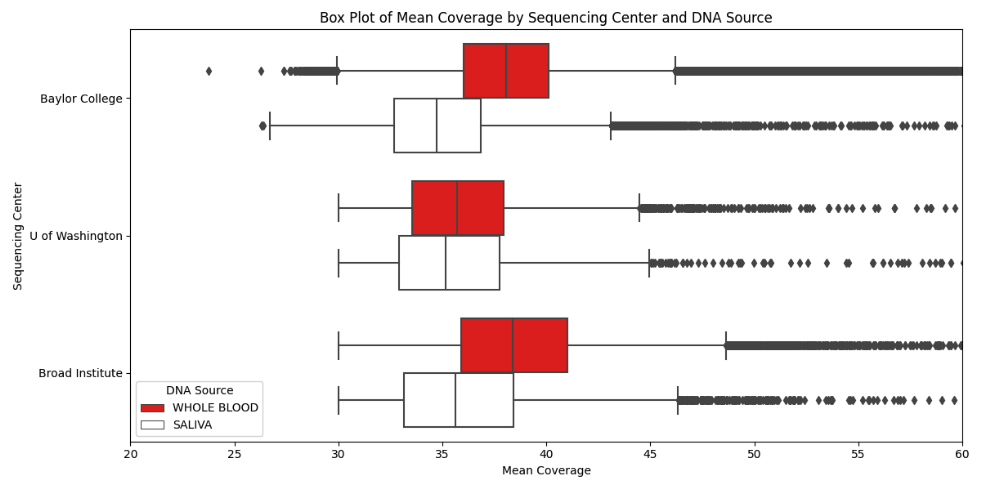

**Supplementary Figure 8:** Box plots of sequencing coverage across three sequencing centers and two DNA sources (whole blood in red, saliva in white) in All of Us. We created three batch covariates to address confounding (**Supplementary Table 24**). Outlier individuals are represented by points.

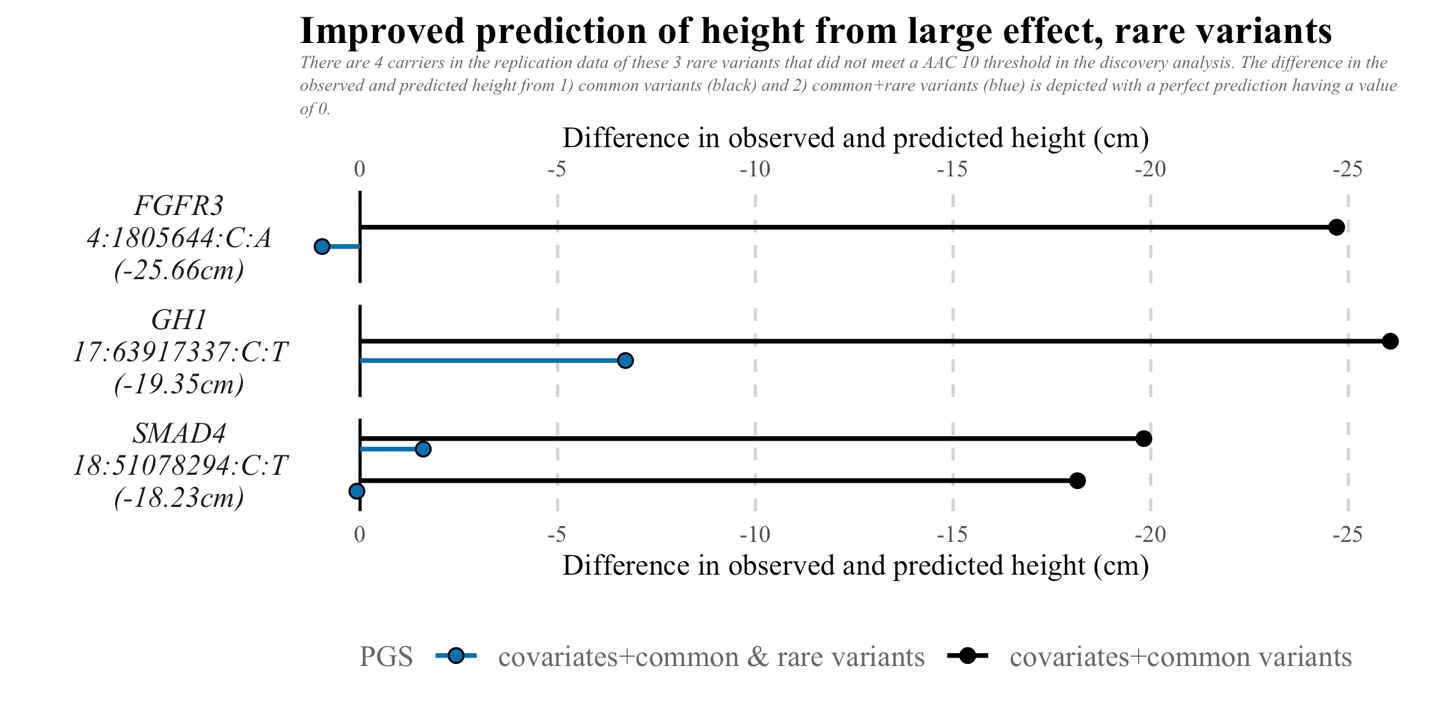

**Supplementary Figure 9**: There are four carriers in the replication data for these three rare variants that did not meet an AAC>9 threshold in the discovery analysis. The difference in the observed and predicted height from 1) common variants (black circles) and 2) common + rare variants (blue circles) are depicted with the distance from a perfect prediction (vertical line at 0).

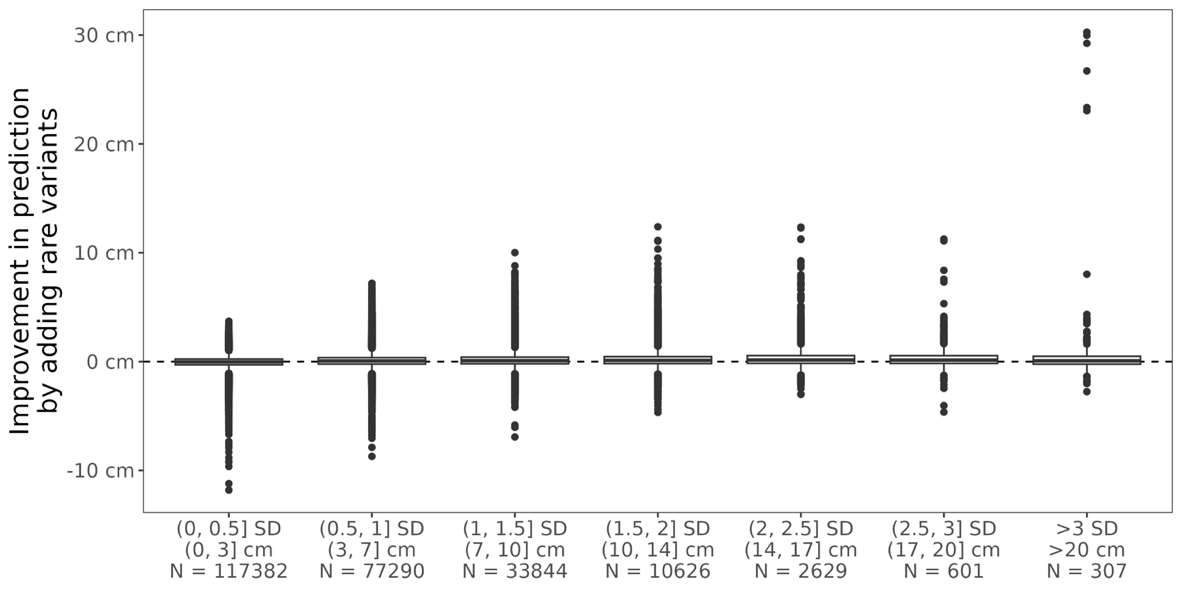

**Supplementary Figure 10**: Improvement in absolute predictive error from adding rare variants into the prediction model. These results reflect the predictions from our primary rare-variant prediction model, which includes rvPGS_SNP_, rvPGS_pLoF_, and rvPGS_missense_ (**Methods**). The outliers with > 3 SD |predictive error| and large improvement in prediction are carriers of a substitution in *FGFR3* resulting in a p.G380R amino acid change.

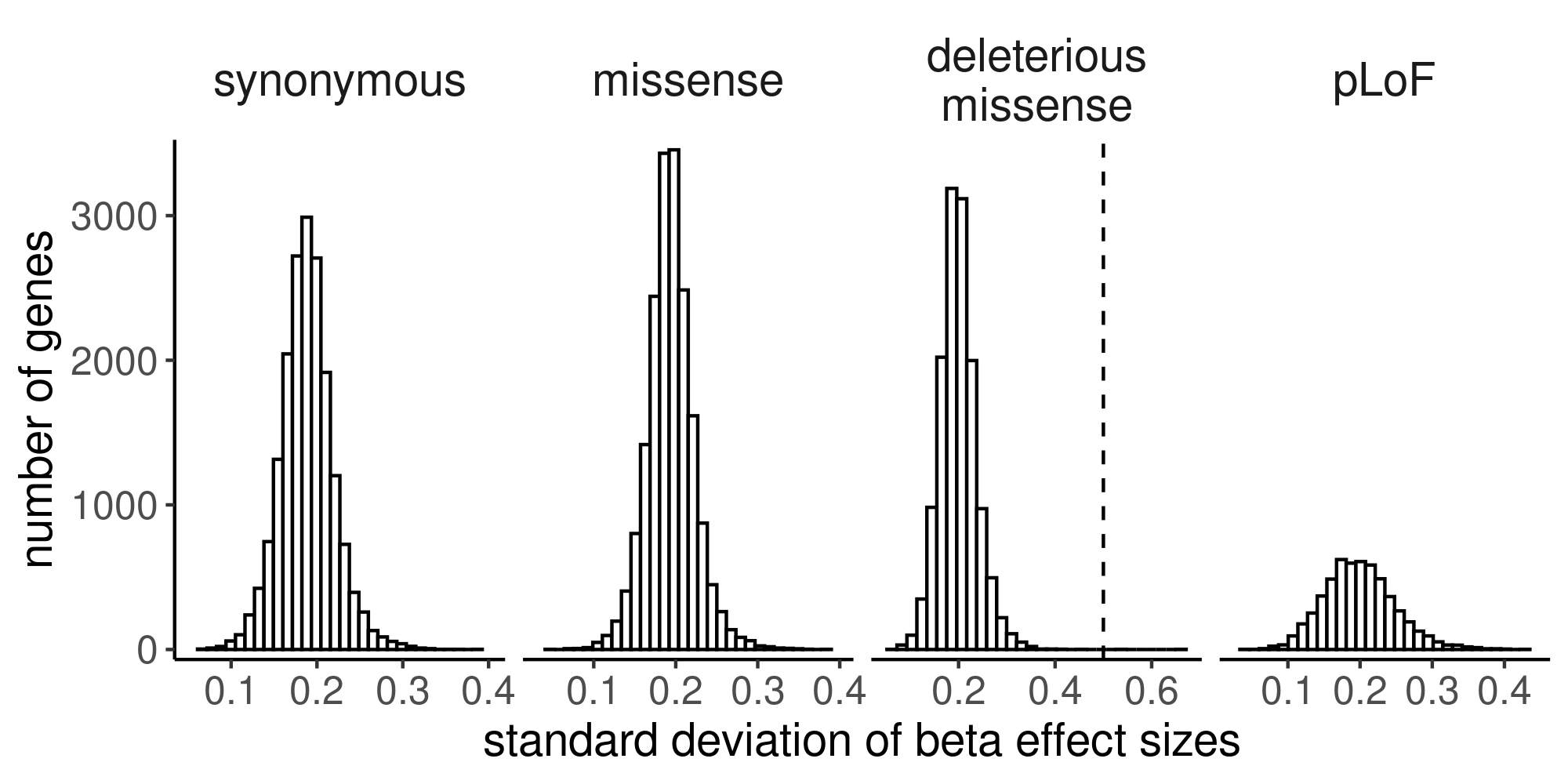

**Supplementary Figure 11**: distribution of absolute effect sizes (in standard deviation units) per variant class across all genes. Dotted black line demarcates 12 SDs beyond the mean.

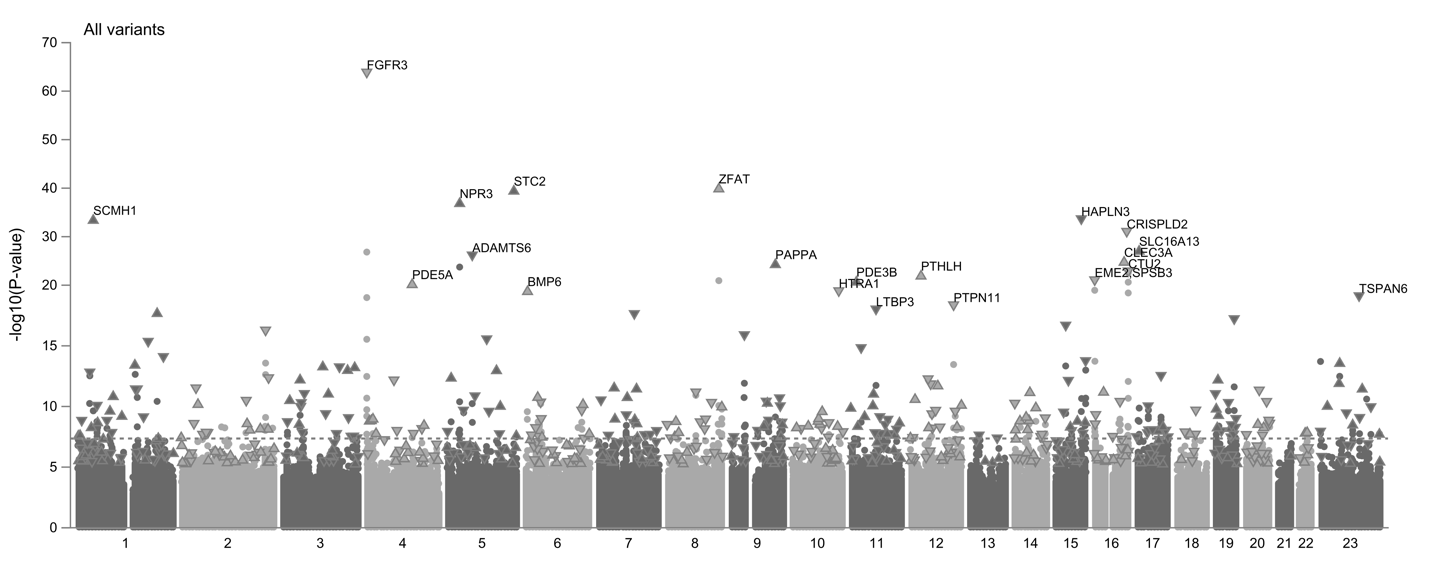
**Supplementary Figure 12**. Manhattan plot of individual exome variants (dots; **Supplementary Table 3**). Dashed black line demarcates the Bonferroni-significant threshold of 1.75x10^-9^. *P*-values come from REGENIE.

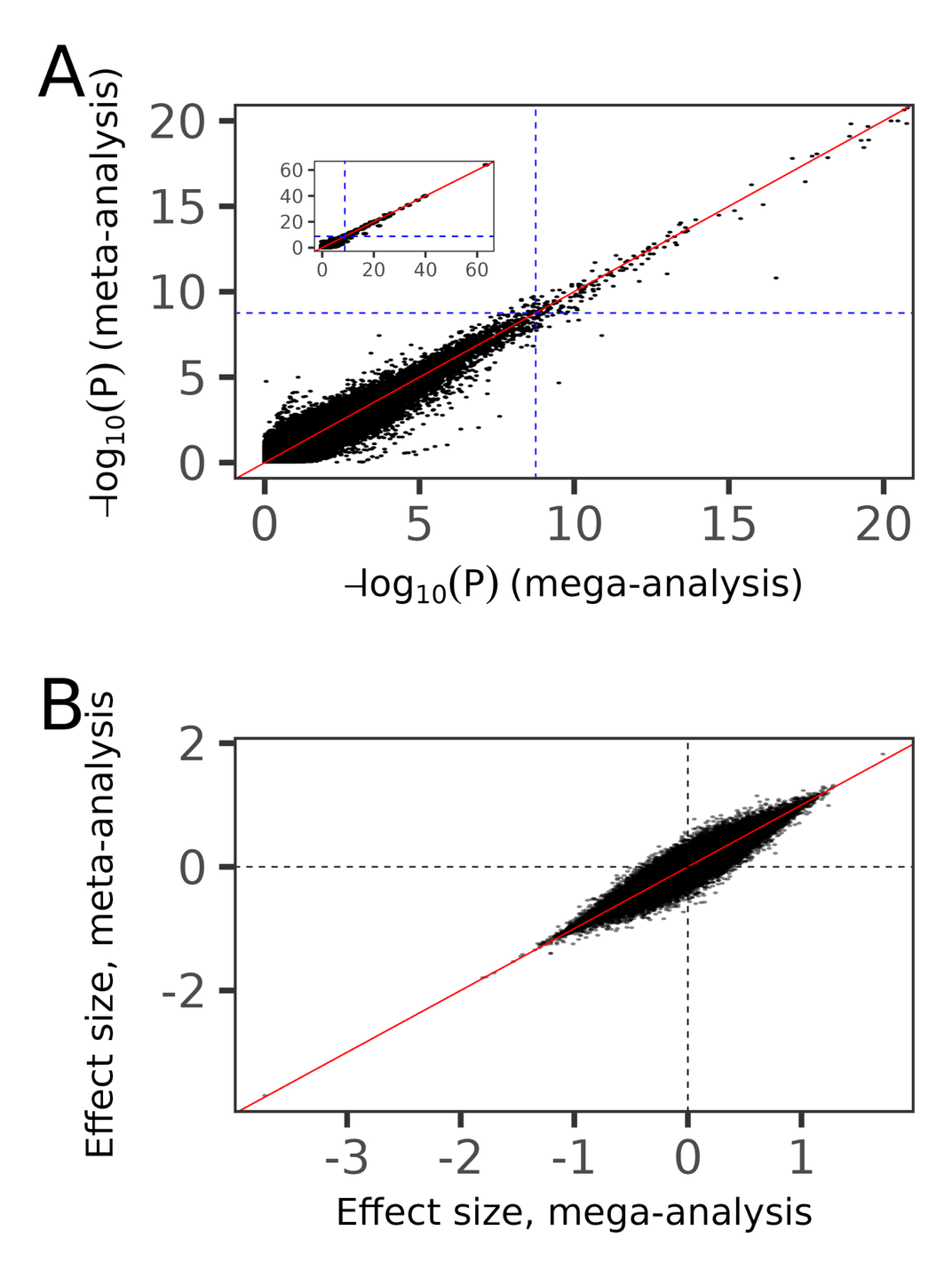

**Supplementary Figure 13**. **A**) Comparison of single-variant (AAC > 9 in both analyses) *P*-values between a mega- and meta-analysis. The inset shows the full range of values. **B)** Comparison of effect sizes (in SD units) between a mega- and a meta-analysis. Each dot represents a variant. Diagonal red lines represent the identity line in all panels, and dashed blue lines represent the experiment-wide significance threshold of 1.75×10^-9^.

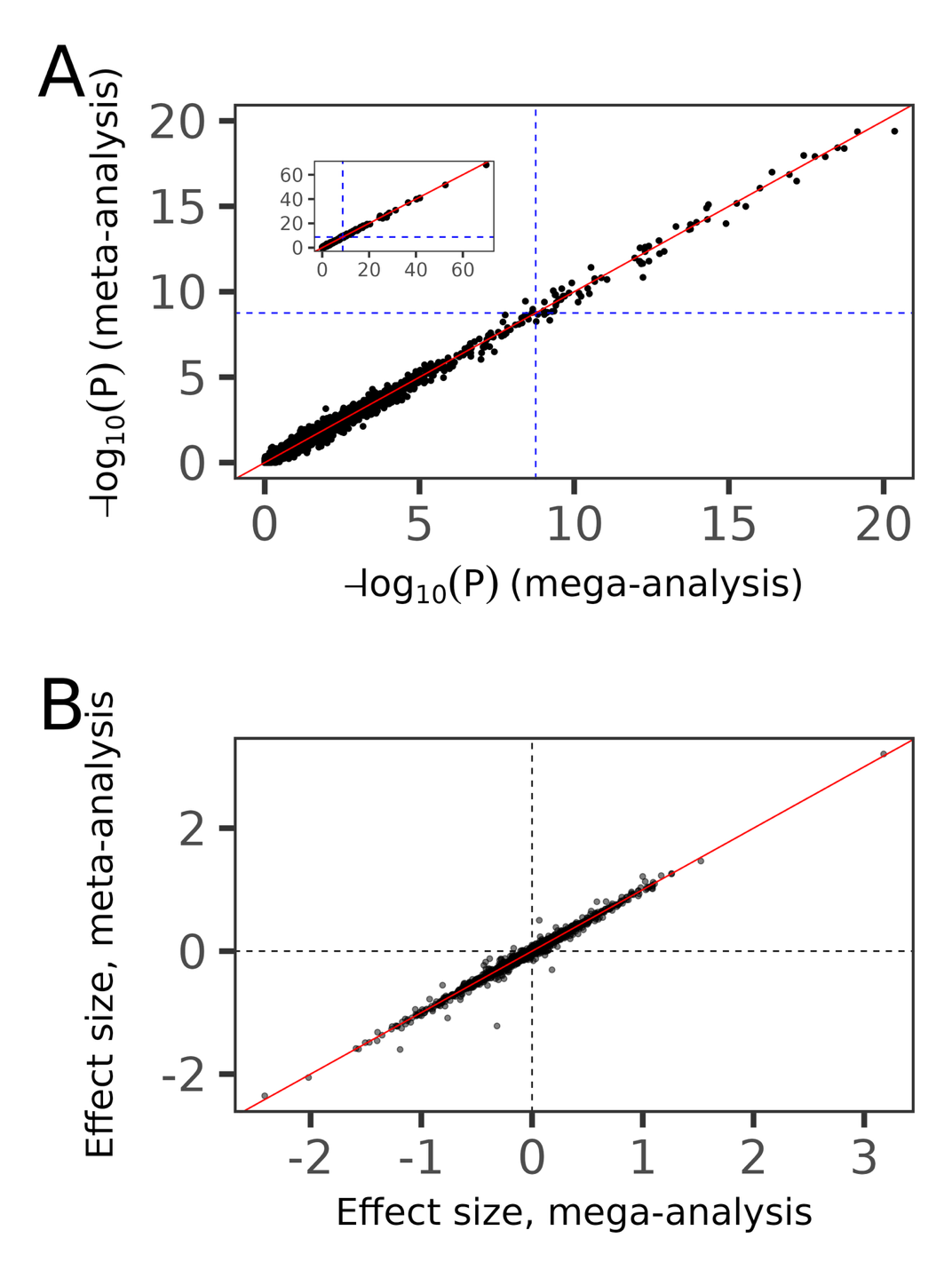

**Supplementary Figure 14**. **A**) Comparison of *P*-values for pLoF burden tests (AAF<1%) between a mega- and meta-analysis. The inset shows the full range of values. **B)** Comparison of pLoF burden test (AAF<1%) effect sizes (in SD units) between a mega- and a meta-analysis. Each point represents a burden test. Diagonal red lines represent the identity line in all panels, and dashed blue lines represent the experiment-wide significance threshold of 1.75×10^-9^. *P*-values come from REGENIE.

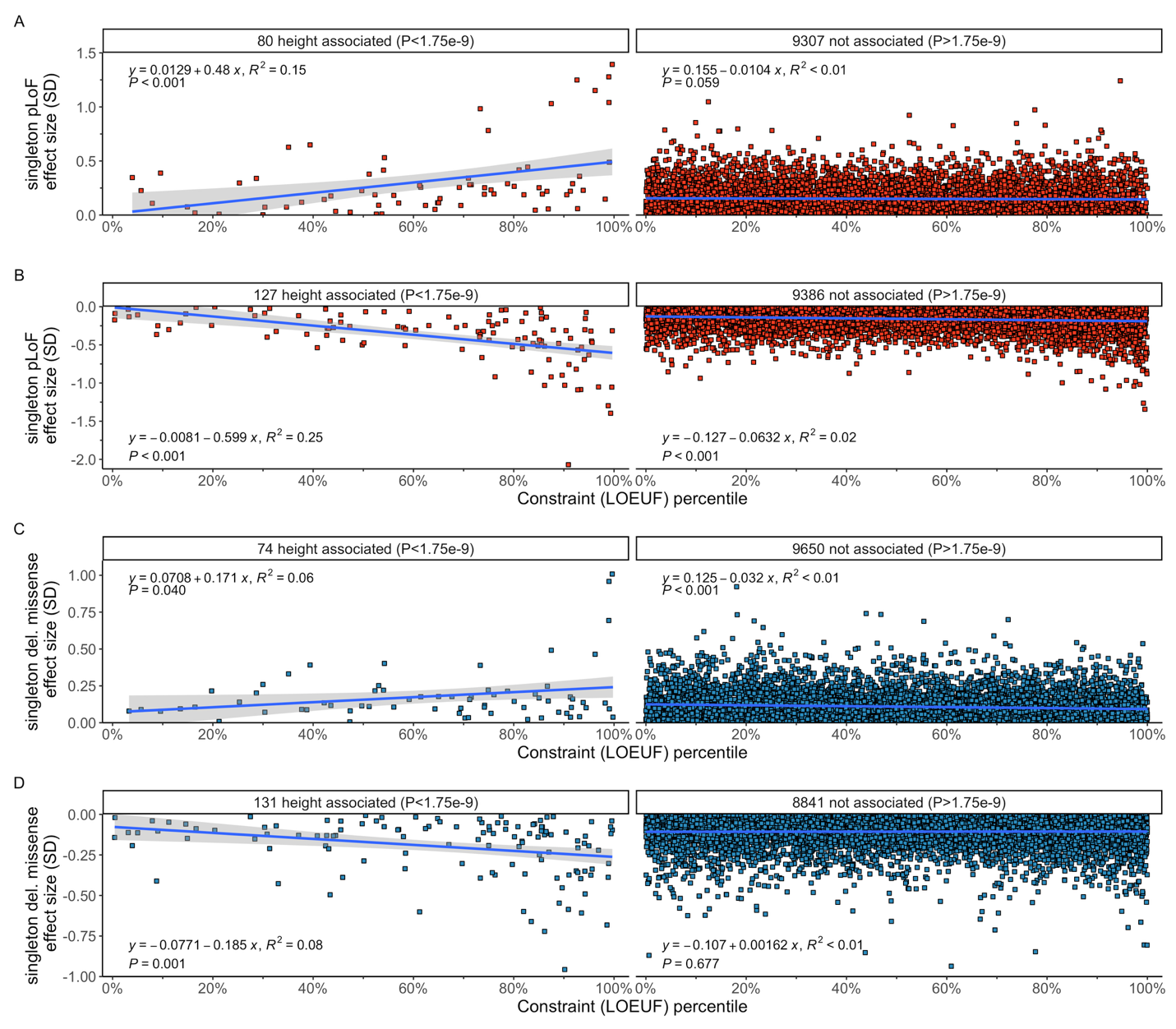
**Supplementary Figure 15**. Correlation between genic constraint (measured using loss-of-function observed/expected upper-bound fraction [LOEUF]^11^) and pLoF singleton burden (**A** and **B**) and deleterious missense (**C** and **D**) effect sizes (in standard deviations [SD]). Each point is a gene and genes are separated by gene-P significance (⍺ = 1.75×10^-9^) and burden effect size direction.

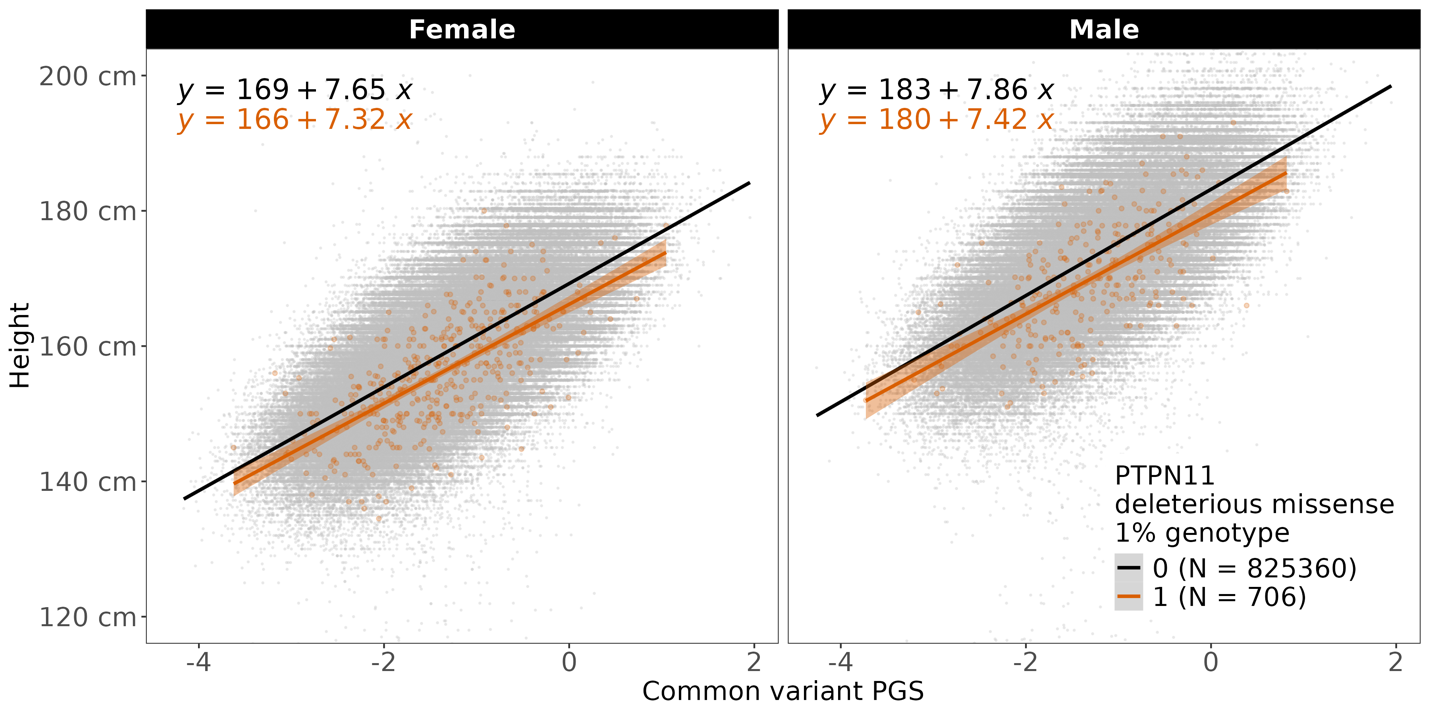

**Supplementary Figure 16**: Interaction effects of *PTPN11* rare deleterious missense variant burden with common-variant PGS, separated by sex. Noncarriers are shown in gray points and carriers are shown in orange. Lines represent univariate linear regression lines of height on cvPGS. The black lines and equations show this regression for noncarriers, and the orange ones describe the regression for carriers.
